# *APOE* stratified genome-wide association studies provide novel insights into the genetic etiology of Alzheimers’s disease

**DOI:** 10.1101/2025.05.07.25327065

**Authors:** Jesper Qvist Thomassen, Hampton Leonard, Brittany Ulms, Benjamin Grenier-Boley, Sami Heikkinen, Pablo Garcia-González, Atahualapa Castillo-Morales, Masataka Kikuchi, Jungsoo Gim, Han Cao, Fahri Küçükali, Najaf Amin, Dabin Yoon, Itziar de Rojas, Pilar Alvarez Jerez, Victoria Alvarez, Beatrice Arosio, Céline Bellenguez, Sverre Bergh, Kimberley Billingsley, Cornelis Blauwendraat, Merce Boada, Barbara Borroni, Paola Bossù, María J. Bullido, Antonio Daniele, Ángel Carracedo, Alexandre de Mendonça, Mark Cookson, Jürgen Deckert, Martin Dichgans, Srdjan Djurovic, Oriol Dols-Icardo, Carole Dufouil, Emrah Düzel, Valentina Escott-Price, Tormod Fladby, Laura Fratiglioni, Amy K.Y. Fu, Daniela Galimberti, Jose Maria García-Alberca, Vilmantas Giedraitis, Guillermo Garcia-Ribas, Caroline Graff, Timo Grimmer, Edna Grünblatt, OIivier Hanon, Lucrezia Hausner, Stefanie Heilmann-Hemibach, Jakub Hort, Frank Jessen, Kendall Jensen, Caroline Jonson, Yoontae Kim, Nicole Kuznetsov, Ville Leinonen, Anssi Lipponen, Jiao Luo, Mary Makarious, Henna Martiskainen, Carlo Masullo, Patrizia Mecocci, Shima Mehrabian, Pablo Mir, Akinori Miyashita, Susanne Moebus, Kin Y. Mok, Laura Molina Porcel, Fermin Moreno, Benedetta Nacmias, Lucilla Parnetti, Pau Pastor, Jordi Pérez-Tur, Oliver Peters, Yolande A.L. Pijnenburg, Gerard Piñol-Ripoll, Julius Popp, Innocenzo Rainero, Luis M Real, Steffi Riedel-Heller, Eloy Rodriguez-Rodriguez, Arvid Rongve, Giacomina Rossi, Jose Luis Royo, Dan Rujescu, Ingvild Saltvedt, María Eugenia Sáez, Raquel Sánchez-Valle, Florentino Sanchez-Garcia, Nicolai Sandau, Nikolaos Scarmeas, Katja Scheffler, Norbert Scherbaum, Anja Schneider, Geir Selbæk, Davide Seripa, Vincenzo Solfrizzi, Marco Spallazzi, Alessio Squassina, Eystein Stordal, Niccoló Tesi, Lucio Tremolizzo, Kumar P. Tripathi, Wiesje M. van der Flier, Julie Williams, Jens Wiltfang, Dag Aarsland, Andrew B Singleton, Philippe Amouyel, Stéphanie Debette, Gael Nicolas, Sven van der Lee, Henne Holstege, Maria Victoria Fernandez, Patrick Gavin Kehoe, Kristel Sleegers, Martin Ingelsson, Roberta Ghidoni, Ole A. Andreassen, Peter A. Holmans, Pascual Sánchez-Juan, Rebecca Sims, Nancy Y. Ip, Kun Ho Lee, Takeshi Ikeuchi, Alfredo Ramirez, Agustin Ruiz, Mikko Hiltunen, Jean-Charles Lambert, Cornelia van Duijn, Mike Nalls, Ruth Frikke-Schmidt

## Abstract

Among the more than 90 identified genetic risk loci for late-onset Alzheimer’s disease (AD) and related dementias, the apolipoprotein E gene (*APOE*) ɛ2/ɛ3/ɛ4 polymorphisms remains the longstanding benchmark for genetic disease risk with a consistently large effect across studies^1–10^. Despite this massive signal, the exact mechanisms for how ɛ4 increases and for how ɛ2 decreases dementia risk is not well-understood. Importantly, recent trials of anti-amyloid therapies suggest less efficacy and higher risks of severe side effects in ε4 carriers^11–13^, hampering the treatment of those with the highest unmet need. To improve our understanding of the genetic architecture of AD in the context of its main genetic driver, we performed genome-wide association studies (GWASs) stratified by ε4 and ε2 carrier status. Such insights may help to understand and overcome side effects, to impact clinical trial enrolment strategies, and to create the scientific basis for targeted mechanism-driven therapies in neurodegenerative diseases.

---

The present work is the largest meta-analysis GWAS attempt to provide the most informative overview of the genetics of AD according to *APOE* ɛ2/ɛ3/ɛ4 stratification, bringing together European, Asian, Asian-American, African-American, and admixed American ancestry cohorts based on clinically diagnosed AD. An overview of the included consortia and cohorts, as well as the analysis strategy is given in Figure 1. Individuals were grouped in ɛ22+ε32, ε33 and ε44+ε43 strata to maximize statistical power and individuals with the ε42 genotype were excluded (Supplementary Tables 1-2). In the ɛ22+ε32 stratum, the meta-analysis was based on 2,606 AD cases, 70,388 controls and 10,336,311 variants (Supplementary Table 3, Supplementary Figure 1). In the ε33 stratum, the meta-analysis was based on 24,033 AD cases, 363,161 controls and 17,127,662 variants (Supplementary Table 4, Supplementary Figure 1). Finally, in the ε44+ε43 stratum, the meta-analysis was based on 29,122 AD cases, 164,206 controls and 14,672,059 variants (Supplementary Table 5, Supplementary Figure 1; Supplementary Tables 6 and Supplementary Figure 1 and Supplementary Figure 2 for strata ε44).

**Figure 1.**
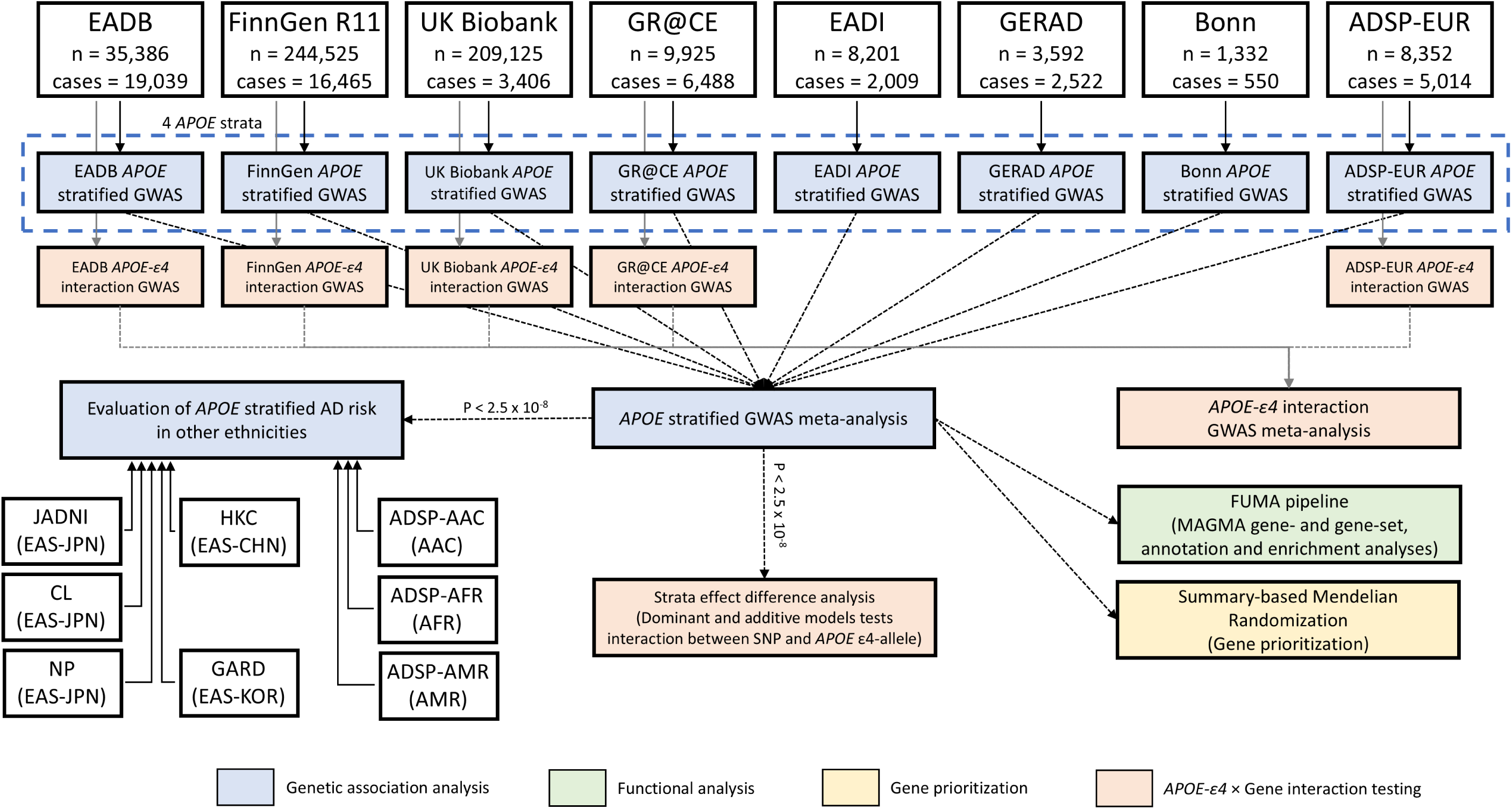
Flowchart of *APOE* stratified GWAS analysis plan. Eight European, five East Asian and three American cohorts were included. Genetic association analysis (genome-wide association study (GWAS)) was performed in four *APOE* strata (ε33, ε44, ε43+ε44, ε22+ε32). Genome-wide significant signals was tested with functional analysis, genetic characterization, and gene-gene interaction analysis.

For the ε22+ε32 stratum no signals reached a genome-wide significance level of <2.5×10^-8^ (Supplementary Figure 3). In total, 25 loci reached a genome-wide significance level in strata ε33 or ε44+ε33 only or in both (Figure 2, Table 1, Supplementary Figures 4-24). For the nine loci found in both strata, they are well known genetic risk loci associated with AD: *CR1*, *BIN1*, *HLA*, *TREM2*, *CLU*, *MS4A64*, *PICALM*, *APH1B*, *ABCA7*. Nine loci were exclusively observed in the ε44+ε43 stratum, 6 are known AD loci: *PILRA*, *SORL1*, *ADAM10*, *ACE*, *LILRA5*, *CASS4* and 3 loci are novel AD loci: *HP1BP3*, *PTPRC, DDHD1.* Notably, *DDHD1* is close to the *FERMT2* locus, which is recognized as a genetic risk factor for AD. However, using conditional testing, we found that the *DDHD1* signal is independent of *FERMT2* (Figure 2, Supplementary Table 7). Finally, among the 7 loci only reaching genome wide significance level in the ε33 stratum, 4 are known as AD risk loci: *TMEM106B, SHARPIN, GRN, MAPT,* and 3 loci are novel: *SCL50A1, NPAS3, CHST9*.

**Figure 2:**
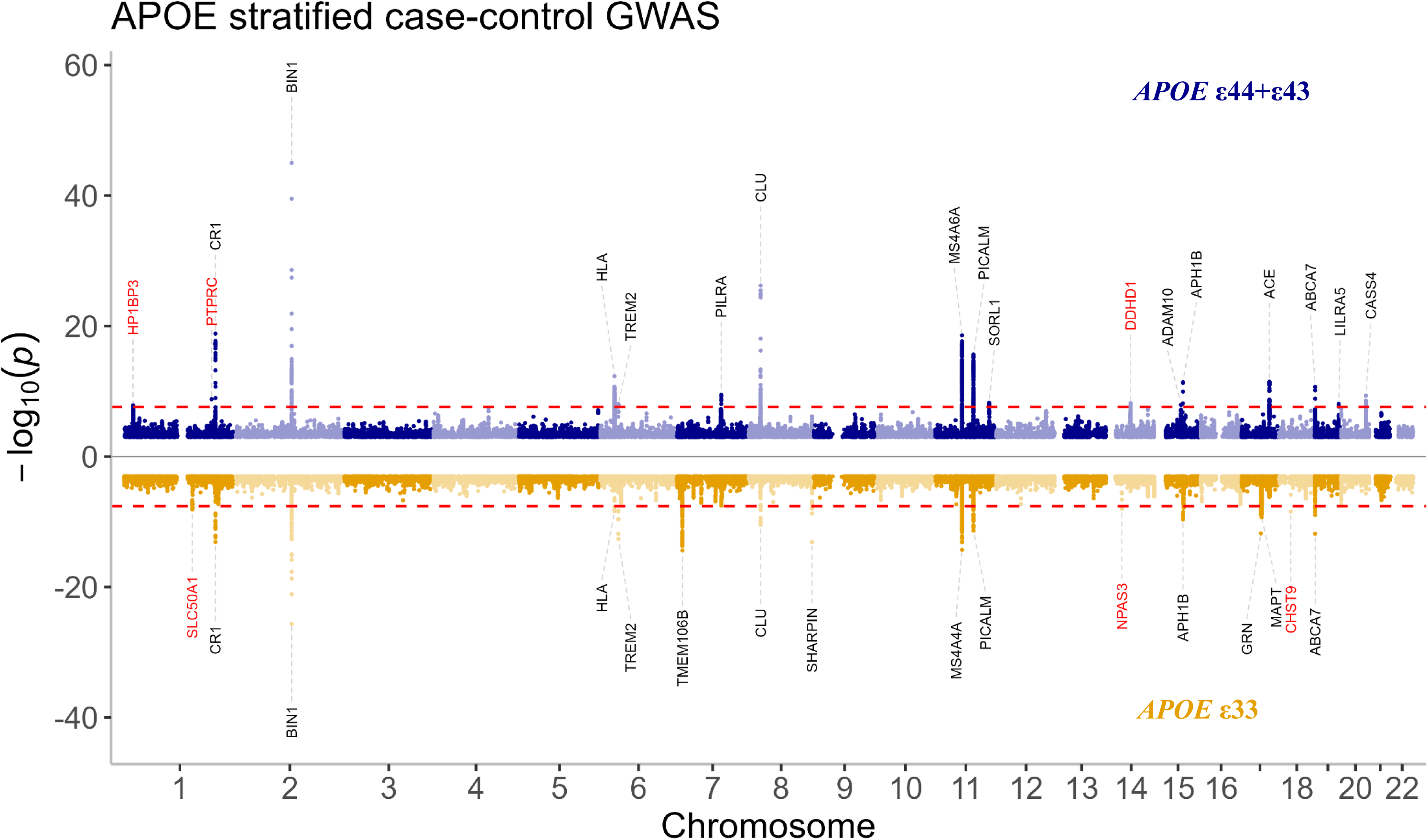
Miami plot of the *APOE* ε33 and ε44+ε43 strata. The Manhattan plot for the *APOE* ε44+ε43 strata is shown in blue in the upper part of the figure and for the *APOE* ε33 strata in orange in the lower part of the figure. Genome wide significant loci are annotated with the nearest gene (known loci in black and new in red). Two-sided raw P-values were derived from a fixed-effect meta-analysis. The red dashed lines show the genome-wide significant level or two GWAS studies (P=2.5×10^-8^). *APOE*: Apolipoprotein E gene.

**Table 1:**
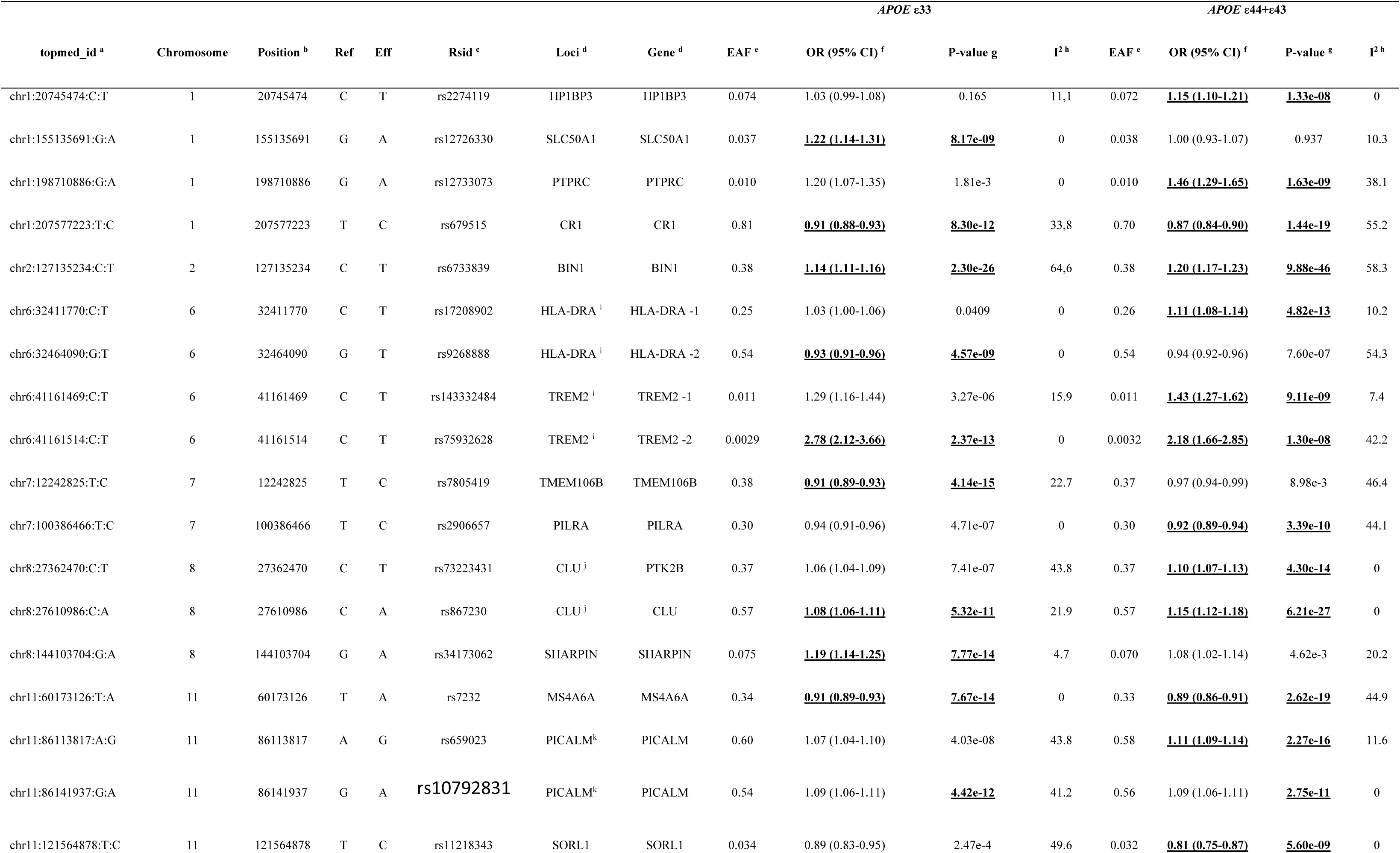
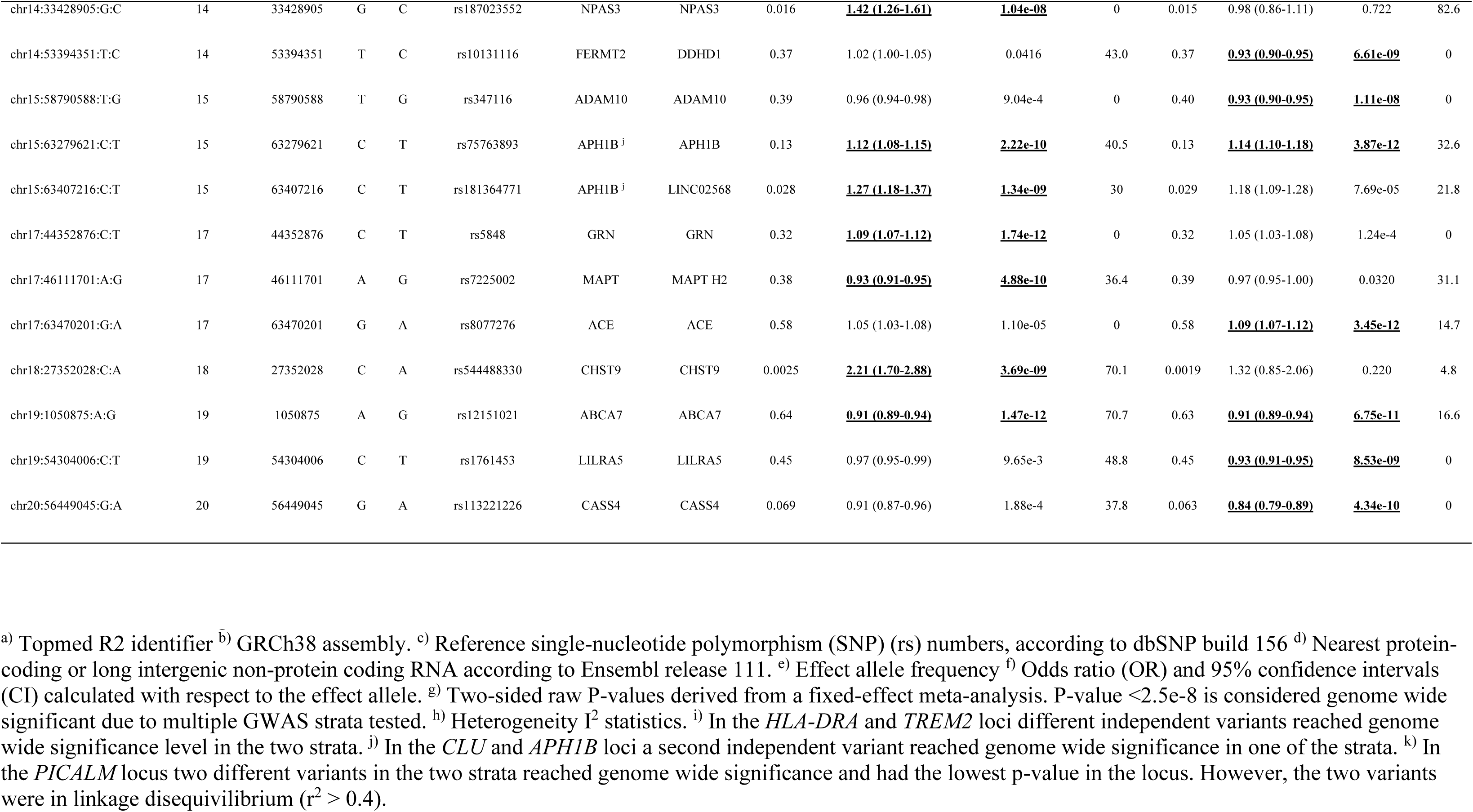
Genome wide significant hits. ^a)^ Topmed R2 identifier ^b)^ GRCh38 assembly. ^c)^ Reference single-nucleotide polymorphism (SNP) (rs) numbers, according to dbSNP build 156 ^d)^ Nearest protein-coding or long intergenic non-protein coding RNA according to Ensembl release 111. ^e)^ Effect allele frequency ^f)^ Odds ratio (OR) and 95% confidence intervals (CI) calculated with respect to the effect allele. ^g)^ Two-sided raw P-values derived from a fixed-effect meta-analysis. P-value <2.5e-8 is considered genome wide significant due to multiple GWAS strata tested. ^h)^ Heterogeneity I^2^ statistics. ^i)^ In the *HLA-DRA* and *TREM2* loci different independent variants reached genome wide significance level in the two strata. ^j)^ In the *CLU* and *APH1B* loci a second independent variant reached genome wide significance in one of the strata. ^k)^ In the *PICALM* locus two different variants in the two strata reached genome wide significance and had the lowest p-value in the locus. However, the two variants were in linkage disequivilibrium (r^2^ > 0.4).

Of note, we also performed a meta-analysis restricted to the ε44 carriers. Only one well-established locus (*BIN1*) was observed in the ε44 stratum including 5,814 AD cases, 14,415 controls and 9,723,486 variants (Supplementary Table 6, Supplementary Figure 1 and Supplementary Figure 2). Forest plots across cohorts and strata are shown in Supplementary Figures 25-53. In addition, we applied clumping procedures and conditional testing to define potential independent signals within each locus detected in the ε33 and ε44+ε43 strata. This approach detected 4 loci presenting two independent signals (*HLA-DRA*, *TREM2*, *CLU*, *APH1B*, Supplementary Table 7), details are specified in legend to Table 1.

The present meta-analyses do not allow us to fully determine whether there is a significant difference between the signals observed in the ε33 and ε44+ε43 strata due to sample and statistical power variations. To formally test for SNP x *APOE* interactions, we employed two complementary approaches. First, we performed interaction tests based on stratified effect comparisons using additive and dominant models with summary statistics from the different *APOE* strata, testing only variants that were significant in one or both strata. Second, we conducted a genome-wide SNP x *APOE* ε4 carrier interaction GWAS using a formal interaction term in a unified model.

In the stratified effect comparison testing dominant *APOE* ε4 effects (meta-analysis of differences between ε33 and ε44+ε43 in each cohort), we found 7 significant stratified effect differences, 3 signals where the effect sizes were attenuated with the presence of an ε4 allele (*SLC50A1*, *TMEM106B*, *NPAS3*) (Table 2, Figure 3) and 3 signals where the effect sizes were augmented with the presence of an ε4 allele (*BIN1*, *HLA-DRA -1*, *CLU*, *DDHD1*) (Table 2, Figure 4). Forest plots of the effect differences across cohorts are shown in Supplementary Figures 54-55. In the additive mixed-effect model we additionally identified *SHARPIN* as interacting with *APOE* ε4, where the effect size was attenuated with an increasing number of ε4 alleles (Table 2, Figure 3). Stratified effect difference sensitivity analyses are shown in Supplementary Table 8.

**Figure 3:**
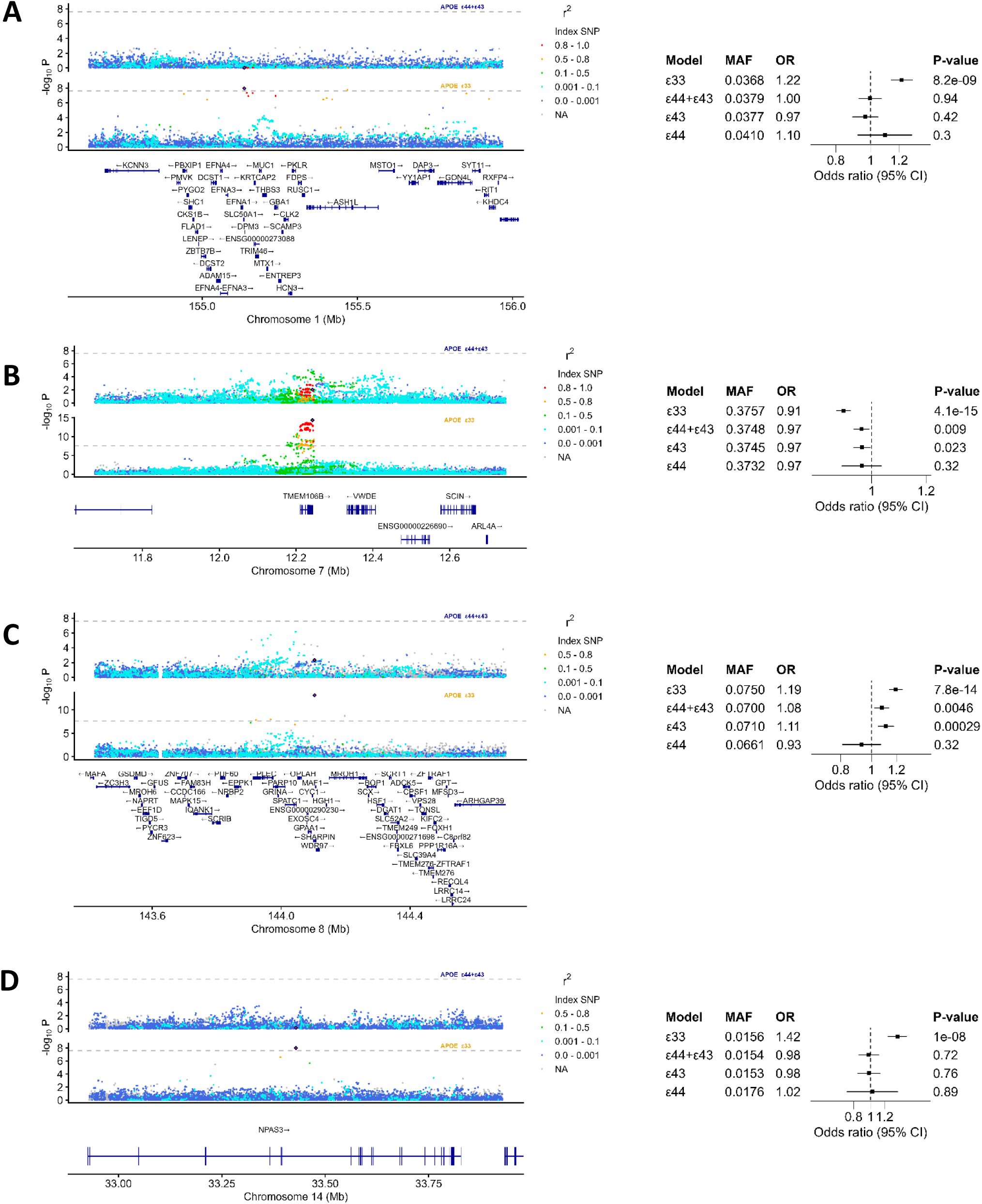
Loci and forest plots for *SLC50A1*, *TMEM106B*, *SHARPIN* and *NPAS3* where the effect attenuates with the *APOE* ε4 allele. Loci plots for *SLC50A1 (*A), *TMEM106B (*B), *SHARPIN (*C*), NPAS3 (*D), in *APOE* ε33 and *APOE* ε44+ε43 strata. Forest plots for the lead SNPs in the four *APOE* strata ε33, ε44+ε43, ε43 and ε44. In the forest plot each *APOE* strata is shown to visualize potential dominant or additive interaction.

**Figure 4:**
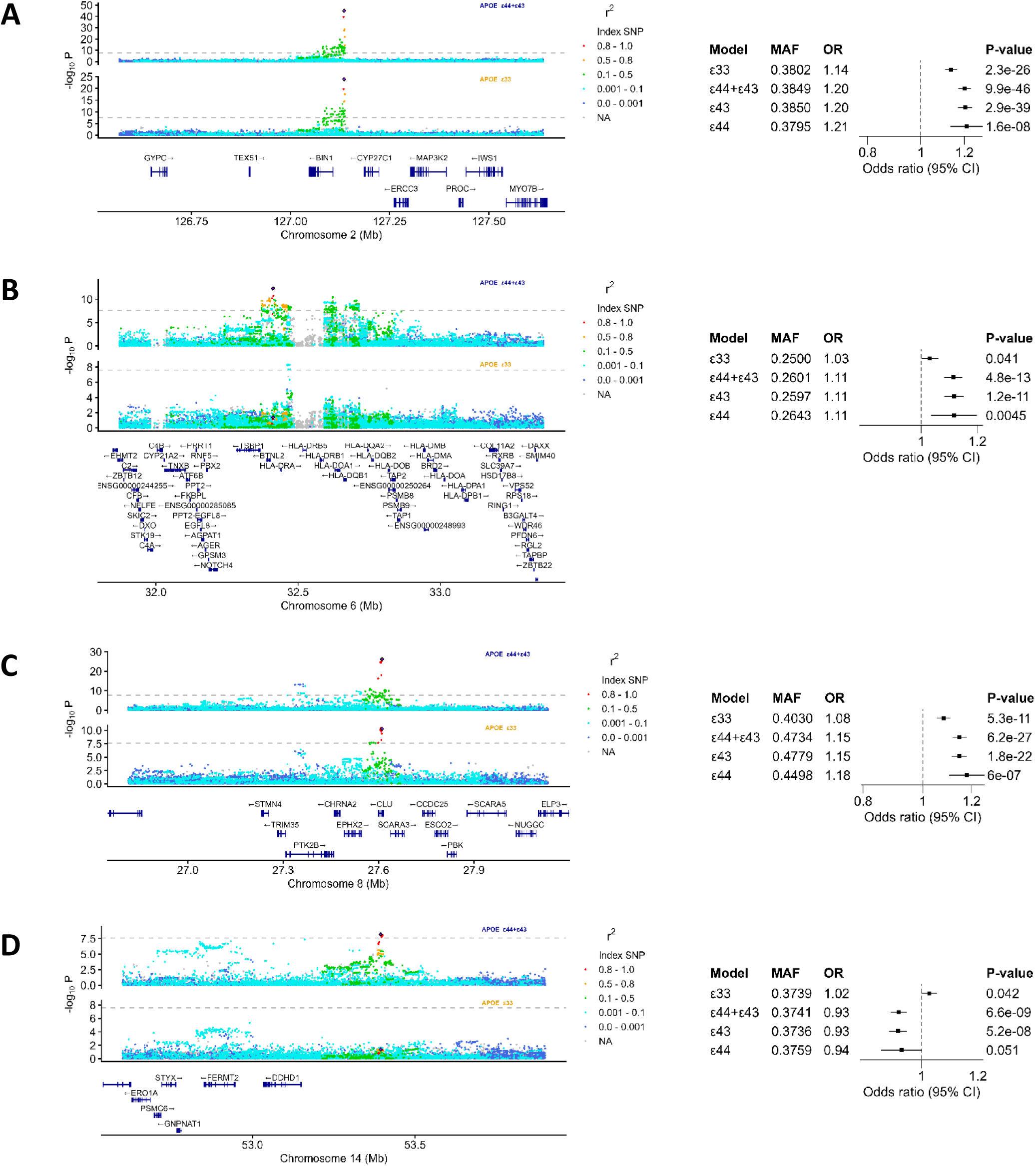
Loci and forest plots for the loci (*BIN1, HLA-DRA -1*, *CLU*, *DDHD1)* where the effect is augmented with the *APOE* ε4 allele. Loci plots for *BIN1* (A), *HLA-DRA -1 (B)*, *CLU (C)*, *DDHD1 (D)* in *APOE* ε33 and *APOE* ε44+ε43 strata. Forest plots for the lead SNPs in the four *APOE* strata ε33, ε44+ε43, ε43 and ε44. In the forest plot each *APOE* strata is shown to visualize potential dominant or additive interaction.

**Table 2:** Interaction testing results. Results of interaction testing with a dominant and an additive fixed effect model on the summary data of the individual cohorts. The dominant model was a difference of effect analysis while the additive model included the number of ε4-allelles. Significant interactions (p-value < 0.0017) are in bold with the significant p-values underlined. ^a)^ Topmed R2 identifier ^b)^ Reference single-nucleotide polymorphism (SNP) (rs) numbers, according to dbSNP build 156 ^c)^ Nearest protein-coding or long intereneric non-coding RNA according to Ensembl release 111. ^d)^ ΔBeta is calculated as β_43+44_- β_33_. ^e)^ Two-sided raw P- values were derived from a fixed-effect meta-analysis. ^f)^ I^2^: I-statistics, residual heterogeneity of the unaccounted variability ^g)^ Het P-value: Test for residual heterogeneity. ^h)^ effect per one ε4-allele.

| topmed_id <sup>a)</sup> | rsid <sup>b)</sup> | Loci | Gene <sup>c)</sup> | Dominant interaction – effect difference |  |  |  |  | Additive interaction – per ε4-allele |  |  |  |  |
| --- | --- | --- | --- | --- | --- | --- | --- | --- | --- | --- | --- | --- | --- |
|  |  |  |  | ΔBeta <sup>d)</sup> | SE | P-value <sup>e)</sup> | I<sup>2</sup> <sup>f)</sup> | Het p-value <sup>g)</sup> | Beta <sup>h)</sup> | SE | P-value | I<sup>2</sup> <sup>f)</sup> | Het p-value <sup>g)</sup> |
| chr1:20745474:C:T | rs2274119 | HP1BP3 | HP1BP3 | 0.092 | 0.034 | 7.2e-3 | 0 | 0.97 | 0.062 | 0.027 | 0.025 | 23 | 0.18 |
| **chr1:155135691:G:A** | **rs12726330** | **SLC50A1** | **SLC50A1** | **-0.19** | **0.051** | **<u>1.6e-4</u>** | 0 | 0.80 | **-0.13** | **0.040** | **<u>0.0013</u>** | 35 | 0.098 |
| chr1:198710886:G:A | rs12733073 | PTPRC | PTPRC | 0.20 | 0.089 | 0.026 | 19 | 0.28 | 0.13 | 0.073 | 0.086 | 0.58 | 0.44 |
| chr1:207577223:T:C | rs679515 | CR1 | CR1 | -0.045 | 0.022 | 0.038 | 46 | 0.072 | -0.021 | 0.017 | 0.226 | 52 | 4.1e-3 |
| **chr2:127135234:C:T** | **rs6733839** | **BIN1** | **BIN1** | **<u>0.064</u>** | **<u>0.018</u>** | **<u>3.7e-4</u>** | 0 | 0.73 | 0.045 | 0.014 | 1.5e-3 | 58 | **<u>6.5e-4</u>** |
| **chr6:32411770:C:T** | **rs17208902** | **HLA-DRA** | **HLA-DRA -1** | **<u>0.078</u>** | **<u>0.020</u>** | **<u>1.1e-4</u>** | 0 | 0.59 | **<u>0.057</u>** | **<u>0.016</u>** | **<u>3.6e-4</u>** | 0 | 0.60 |
| chr6:32464090:G:T | rs9268888 | HLA-DRA | HLA-DRA -2 | 0.0089 | 0.018 | 0.61 | 0 | 0.55 | -0.0027 | 0.014 | 0.85 | 24 | 0.16 |
| chr6:41161469:C:T | rs143332484 | TREM2 | TREM2 -1 | 0.098 | 0.085 | 0.25 | 0 | 0.74 | 0.087 | 0.070 | 0.22 | 7.5 | 0.37 |
| chr6:41161514:C:T | rs75932628 | TREM2 | TREM2 -2 | -0.29 | 0.20 | 0.16 | 8.3 | 0.36 | -0.14 | 0.22 | 0.51 | 0 | 0.53 |
| **chr7:12242825:T:C** | **rs7805419** | **TMEM106B** | **TMEM106B** | **<u>0.065</u>** | **<u>0.018</u>** | **<u>3.1e-4</u>** | 27 | 0.21 | **<u>0.046</u>** | **<u>0.014</u>** | **<u>1.1e-3</u>** | 49 | 7.9e-3 |
| chr7:100386466:T:C | rs2906657 | PILRA | PILRA | -0.015 | 0.019 | 0.43 | 0 | 0.88 | -0.014 | 0.015 | 0.34 | 40 | 0.036 |
| chr8:27362470:C:T | rs73223431 | CLU | PTK2B | 0.036 | 0.018 | 0.042 | 0 | 0.58 | 0.041 | 0.014 | 0.0042 | 0.17 | 0.46 |
| **chr8:27610986:C:A** | **rs867230** | **CLU** | **CLU** | **<u>0.060</u>** | **<u>0.018</u>** | **<u>8.0e-4</u>** | 0 | 0.99 | **<u>0.051</u>** | **<u>0.014</u>** | **<u>2.8e-4</u>** | 0 | 0.49 |
| **chr8:144103704:G:A** | **rs34173062** | **SHARPIN** | **SHARPIN** | -0.10 | 0.037 | 4.9e-3 | 47 | 0.080 | **<u>-0.098</u>** | **<u>0.029</u>** | **<u>7.8e-4</u>** | 15 | 0.29 |
| chr11:60173126:T:A | rs7232 | MS4A6A | MS4A6A | -0.030 | 0.019 | 0.11 | 44 | 0.087 | -0.027 | 0.015 | 0.071 | 6.7 | 0.37 |
| chr11:86113817:A:G | rs659023 | PICALM | PICALM | 0.045 | 0.018 | 0.015 | 13. | 0.33 | 0.039 | 0.015 | 0.0071 | 46 | 0.020 |
| chr11:121564878:T:C | rs11218343 | SORL1 | SORL1 | -0.079 | 0.050 | 0.12 | 33. | 0.16 | -0.096 | 0.041 | 0.019 | 18 | 0.25 |
| **chr14:33428905:G:C** | **rs187023552** | **NPAS3** | **NPAS3** | **<u>-0.37</u>** | **<u>0.091</u>** | **<u>4.2e-05</u>** | 46 | 0.17 | **<u>-0.26</u>** | **<u>0.073</u>** | **<u>0.00032</u>** | 65 | 0.034 |
| **chr14:53394351:T:C** | **rs10131116** | **FERMT2** | **DDHD1** | **<u>-0.10</u>** | **<u>0.018</u>** | **<u>1.1e-08</u>** | 18. | 0.29 | **<u>-0.069</u>** | **<u>0.014</u>** | **<u>1.2e-06</u>** | 32 | 0.088 |
| chr15:58790588:T:G | rs347116 | ADAM10 | ADAM10 | -0.034 | 0.018 | 0.068 | 0 | 0.74 | -0.023 | 0.015 | 0.11 | 0 | 0.62 |
| chr15:63279621:C:T | rs75763893 | APH1B | APH1B | 0.017 | 0.026 | 0.51 | 0 | 0.64 | 0.017 | 0.021 | 0.41 | 12 | 0.30 |
| chr15:63407216:C:T | rs181364771 | APH1B | LINC02568 | -0.082 | 0.058 | 0.16 | 21 | 0.27 | -0.067 | 0.047 | 0.15 | 30. | 0.12 |
| chr17:44352876:C:T | rs5848 | GRN | GRN | -0.041 | 0.019 | 0.030 | 0 | 0.86 | -0.034 | 0.015 | 0.021 | 0 | 0.98 |

**Table 2: Interaction testing results**
| topmed_id <sup>a)</sup> | rsid <sup>b)</sup> | Loci | Gene <sup>c)</sup> | Dominant interaction – effect difference | | | | | Additive interaction – per $\epsilon 4$-allele | | | | |
| --- | --- | --- | --- | --- | --- | --- | --- | --- | --- | --- | --- | --- | --- |
| | | | | $\Delta\text{Beta}$ <sup>d)</sup> | SE | P-value <sup>e)</sup> | I<sup>2</sup> <sup>f)</sup> | Het p-value <sup>g)</sup> | Beta <sup>h)</sup> | SE | P-value | I<sup>2</sup> <sup>f)</sup> | Het p-value <sup>g)</sup> |
| chr17:46111701:A:G | rs7225002 | MAPT | MAPT H2 | 0.045 | 0.018 | 0.012 | 64 | 0.0075 | 0.032 | 0.014 | 0.025 | 34 | 0.066 |
| chr17:63470201:G:A | rs8077276 | ACE | ACE | 0.031 | 0.018 | 0.088 | 0 | 0.82 | 0.028 | 0.014 | 0.048 | 0 | 0.82 |
| chr19:1050875:A:G | rs12151021 | ABCA7 | ABCA7 | -0.012 | 0.019 | 0.52 | 0 | 0.89 | 0.012 | 0.015 | 0.43 | 55 | **<u>1.6e-3</u>** |
| chr19:54304006:C:T | rs1761453 | LILRA5 | LILRA5 | -0.036 | 0.018 | 0.046 | 0 | 0.61 | -0.032 | 0.014 | 0.023 | 0 | 0.62 |
| chr20:56449045:G:A | rs113221226 | CASS4 | CASS4 | -0.098 | 0.039 | 0.011 | 3.7 | 0.40 | -0.058 | 0.031 | 0.061 | 2.9 | 0.42 |

In the genome-wide SNP x *APOE* ε4 carrier interaction GWAS, two new loci (*SMYD2* and *ANK3*) reached genome-wide significance in the SNP term (comparable to *APOE* ε33 strata) (Supplementary Figure 56, Supplementary Table 9. The *APOE* ε4 carrier term was highly significant (p<1×10^-300^) and SNP independent. In the interaction term (SNP x *APOE* ε4 carrier) only the *APOE* locus was significant. However, the *DDHD1* locus reached an interaction p-value of 1.62×10^-6^. The loci with an interaction p-value below 1×10^-5^ are shown in Supplementary Table 10 and the results from the interaction GWAS for the 29 regions of interest are shown in Supplementary Table 11.

We evaluated the 29 regions of interest in additional cohorts of East Asian (EAS) ancestry, representing Japanese (JADNI, CL, NP; EAS-JPN), Chinese (HKS; EAS-CHN), and Korean (GARD; EAS-KOR) populations, as well as in Asian American (ADSP-AAC), African American (ADSP-AFR), and in admixed American (ADSP-AMR) multi-ancestry populations (Supplementary Table 12). Despite several limitations, i.e. difference in linkage structure between cohorts of different ancestries, limitation in statistical power and lead variants being different from the causal variants, similar signals could be observed for several variants (Supplementary Figures 57-83). The meta-analyzed results for *HLA-DRA-1* and *DDHD1* were similar in the East Asian cohorts compared with the European cohorts (Figure 5). A summary of the main interaction results are presented in Figure 6.

**Figure 5:**
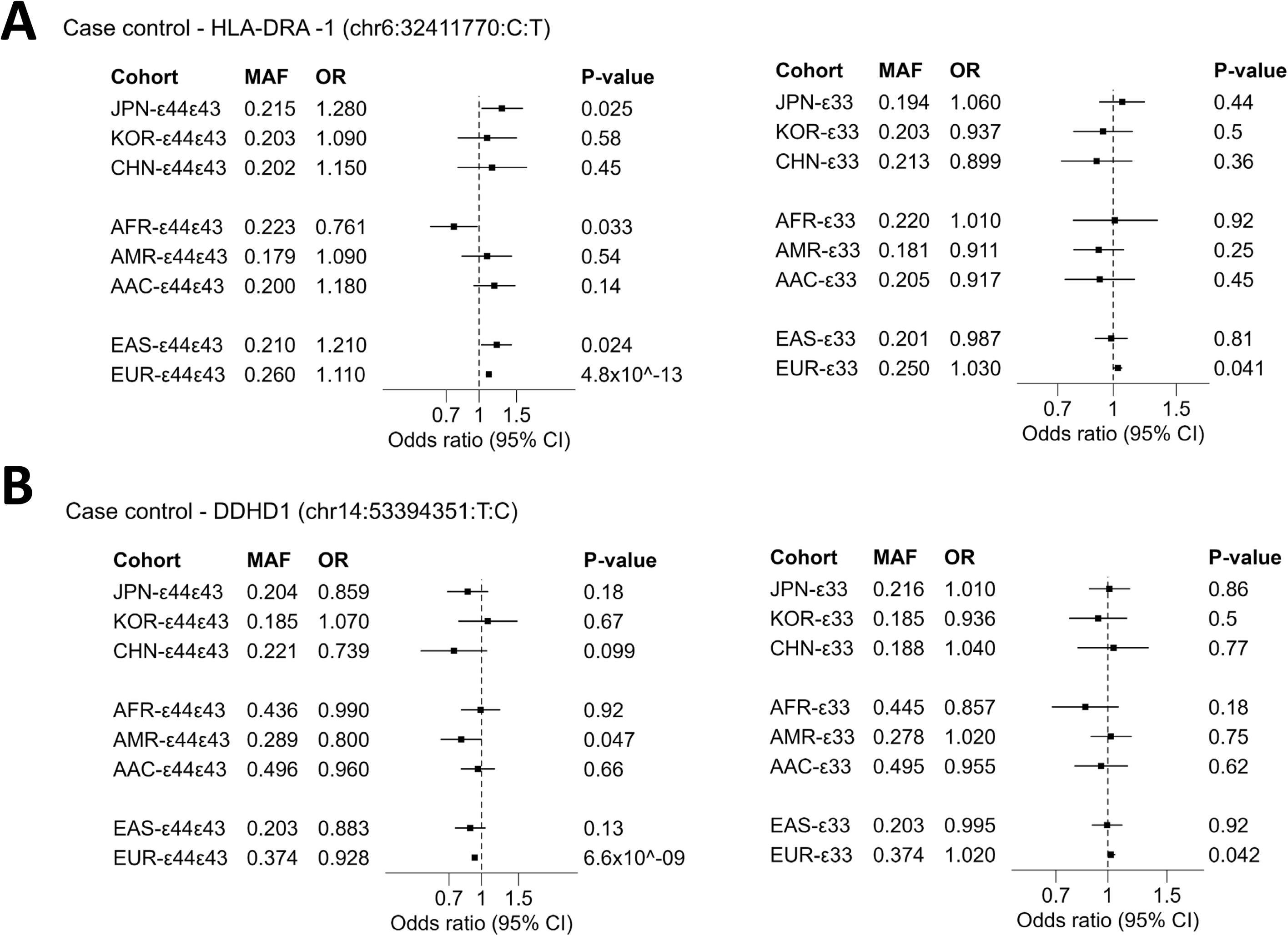
Multi-ancestry evaluation. A: Multi-ancestry results for lead variant in *HLA-DRA* locus. B: Multi-ancestry results for lead variant in *DDHD1* locus. AAC: Asian American, AFR: African American, AMR: Admixed American, CHN: Hong-Kong Chinese, JPN: Japanese, KOR: Korean, EAS: East Asian ancestry meta-analysis, EUR: European ancestry meta-analysis.

**Figure 6:**
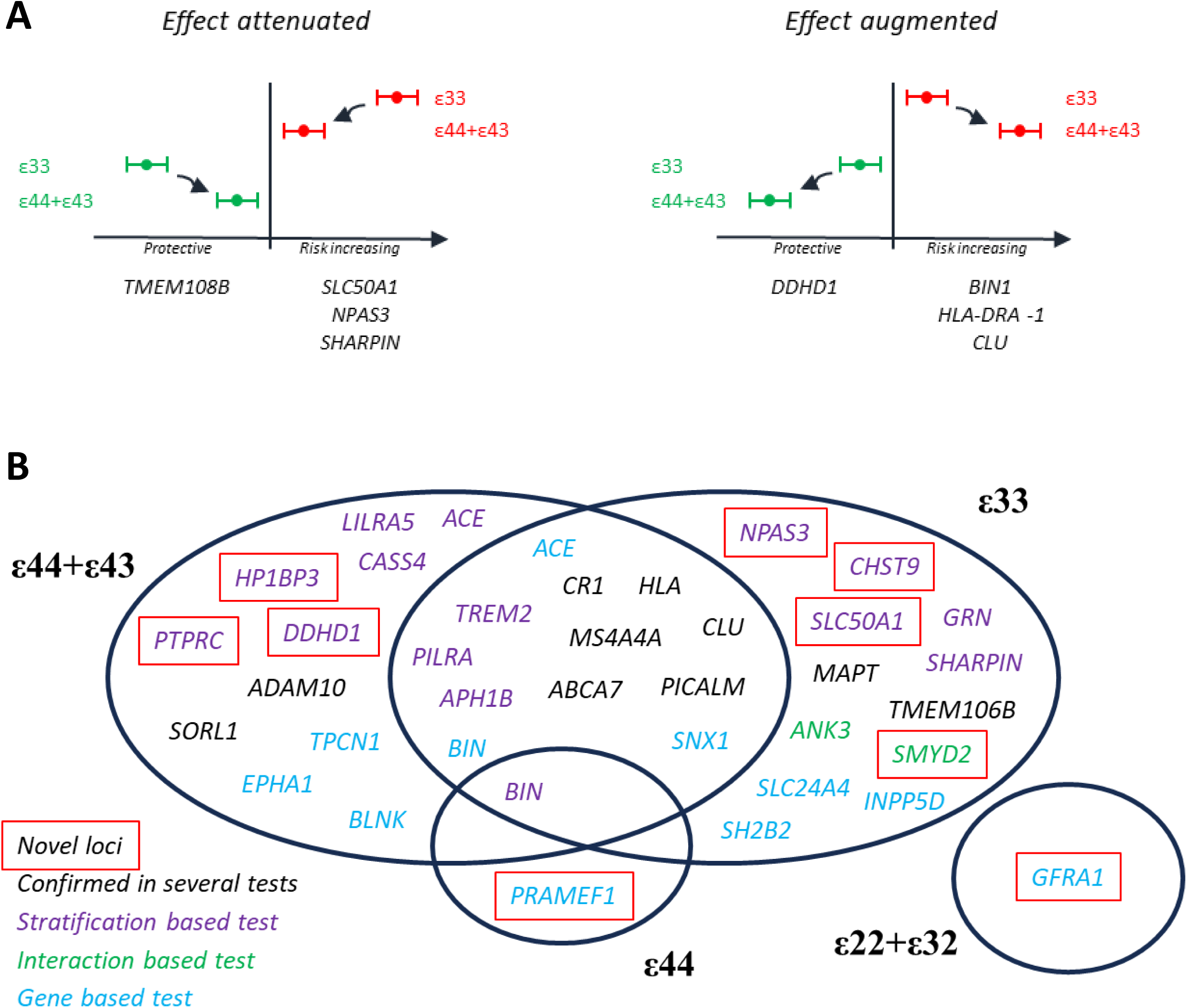
Summary of findings. A) Summary of interaction findings. Left is illustrating attenuated protective and attenuated risk increasing interaction effects for the *APOE*-ε4 allele. Right is illustrating the similar augmented protective and augmented risk increasing interaction effects for the *APOE*-ε4 allele. Loci exhibiting the illustrated effect are noted below. The vertical line illustrates an odds ratio of 1 (no effect). B) Venn-diagram of the genome-wide significant loci associated with AD, showing the overlap between *APOE-*strata and between variant based testing (stratified or interaction GWAS) and gene-based testing (MAGMA).

For lead variant rs10131116 in *DDHD1* a significant eQTL associated with decreased *DDHD1* expression was observed in the ROSMAP dorsolateral prefrontal cortex (β=-0.098, eQTL p=1.67×10^-5^, n=560)^6^ and in the GTEx (v10) brain-putamen (basal-ganglia) (β=-0.23, eQTL p=3.6×10^-5^, n=253)^14^. We tested whether rs10131116 was associated with lower *DDHD1* expression according to *APOE* strata, and nominal p-values were 9×10^-4^ for ε33 and 7×10^-3^ for ε44+ε43 (Supplementary Table 13, Supplementary Figure 84). Further, a gene-based analysis confirmed several of the significant loci from the main stratified GWAS analysis (Figure 6). Also, in the ε44+ε43 stratum the known AD genes *EPHA1*, *TCPN1* and *BLNK* and in the ε33 stratum the *SLC24A4*, *INPP5D* and *SH2B2* (novel but close to the known loci *SPDYE3/PILRA/TMEM225B*) reached significance. *ACE* and *SNX1* were significant in both the ε44+ε43 and ε33 strata, whereas *PRAMEF1* was significant in the ε44 stratum and *GFRA1* in the ε22 stratum (Figure 6, Supplementary Tables 14-17). Next, we performed a pathway enrichment analysis on the *APOE* stratified GWAS results from the eight European studies (Supplementary Tables 18 and 19). After correction for multiple testing in each stratum (q<0.05), 12 and 33 pathways reached statistical significance for the *APOE* ε33 and ε44+ε43 strata, respectively. Overall, pathways related to the complement and immune systems were overrepresented in the *APOE* ε44+ε44 stratum compared to the ε33 stratum, whereas amyloid and neurofibrillary tangle biology was highlighted in both strata. No pathway analysis reached statistical significance for the ε22+ε32 nor for the ε43 stratum. Lastly, we performed a Summary-data-based Mendelian Randomization to test for potential effects of expression on AD that are shared by a causal variant for both *APOE* ε4 carriers (ε44+ε43) and non-carriers (ε33). Four genes passed our significance threshold and HEIDI filtering: *STAG3* (*PILRA* locus) in the ε44+ε43 stratum (Cortex) and *LRRC37A*, *ARL17B* and *LRRC37A2* (*MAPT* locus) in the ε33 stratum (multiple brain regions) (Supplementary Table 20).

## Discussion

By conducting a comprehensive series of *APOE* stratified GWAS analyses, we identified a number of biologically plausible genomic signals that modify the effect of the strongest genetic AD risk variant to date - the *APOE* ε4 allele. As these findings suggest differential biological pathways dependent on the *APOE* strata, they may provide inspiration to how we use genetics in designing randomized clinical trials of future AD medicines.

### New genetic signals

*HP1BP3*, *SLC50A1*, *PTPRC*, *NPAS3*, *DDHD1*, and *CHST9* from the stratified variant based GWAS analysis, *SMYD2* from the interaction GWAS and *PRAMEF1* and *GFRA1* from the gene-based analysis are novel genomic signals for AD risk, only appearing when stratified by *APOE* carrier status, and supported by significant stratified effect comparison tests for *SLC50A1*, *NPASS*, and *DDHD1*, discussed in detail in paragraphs below. *HP1BP3* encodes heterochromatin protein 1 binding protein 3 and is a regulator of cell cycle progression^15^. *PTPRC* encodes protein tyrosine phosphatase receptor type C also known as CD45 that increasingly is understood to play a role in the innate immune system^16,17^. C*HST9* encodes carbohydrate sulfotransferase 9, an enzyme that transfers sulphate to the 4-position of GalNAc. GalNAc4ST-1 and -2 transcripts are highly expressed in the pituitary gland and trachea^19,20^. *SMYD2* encodes a Protein-lysine N-methyltransferase that methylates both histones and non-histone-proteins^21^. *PRAMEF1* encodes a protein **PRAME family member 1** and has been associated with cancer^22^. *GFRA1* encodes for GDNF family receptor alpha-1 which is a receptor for both Glial cell line-derived neurotrophic factor (GDNF) and neurturin (NTN); both potent neurotrophic factors and key regulators of neuron survival and differentiation^23^. GDNF family receptor alpha-1 has been linked to the restoration of AD neuron survival^24^.

### Genetic variants interacting significantly with APOE carrier status with attenuated effect size in ε4 carriers

*SLC50A1* is a novel AD signal that only emerges in the *APOE* ε33 stratum, and encodes solute carrier family 50 member 1, which is a sugar transporter for intercellular exchange and nutrition of pathogens^25^. The previously identified signal, *TMEM106B* ^6^ encodes the lysosomal type II transmembrane protein 106B, and residues of the protein have recently been shown to be amyloidogenic in an age dependent manner and in several neurodegenerative diseases including AD^26^. The presently identified lead hit is in the regulatory 3’UTR part of the gene and is in full LD (r^2^ =0.99) with the previously reported rs1990622 variant – a variant that is associated with reduced expression of *TMEM106B*^27,28^ and with earlier age-at-onset of frontotemporal lobar degeneration in *GRN* mutation carriers. Both the *TMEM106B* and *GRN* signals are sufficiently strong to reach genome-wide significance level in our previous overall GWAS^6^, however the present *APOE* stratified analyses illustrate that these signals only manifest in the *APOE* ε33 context, although only *TMEM106B* reached statistical significance in the stratified effect comparison test.

Interestingly, *TMEM106B* and *GRN* were recently associated only with non-AD pathology in a comprehensive GWAS of multiple neuropathology endophenotypes of dementia^29^. Further aspects of pathophysiology are discussed in the Supplementary Note. *NPAS3* encodes a neuronal transcription factor implicated in several neuropsychiatric conditions^30^ and is reported to have a regulatory function on the expression of reelin^31^. In adults, reelin binds to the ApoE-Receptor2 (apoER2) and the very low-density lipoprotein receptor (VLDLR) modulating AMPA and NMDA activity in the post-synaptic region, affecting APP processing and tau hyperphosphorylation, and competes with apoE in receptor binding^32^. Further, *SHARPIN* variants have been shown to affect NF-kB signalling in the nervous system, a central mediator of inflammatory and immune responses, and apoE is suggested to interact with NF-kB signalling^33,34^. Additionally, we observed consistent directionality in all European cohorts, even though the interaction test did not reach statistical significance, suggesting that the effect of *MAPT* is attenuated in an ε4 context in agreement with a previous report^35^. Finally, Forest plots of the effect differences for the significant stratified effect comparison tests show similar directionality across cohorts.

### Genetic variants interacting significantly with APOE carrier status with augmented effect size in ε4 carriers

The *HLA* region on chromosome 6 is highly complex. The present data add an extra layer to this complexity since we observed that one *HLA* locus associates with increased risk of AD in ε4 carriers, but not in ε3 carriers, while another independent *HLA* locus associates with increased risk of AD in both strata. Further, in the present study we confirmed *CLU* as one of the strongest genomic signals for AD, and documented for the first time that this signal was substantially stronger in *APOE* ε4 carriers compared to ε33 carriers. We also identified a new independent signal within the *FERMT2* locus, where the nearest gene is *DDHD1* (distance 241,028 bp) which encodes a member of the Phospholipase A1 family important in lipid and phospholipid metabolism^23^. The fact that the rs10131116 *DDHD1* variant in the present GWAS was associated with a decreased risk of AD specifically in *APOE* ε4 carriers together with the recent identification of the rs10131116 as an eQTL associated with decreased *DDHD1* gene expression^6,36^, highlights *DDHD1* as an interesting focus for drug discovery. *DDHD1* is also in the same biological pathway as a genome-wide significant known signal (*PLCG2*)^6^. Recently *DDHD2* – a gene known to be involved in hereditary spastic paraplegia - was reported to provide flux of saturated fatty acids for neuronal energy and function^37^, supporting a potential importance of this class of genes in neuronal functioning. Importantly, the *HLA* and *DDHD1* signals were similar in European and Asian cohorts, despite differences in statistical power. Finally, Forest plots of the effect differences also here show similar directionality across cohorts.

## Conclusion

By performing the to date largest *APOE* stratified GWAS, we have identified novel as well as well-established AD loci, where the effect is manifested specifically in an *APOE ε*4 carrier or in an ε33 genotype context. These findings are supported by pathway analysis, highlighting distinct *APOE* carrier status dependent biological mechanisms, and may have the potential to change our current understanding of the pathogenesis of AD. Based on our present findings and based on the observed less efficacy and higher risks of severe side effects in ε4 carriers in recent trials of anti-amyloid therapies, it may now be timely to consider including *APOE* stratification a priori in future randomized clinical trials of emerging AD medicines.

## Methods

### Populations

We used the following European ancestry consortia/biobanks: European Alzheimer’s Disease & Dementia BioBank (EADB), FinnGen, European Alzheimer’s Disease Initiative (EADI), Bonn, Genome Research at Fundacio ACE (Gr@CE), Genetic and Environmental Risk in Alzheimer’s Disease (GERAD), Alzheimer’s Disease Sequencing Project (ADSP), and UK Biobank (UKB). Additionally, we evaluated 33 regions of interest in cohorts of East Asian (EAS) ancestry, representing Japanese (JADNI, CL, NP; EAS-JPN), Chinese (HKS; EAS-CHN), and Korean (GARD; EAS-KOR) populations, as well as in Asian American (ADSP-AAC), African American (ADSP-AFR), and in native admixed American (ADSP-AMR) multi-ancestry populations. *APOE* genotype was determined by the imputed data using rs7412 and rs429358. Where directly genotyped data was available, samples with a mismatch between the imputed and genotyped *APOE* genotype were excluded. FinnGen R11 was used, excluding samples from the ADGEN study as they are embedded in the EADB. AD cases were defined by diagnosis or by use of AD medication (ATC code: N06D). Individuals diagnosed with other forms of dementia were excluded from the controls, and all individuals with an AD diagnosis were included independent of other dementia diagnoses. In UKB only those defined as white British were included, and AD was defined as being diagnosed with AD from electronic medical records (EMR). No information regarding proxies was used in the AD definition or as exclusion criteria. In all datasets, controls younger than 60 years were excluded to better balance the age distributions in the cases and controls and to avoid the inclusion of young controls. All cases were kept. Written informed consent was obtained from study participants or, for those with substantial cognitive impairment, a caregiver, legal guardian, or other proxy. Study protocols for all cohorts were reviewed and approved by the appropriate institutional review boards.

### Quality control and imputation

A standard quality control was performed on the samples and variants in all datasets. The samples were imputed using the TOPMed except for ADSP and FinnGen (Supplementary Table 2). FinnGen was imputed with a Finnish whole-genome sequencing (WGS) reference panel (SiSu v4). Ancestry estimates and QC for UKB and ADSP were done using GenoTools^38^

### GWAS analysis

Test of the association between clinical AD status and autosomal genetic variants were conducted separately in each cohort by means of logistic regression or mixed models using an additive genetic model. Three software implementations were used, SNPTEST 2.5.6^39^, PLINK2^40^ and REGENIE^41^ and adjusted for sex, age, genomic principle components (gPCs) and genotyping centers/batches when necessary (Supplementary Table 2). gPCs were calculated in the full cohort and not in the individual strata to avoid generating possible ascertainment bias effects. Sensitivity analyses were carried out in some datasets to check that adjusting for age did not introduce spurious findings. In SNPTEST we analyzed the genotype probabilities using the newml method. In PLINK2 and REGENIE dosages were used combined with the glm regression (Firth regression if failed convergence) in PLINK2 and Firth regression in REGENIE. For each dataset we filtered out variants with (a) missing data on the effect size, standard error or p-value, (b) an absolute effect above 5, (c) an imputation quality below 0.3, and (d) variants not fulfilling 2×min(N_cases_,N_controls_)×MAF×info > 5 (an unbalanced MAC-info score), where info is imputation quality. A fixed-effect meta-analysis using an inverse-variance weighted as implemented in METAL v2020-05-05 was performed combining the results from each dataset. Variants were excluded if a heterogeneity p-value was below 5×10^-8^ or if variants did not pass quality control in at least two of the three major datasets (EADB-TOPMed, FinnGen, UKB). The genomic inflation factor was computed with a median approach after exclusion of the *APOE* region (44-46 Mb on chromosome 19 in GRCh38) both for variants with MAF>1% and in the entire dataset. Manhattan plots were made using topr package^42^ (v.2.0.2) in R (v. 4.3.3).

### Definition of loci

Around each variant with a p-value below 2.5×10^-8^, a region of ±500kb was defined per fixed-effect meta-analysis. We assumed the individual stratified GWASs as separate families of analyses when setting the p-value threshold. Most of the variants will be highly correlated across the strata except for those variants that interact with the *APOE* genotype which is expected to be a small minority of the total tested variants. Hence, if one was to consider all GWAS tests as belonging to the same family it would lead to a large increase in risk of Type II errors without much gain in controlling for Type I errors. However, to be conservative in setting a p-value threshold for the stratified GWAS analysis, we set a stricter Bonferroni correction of *p<5*×10^-8^*/2=2.5*×10^-8^ to take into account the addition of the two main strata (ε33 and ε43+ε44).We used the PLINK2 clumping procedure to define independent hits in each region. This procedure is iterative, starting with the variant with the lowest p-value in the respective region (the lead SNP). All variants within loci and in linkage disequilibrium (LD) with the lead variant (r^2^ higher that 0.01) are assigned to the clump belonging to the lead SNP. If any variant with a p-value below 2.5×10^-8^ is unassigned to a clump in the respective region, the variant with the lowest p-value is found among the remaining variants and the clumping is repeated until all variants have been assigned a clump. LD in the EADB TOPMed imputed dataset was computed using high quality imputed dosages (imputation info>0.8). The clumping procedure was run in both the ε44+ε43 and ε33 GWAS results and the results for the respective loci were compared. Loci plots were generated using locuszoomr (v. 0.3.5) in R (v. 4.3.3). Forest plots of variant effects across *APOE* strata were generated using forestplotter (v. 1.1.2) in R (v. 4.3.3). The independence of several signals within a locus was tested by conditional meta-analysis in the five largest cohorts (EADB TOPMed, FINNGEN, UK Biobank, GR@CE, ADSP). and meta-analyzed.

### Stratified effect comparison tests

Two different tests of stratified effect comparisons between autosomal variants and the *APOE* strata were performed using summary statistics results from the different *APOE* strata. P-value threshold for significance was determined using a Bonferroni correction for 29 regions of interest (including the nine loci in common in both strata, the nine and seven loci found in the ε44+ε43 and ε33 strata respectively and the four additional signals found in four of the 25 loci; p<0.05/29 regions of interest=0.0017). The first test analysed the effect difference between the ε44+ε43 and ε33 *APOE* strata. The effect difference was calculated in each cohort separately (Δ*β*_*i*_ = *β*_*i*,*ε*44+*ε*43_ − *β*_*i*,*ε*33_, *i* is the cohort, *β* is the autosomal effect estimated in the stratified GWASs), with the SE of the effect difference calculated as the square root of sum of squares of SE for the effects 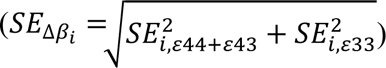. The effect differences were combined across studies in a fixed effect meta-analysis with an inverse-variance weighted approach (METAL v2020-05-05 software). The test was referred to as the dominant test because it tests if the presence of an ε4-allele changes the effect of the autosomal variant (independent of number of ε4-alleles). Forest plots for the calculated effect difference in each cohort was provided to access the robustness of the stratified effect comparison test across the cohorts. The second test was a fixed effect model estimating the effect from the number of ε4 alleles: *β*_*ij*_ = *γ*_0_ + *γ*_*ε*4_*x*_*ij*_ + *γ*_*cohort*_*C*_*ij*_ + *ε*_*ij*_, where *i* is the cohort, *j* is the strata (ε33, ε43, ε44), *β_ij_* is the autosomal effect, *x_ij_* is the number of ε4-alleles (0,1,2), *C_ij_* is the cohort, ɣ_0_ is the intercept, ɣ_ε4_ is the effect of one ε4 allele, ɣ_cohort_ is the cohort effect and *ε_ij_* the error term. The second model was referred to as the additive model and was estimated using R (v.4.3.3) and Metafor package (v4.6.0).

We performed mixed effect sensitivity interaction models to test if the stratified effect comparisons results were prone to between-strata relatedness bias, which could be a potential bias in the FinnGen cohort. The models were specified as following. Dominant mixed effect model: *β*_*ik*_ = *γ*_0_ + *γ*_*k*_*x*_*ik*_ + *u*_*i*_ + *ε*_*ik*_, where *i* is the cohort, *k* is the strata (ε33, ε44+ε43) *x_ik_* is the strata (ε33, ε44+ε43, categorical), ɣ*_k_* is the strata effect (ɣ_ε43+ε44_ is the reference) and *u_i_* is the random effect. Additive mixed model: *β*_*ij*_ = *γ*_0_ + *γ*_*ε*4_*x*_*ij*_ + *u*_*i*_ + *ε*_*ij*_, with the same notation as above. For the calculation of effect difference both the ε33 and ε44+ε43 effects should be available for a cohort to be included in the analysis. In the other models we included all available data, e.g. the ε44 GWAS in the Bonn cohort was not performed due to power, but the data from the Bonn ε33, ε43 and ε44+ε43 GWASs were included if the SNPs passed QC.

### Genome-wide interaction testing

Interaction testing between *APOE* ε4 carrier status and autosomal genetic variants for association with clinical AD was conducted separately in the five largest cohorts (EADB TOPMed, FinnGen, UK Biobank, GR@CE, ADSP) using logistic regression or mixed models with an additive genetic model. We implemented a gene-environment (GxE) interaction framework, treating *APOE* ε4 carrier status as the exposure modifier. The aim of the analysis was to mimic as closely as possible the main analysis contrasting the ε33 and ε44+ε43 strata. A full multi-allelic gene-gene interaction GWAS across all *APOE* genotypes was regarded as outside the scope of this project.

*APOE* ε4 carrier status was defined as 0 for individuals in the *APOE* ε33 strata and 1 for individuals in the *APOE* ε44+ε43 stratum. All other strata were excluded from the analysis. The model used for analysis was AD ∼ SNP + *APOE* ε4 carrier + SNP×*APOE* ε4 carrier + age + sex + cohort-specific covariates. The software and the specific adjustments for the different cohorts is detailed in Supplementary Table 2. Results from the individual cohorts were QCed as described in the stratified GWAS and the SNP and SNP×*APOE*_ε4_carrier estimates were meta-analysed separately using an inverse-variance weighted fixed-effect meta-analysis as implemented in METAL v2020-05-05. The same significance threshold (*p<2.5*×10^-8^) was used in the interaction model as in the stratified GWAS meta-analysis, as SNP and interaction terms constitute independent tests. In this setup the reference level in the interaction model corresponds to the reference level in the *APOE* ε33 strata GWAS and hence 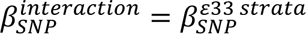. Likewise the estimated effect for a SNP in the *APOE* ε44+ε43 strata corresponds to 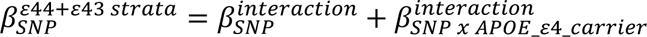 but with the reference value shifted by the *APOE*_ε4_carrier estimate 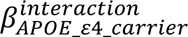, which is independent of the individual SNP.

### Pathway analysis

Pathway analyses were performed on each meta-analysis stratum result separately using FUMA v1.6.1^43^. As requested by FUMA, all variants were annotated with an rsID using VEP release 112 and then lifted to the GRCh37 assembly using Picard LiftoverVcf tool (v3.1.1)^44^. Variants having no rsID or failing the lift to the GRCh37 assembly were removed from the analysis. Remaining variants were then uploaded to FUMA and the pathway analysis was performed by MAGMA v1.08 ^45^ using two different windows to assign a variant to a gene: 0kb (main analysis) and a window of 35kb upstream and 10kb downstream (second analysis). To account for multiple testing, we computed a false discovery rate (FDR, Benjamini-Hochberg) based on the number of genes included in the analysis. Pathways having a q-value<0.05 in either of the two windows were considered significant.

### eQTL analysis

We performed an *APOE* stratified cis-eQTL mapping analysis (*APOE* ε33 (n=342) and *APOE* ε44+ε43 (n=130)) in the ROSMAP dorsolateral prefrontal cortex (DLPFC; n=560) cohort to investigate association of the lead variant in the *DDHD1* locus (*rs10131116*) with the RNA expression of nearby genes in a 1 Mb window around the variant, following the methodology described as before^6^. For the *APOE* stratified cis-eQTL mapping, the genetic principal components (gPCs) and the gene expression Probabilistic Estimation of Expression Residuals (PEER) factors were calculated within the respective strata separately; and sex, first 3 gPCs, and PEER factors (first 45 for *APOE* ε33 and first 15 for *APOE* ε44+ε43) were included as covariates.

### Summary data-based Mendelian Randomization

Summary data-based Mendelian Randomization (SMR) was performed using the SMR software developed and maintained by the Yang Lab, with default parameters^46,47^ using summary data from the *APOE* ε33 and *APOE* ε44+ε43 strata GWAS. We used cis-eQTL data from 5 of the 7 regions in the MetaBrain Consortium dataset (cerebellum, cortex, basal ganglia, hippocampus, and spinal cord)^48^. We applied a significance threshold of pSMR_multi <6.12E-06, which corresponds to the Bonferroni-corrected value at α = 0.05 for 8,166 unique genes tested across all regions.

Additionally, we filtered at a HEIDI p-value threshold of pHEIDI >0.01 to remove associations with inferred pleiotropy and only kept results where the number of SNPs included in the HEIDI tests was greater than 3 (nsnp_HEIDI >3).

## Data availability

Summary statistics will be made available upon publication through the European Bioinformatics Institute GWAS Catalog (https://www.ebi.ac.uk/gwas/).

## Code availability

The software used is referenced in the Online Methods and the applied codes will be made available on https://github.com/JesperQT/APOEstratGWAS.

## Acknowledgements

We thank the many study participants, researchers and staff for collecting and contributing to the data, the high-performance computing service at the University of Lille and the staff at CEA-CNRGH for their help with sample preparation and genotyping and excellent technical assistance. This work was funded by a grant (EADB) from the EU Joint Programme – Neurodegenerative Disease Research. INSERM UMR1167 is also funded by the INSERM, Institut Pasteur de Lille, Lille Métropole Communauté Urbaine and French government’s LABEX DISTALZ program (development of innovative strategies for a transdisciplinary approach to AD). This work was further funded by grants from the Lundbeck Foundation (R278-2018-804), the Danish Heart Foundation, and the Research Fund at Sygeforsikringen Danmark (2021-0245). Najaf Amin is funded by National Institute on Aging (NIH) and Oxford-GSK Institute of Molecular and Computational Medicine (IMCM). Cornelia M van Duijn is supported by the US National Institute on Aging (NIH), NovoNordisk, the Oxford-GSK Institute of Molecular and Computational Medicine (IMCM), Centre of Artificial Intelligence for Precision Medicines (CAIPM) of the University of Oxford and King Abdul Aziz University, Alzheimer Research UK (ARUK), UK National Institute for Health and Care Research (NIHR) Oxford Research Center (BRC), ZonMW (Delta Dementie) and Alzheimer Nederland. This research was also supported in part by the Intramural Research Program of the NIH, National Institute of Aging (NIA), National Institutes of Health, Department of Health and Human Services; project number ZO1 AG000534, as well as the National Institute of Neurological Disorders and Stroke. This work utilized the computational resources of the NIH STRIDES Initiative (https://cloud.hoh.gov) through the Other Transaction agreement – Azure: OT2OD032100, Google Cloud Platform: OT2OD027060, Amazon Web Services: OT2OD027852. This work utilized the computational resources of the NIH HPC Biowulf cluster (https://hpc.noh.gov). Part of this research has been conducted using the UK Biobank Resource under Application Number 33601, and this work uses data provided by patients and collected by the National Health Service as part of their care and support. H Leonard is collaborator on UK Biobank Application Number 33601 and had access to the data under this approved project. Full consortium acknowledgements and funding are in the Supplementary Note.

## Author contributions

Conception and design: JQ Thomassen, H Leonard, B Ulms, B Grenier-Boley, JC Lambert, C van Duijn, M Nalls, R Frikke-Schmidt.

Data analysis: JQ Thomassen, H Leonard, B Ulms, B Grenier-Boley, S Heikkinen, P Garcia, A Castillo-Morales, M Kikuchi, J Gim, H Cao, F Küçükali, N Amin, D Yoon.

Sample contribution: All authors.

Interpretation of data: JQ Thomassen, H Leonard, B Ulms, B Grenier-Boley, F Küçükali, N Amin, JC Lambert, C van Duijn, M Nalls, R Frikke-Schmidt.

Core writing group: JQ Thomassen, H Leonard, B Grenier-Boley, JC Lambert, C van Duijn, M Nalls, R Frikke-Schmidt.

## Additional information

**Correspondence and requests for materials** should be addressed to Jesper Qvist Thomassen, Mike Nalls, and Ruth Frikke-Schmidt.

## References

1. Lambert, J.-C. et al. Meta-analysis of 74,046 individuals identifies 11 new susceptibility loci for Alzheimer’s disease. Nature Genetics 45, 1452–1458 (2013).

2. Jansen, I.E. et al. Genome-wide meta-analysis identifies new loci and functional pathways influencing Alzheimer’s disease risk. Nat Genet 51, 404–413 (2019).

3. Kunkle, B.W. et al. Genetic meta-analysis of diagnosed Alzheimer’s disease identifies new risk loci and implicates Abeta, tau, immunity and lipid processing. Nat Genet 51, 414–430 (2019).

4. Wightman, D.P. et al. A genome-wide association study with 1,126,563 individuals identifies new risk loci for Alzheimer’s disease. Nat Genet 53, 1276–1282 (2021).

5. de Rojas, I., et al. Common variants in Alzheimer’s disease: Novel association of six genetic variants with AD and risk stratification by polygenic risk scores. *medRxiv* (2020).

6. Bellenguez, C. et al. New insights into the genetic etiology of Alzheimer’s disease and related dementias. Nat Genet 54, 412–436 (2022).

7. Lambert, J.C., Ramirez, A., Grenier-Boley, B. & Bellenguez, C. Step by step: towards a better understanding of the genetic architecture of Alzheimer’s disease. Mol Psychiatry 28, 2716–2727 (2023).

8. Corder, E. et al. Gene Dose of Apolipoprotein E type 4 Allele and the Risk of Alzheimer’s Disease in Late Onset Families. Science 261, 921–3 (1992).

9. Lake, J. et al. Multi-ancestry meta-analysis and fine-mapping in Alzheimer’s disease. Mol Psychiatry 28, 3121–3132 (2023).

10. Fernandez-Calle, R. et al. APOE in the bullseye of neurodegenerative diseases: impact of the APOE genotype in Alzheimer’s disease pathology and brain diseases. Mol Neurodegener 17, 62 (2022).

11. Mintun, M.A. et al. Donanemab in Early Alzheimer’s Disease. N Engl J Med 384, 1691–1704 (2021).

12. van Dyck, C.H. et al. Lecanemab in Early Alzheimer’s Disease. N Engl J Med 388, 9–21 (2023).

13. Sperling, R.A. et al. Trial of Solanezumab in Preclinical Alzheimer’s Disease. N Engl J Med 389, 1096–1107 (2023).

14. GTEx Portal (https://www.gtexportal.org/home/snp/rs10131116).

15. Dutta, B. et al. Profiling of the Chromatin-associated Proteome Identifies HP1BP3 as a Novel Regulator of Cell Cycle Progression. Mol Cell Proteomics 13, 2183–97 (2014).

16. Al Barashdi, M.A., Ali, A., McMullin, M.F. & Mills, K. Protein tyrosine phosphatase receptor type C (PTPRC or CD45). J Clin Pathol 74, 548–552 (2021).

17. Kaplan, R. et al. Cloning of Three Human Tyrosine Phosphatases Reveals a Multigene Family of Receptor-Linked Protein-Tyrosine-Phosphatases Expressed in Brain. Proc Natl Acad Sci U S A 87, 7000–7004.

18. Hoeng, J.C. et al. Identification of new human cadherin genes using a combination of protein motif search and gene finding methods. J Mol Biol 337, 307–17 (2004).

19. Kang, H.-G., Evers, M.R., Xia, G., Baenziger, J.U. & Schachner, M. Molecular Cloning and Expression of anN-Acetylgalactosamine-4-O-sulfotransferase That Transfers Sulfate to Terminal and Non-terminal β1,4-LinkedN-Acetylgalactosamine. Journal of Biological Chemistry 276, 10861–10869 (2001).

20. Hiraoka, N., Misra, A., Belot, F., Hindsgaul, O. & Fukuda, M. Molecular cloning and expression of two distinct human N-acetylgalactosamine 4-O-sulfotransferases that transfer sulfate to GalNAcβ1-4GlcNAcβ1-R in both N- and O-glycans. Glycobiology 11, 495–504 (2001).

21. Yi, X., Jiang, X.J. & Fang, Z.M. Histone methyltransferase SMYD2: ubiquitous regulator of disease. Clin Epigenetics 11, 112 (2019).

22. https://www.ncbi.nlm.nih.gov/gene/65121.

23. The Human Protein Atlas (proteinatlas.org).

24. Konishi, Y. et al. Deficiency of GDNF Receptor GFRalpha1 in Alzheimer’s Neurons Results in Neuronal Death. J Neurosci 34, 13127–38 (2014).

25. Chen, L.Q. et al. Sugar transporters for intercellular exchange and nutrition of pathogens. Nature 468, 527–32 (2010).

26. Schweighauser, M. et al. Age-dependent formation of TMEM106B amyloid filaments in human brains. Nature 605, 310–314 (2022).

27. Cruchaga, C. et al. Association of TMEM106B gene polymorphism with age at onset in granulin mutation carriers and plasma granulin protein levels. Arch Neurol 68, 581–6 (2011).

28. Van Deerlin, V.M. et al. Common variants at 7p21 are associated with frontotemporal lobar degeneration with TDP-43 inclusions. Nat Genet 42, 234–9 (2010).

29. Shade, L.M.P. et al. GWAS of multiple neuropathology endophenotypes identifies new risk loci and provides insights into the genetic risk of dementia. Nat Genet 56, 2407–2421 (2024).

30. Michaelson, J.J. et al. Neuronal PAS Domain Proteins 1 and 3 Are Master Regulators of Neuropsychiatric Risk Genes. Biol Psychiatry 82, 213–223 (2017).

31. Erbel-Sieler, C. et al. Behavioral and regulatory abnormalities in mice deficient in the NPAS1 and NPAS3 transcription factors. Proc Natl Acad Sci U S A 101, 13648–53 (2004).

32. Stranahan, A.M., Erion, J.R. & Wosiski-Kuhn, M. Reelin signaling in development, maintenance, and plasticity of neural networks. Ageing Res Rev 12, 815–22 (2013).

33. Asanomi, Y. et al. A rare functional variant of SHARPIN attenuates the inflammatory response and associates with increased risk of late-onset Alzheimer’s disease. Mol Med 25, 20 (2019).

34. Arnaud, L. et al. APOE4 drives inflammation in human astrocytes via TAGLN3 repression and NF-kappaB activation. Cell Rep 40, 111200 (2022).

35. Jun, G. et al. A novel Alzheimer disease locus located near the gene encoding tau protein. Mol Psychiatry 21, 108–17 (2016).

36. <final supplementary data Bellenguez et al Nature Genetics.pdf>.

37. Saber, S.H. et al. DDHD2 provides a flux of saturated fatty acids for neuronal energy and function. Nat Metab 7, 2117–2141 (2025).

38. Vitale, D. et al. GenoTools: an open-source Python package for efficient genotype data quality control and analysis. G3 (Bethesda) 15(2025).

39. Marchini, J., Howie, B., Myers, S., McVean, G. & Donnelly, P. A new multipoint method for genome-wide association studies by imputation of genotypes. Nat Genet 39, 906–13 (2007).

40. Chang, C.C. et al. Second-generation PLINK: rising to the challenge of larger and richer datasets. GigaScience 4(2015).

41. Mbatchou, J. et al. Computationally efficient whole-genome regression for quantitative and binary traits. Nat Genet 53, 1097–1103 (2021).

42. Juliusdottir, T. topr: an R package for viewing and annotating genetic association results. BMC Bioinformatics 24, 268 (2023).

43. Watanabe, K., Taskesen, E., van Bochoven, A. & Posthuma, D. Functional mapping and annotation of genetic associations with FUMA. Nat Commun 8, 1826 (2017).

44. McLaren, W. et al. The Ensembl Variant Effect Predictor. Genome Biol 17, 122 (2016).

45. de Leeuw, C.A., Mooij, J.M., Heskes, T. & Posthuma, D. MAGMA: generalized gene-set analysis of GWAS data. PLoS Comput Biol 11, e1004219 (2015).

46. Wu, Y. et al. Integrative analysis of omics summary data reveals putative mechanisms underlying complex traits. Nat Commun 9, 918 (2018).

47. Zhu, Z. et al. Integration of summary data from GWAS and eQTL studies predicts complex trait gene targets. Nat Genet 48, 481–7 (2016).

48. de Klein, N. et al. Brain expression quantitative trait locus and network analyses reveal downstream effects and putative drivers for brain-related diseases. Nat Genet 55, 377–388 (2023).

