## Supplementary figures for "*APOE* stratified genome-wide association studies provide novel insights into the genetic etiology of Alzheimers’s disease"

### Supplementary Figure 1

ε22+ ε32

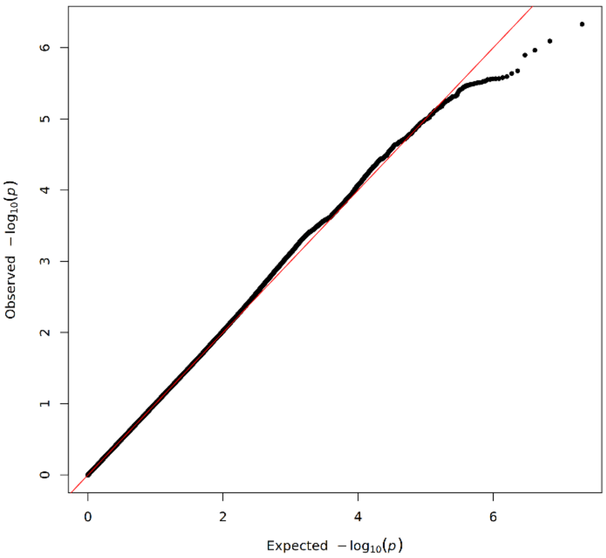


ε33

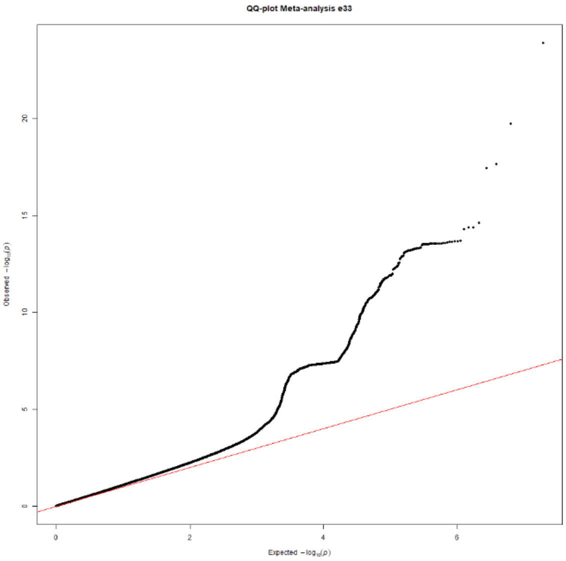


ε44+ε43

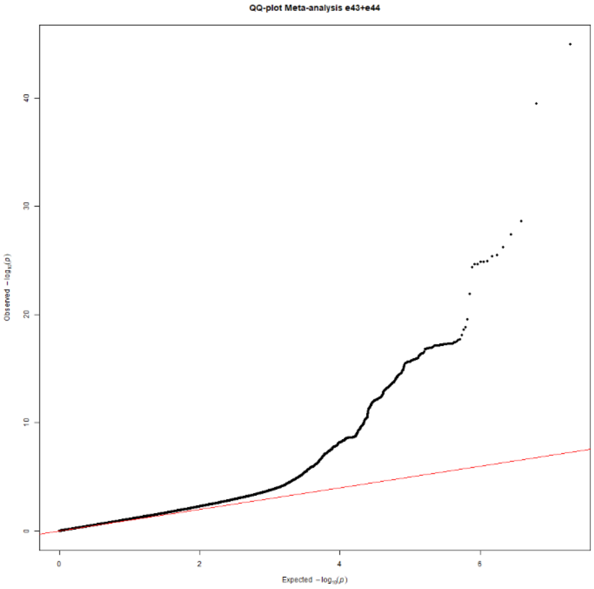


ε44

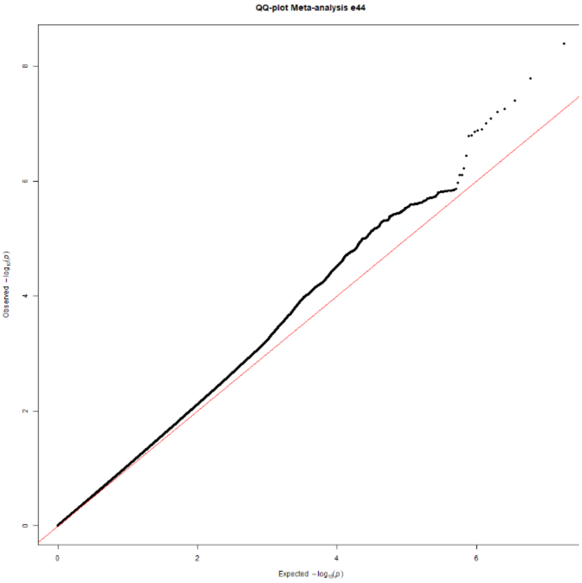


**Supplementary Figure 1: QQ-plots of *APOE* stratified GWAS meta-analysis**

QQ-plots for the respective *APOE* strata for variants MAF>0.01.

**Supplementary Figure 2**

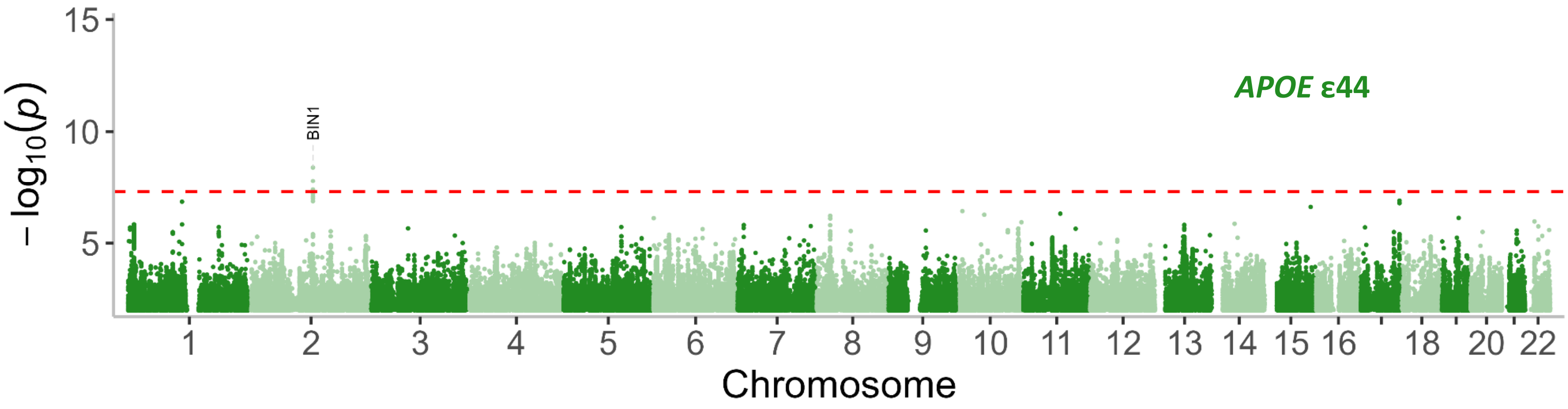


**Supplementary Figure 2: Manhattan plot of the *APOE* ε44 strata.**

Genome wide significant loci are annotated with nearest gene (known loci in black and new in red). Two-sided raw P-values were derived from a fixed-effect meta-analysis. The red dashed line shows the genome-wide significance level ($P=2.5\times10^{-8}$). *APOE*: Apolipoprotein E gene.

**Supplementary Figure 3**

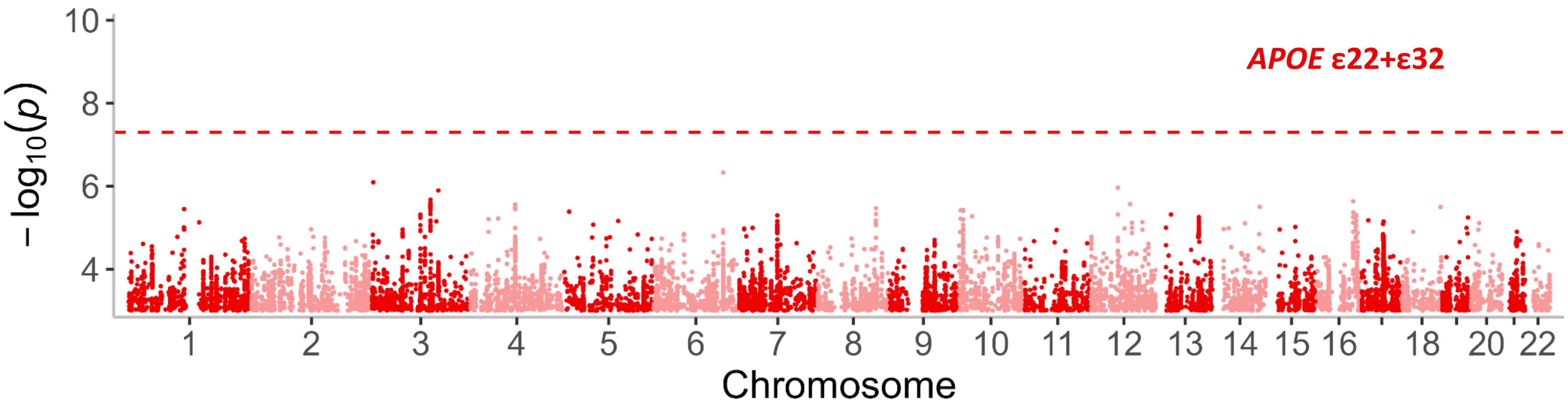


**Supplementary Figure 3: Manhattan plot of the *APOE* ε22+ε32 strata.**

Two-sided raw P-values were derived from a fixed-effect meta-analysis. The red dashed line shows the genome-wide significance level ($P=2.5\times10^{-8}$). *APOE*: Apolipoprotein E gene.

### Supplementary Figure 4

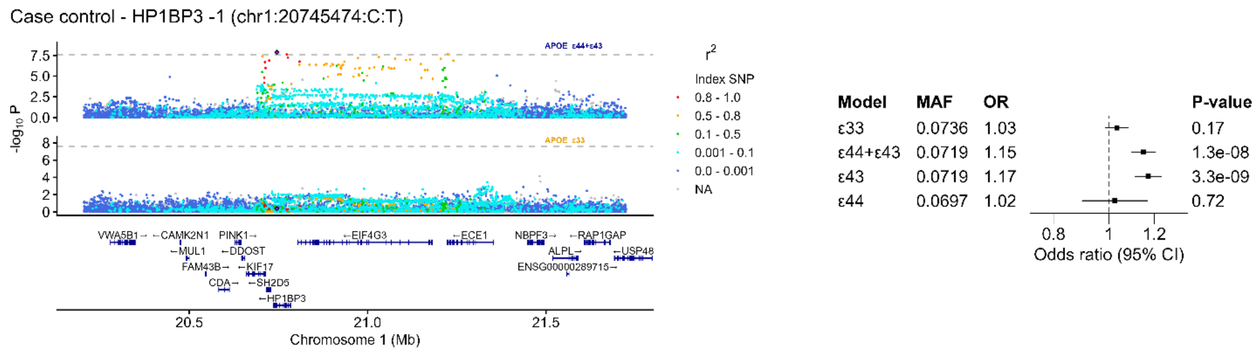


**Supplementary Figure 4: Loci and forest plot for *HP1BP3.***

Loci plots for *HP1BP3* in both the *APOE* ε33 and *APOE* ε44+ε43 strata and forest plots for the lead SNPs for *HP1BP3* in the four *APOE* strata ε33, ε44+ε43, ε43 and ε44. In the forest plot each *APOE* strata is shown to visualize potential dominant or additive interaction.

### Supplementary Figure 5

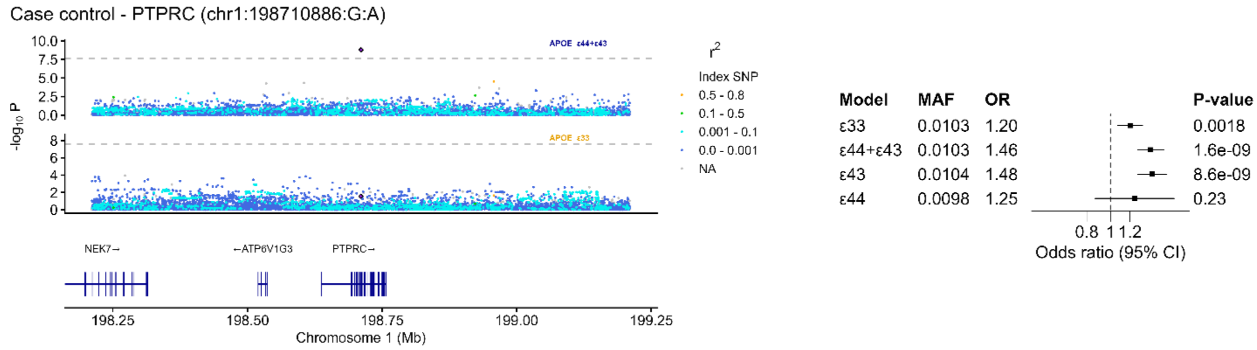


**Supplementary Figure 5: Loci and forest plot for *PTPRC.***

Loci plots for *PTPRC* in both the *APOE* ε33 and *APOE* ε44+ε43 strata and forest plots for the lead SNPs for *PTPRC* in the four *APOE* strata ε33, ε44+ε43, ε43 and ε44. In the forest plot each *APOE* strata is shown to visualize potential dominant or additive interaction.

### Supplementary Figure 6

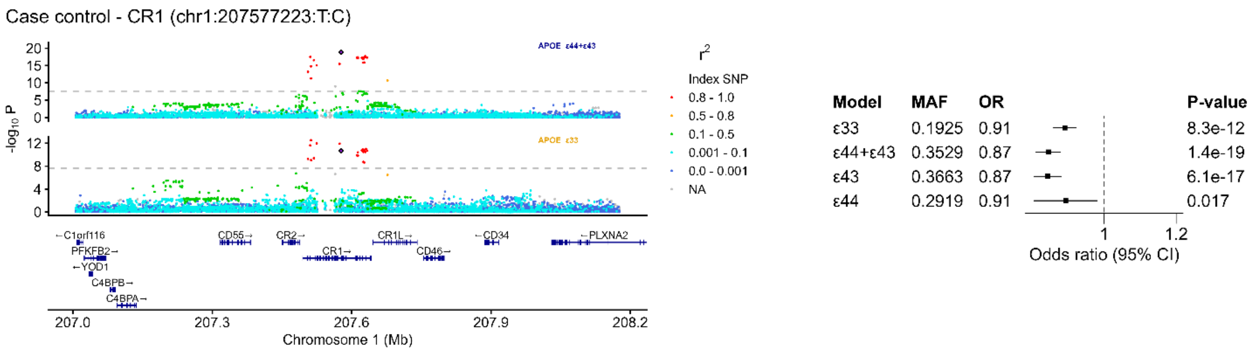


### Supplementary Figure 6: Loci and forest plot for *CR1.*

Loci plots for *CR1* in both the *APOE* ε33 and *APOE* ε44+ε43 strata and forest plots for the lead SNPs for *CR1* in the four *APOE* strata ε33, ε44+ε43, ε43 and ε44. In the forest plot each *APOE* strata is shown to visualize potential dominant or additive interaction.

### Supplementary Figure 7

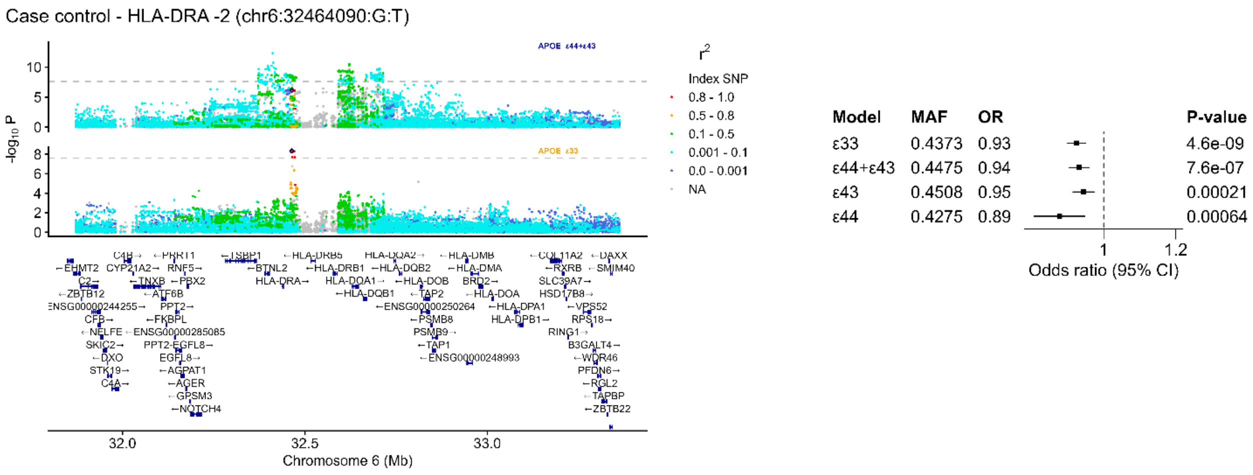


### Supplementary Figure 7: Loci and forest plot for *HLA-DRA -2.*

Loci plots for *HLA-DRA -2* in both the *APOE* ε33 and *APOE* ε44+ε43 strata and forest plots for the lead SNPs for *HLA-DRA -2* in the four *APOE* strata ε33, ε44+ε43, ε43 and ε44. In the forest plot each *APOE* strata is shown to visualize potential dominant or additive interaction.

### Supplementary Figure 8

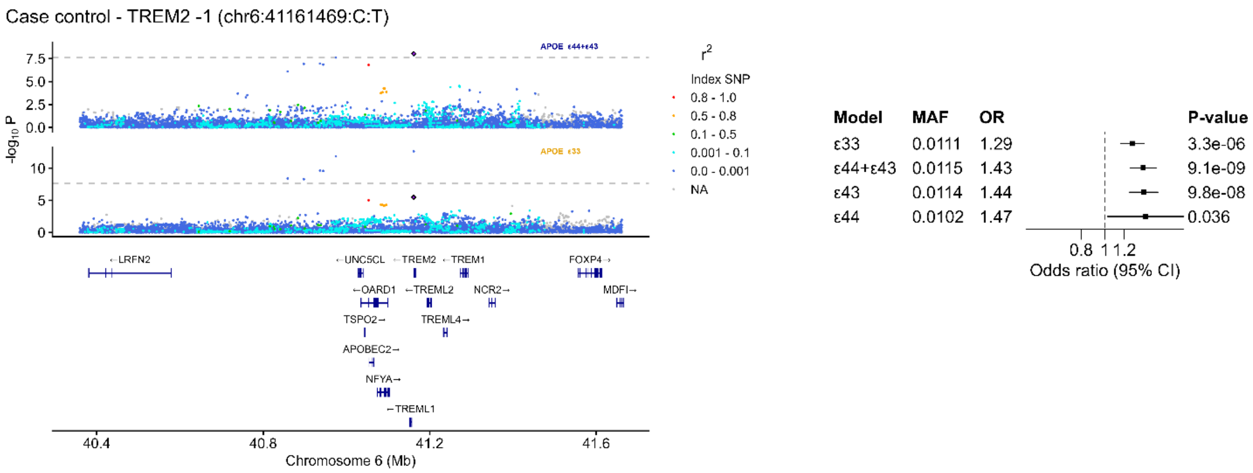


### Supplementary Figure 8: Loci and forest plot for *TREM2 -1*

Loci plots for *TREM2 -1* in both the *APOE* ε33 and *APOE* ε44+ε43 strata and forest plots for the lead SNPs for *TREM2 -1* in the four *APOE* strata ε33, ε44+ε43, ε43 and ε44. In the forest plot each *APOE* strata is shown to visualize potential dominant or additive interaction.

### Supplementary Figure 9

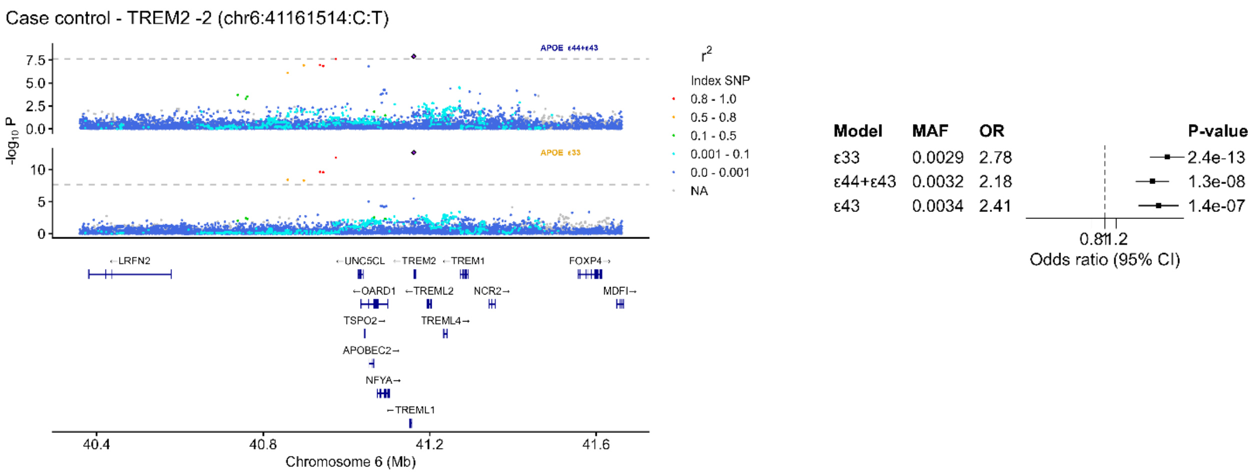


### Supplementary Figure 9: Loci and forest plot for *TREM2 -2*

Loci plots for *TREM2 -2* in both the *APOE* ε33 and *APOE* ε44+ε43 strata and forest plots for the lead SNPs for *TREM2 -2* in the four *APOE* strata ε33, ε44+ε43, ε43 and ε44. In the forest plot each *APOE* strata is shown to visualize potential dominant or additive interaction.

### Supplementary Figure 10

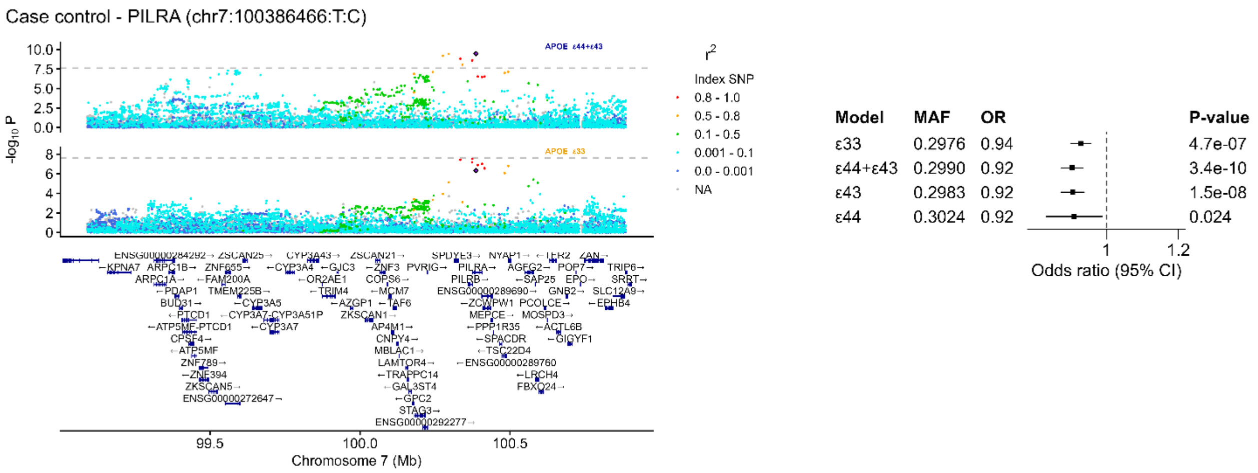


**Supplementary Figure 10: Loci and forest plot for *PILRA.***

Loci plots for *PLIRA* in both the *APOE* ε33 and *APOE* ε44+ε43 strata and forest plots for the lead SNPs for *PLIRA* in the four *APOE* strata ε33, ε44+ε43, ε43 and ε44. In the forest plot each *APOE* strata is shown to visualize potential dominant or additive interaction.

### Supplementary Figure 11

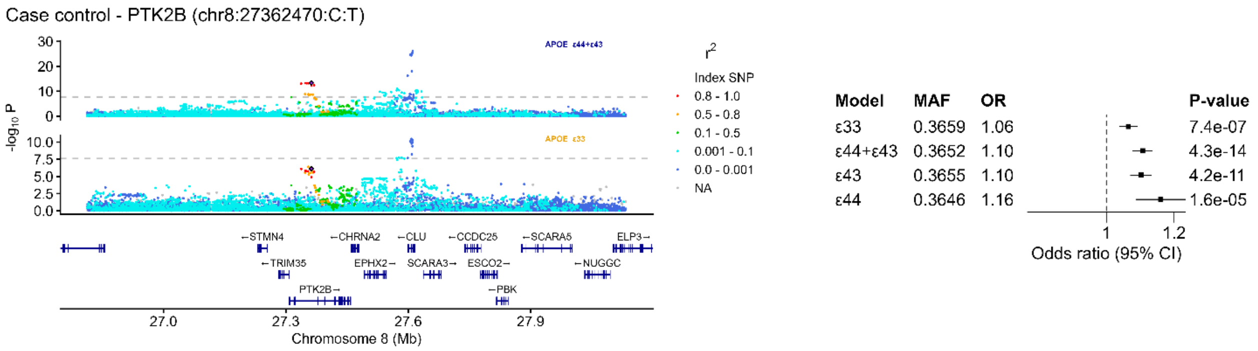


**Supplementary Figure 11: Loci and forest plot for *PTK2B.***

Loci plots for *PTK2B* in both the *APOE* ε33 and *APOE* ε44+ε43 strata and forest plots for the lead SNPs for *PTK2B* in the four *APOE* strata ε33, ε44+ε43, ε43 and ε44. In the forest plot each *APOE* strata is shown to visualize potential dominant or additive interaction.

### Supplementary Figure 12

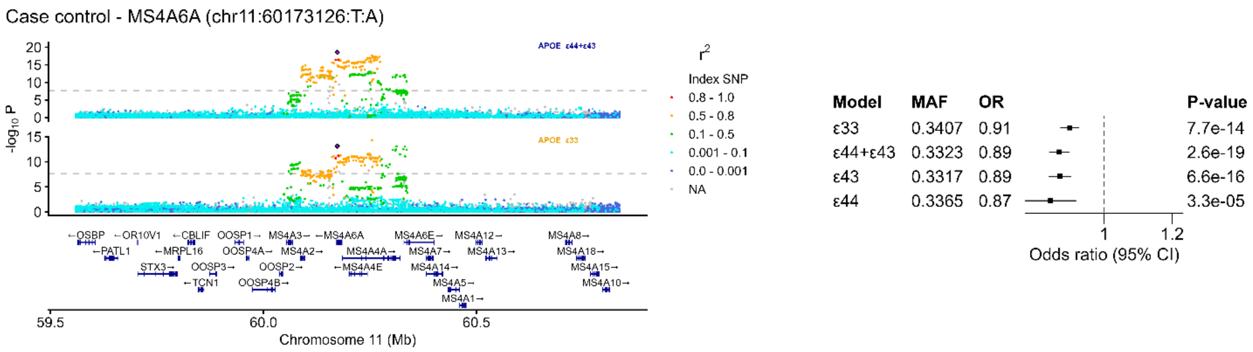


#### Supplementary Figure 12: Loci and forest plot for *MS4A6A.*

Loci plots for *MS4A6A* in both the *APOE* ε33 and *APOE* ε44+ε43 strata and forest plots for the lead SNPs for *MS4A6A* in the four *APOE* strata ε33, ε44+ε43, ε43 and ε44. In the forest plot each *APOE* strata is shown to visualize potential dominant or additive interaction.

### Supplementary Figure 13

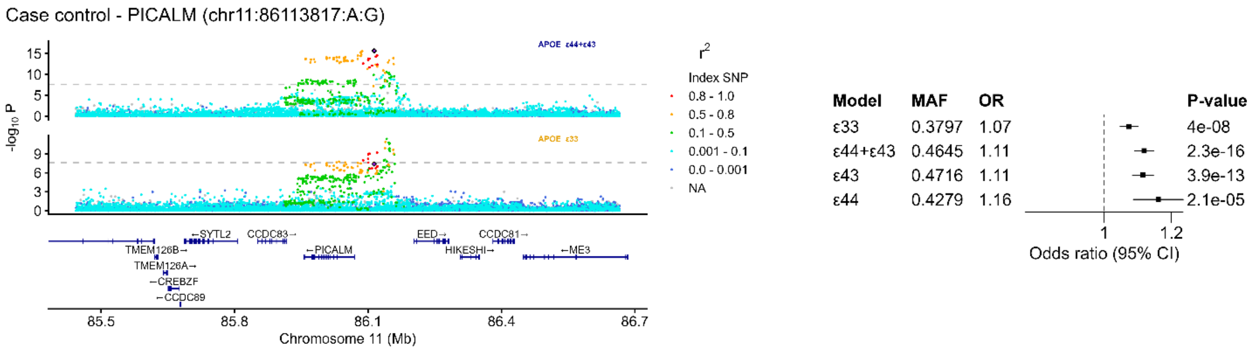


#### Supplementary Figure 13: Loci and forest plot for *PICALM.*

Loci plots for *PICALM* in both the *APOE* ε33 and *APOE* ε44+ε43 strata and forest plots for the lead SNPs for *PICALM* in the four *APOE* strata ε33, ε44+ε43, ε43 and ε44. In the forest plot each *APOE* strata is shown to visualize potential dominant or additive interaction.

### Supplementary Figure 14

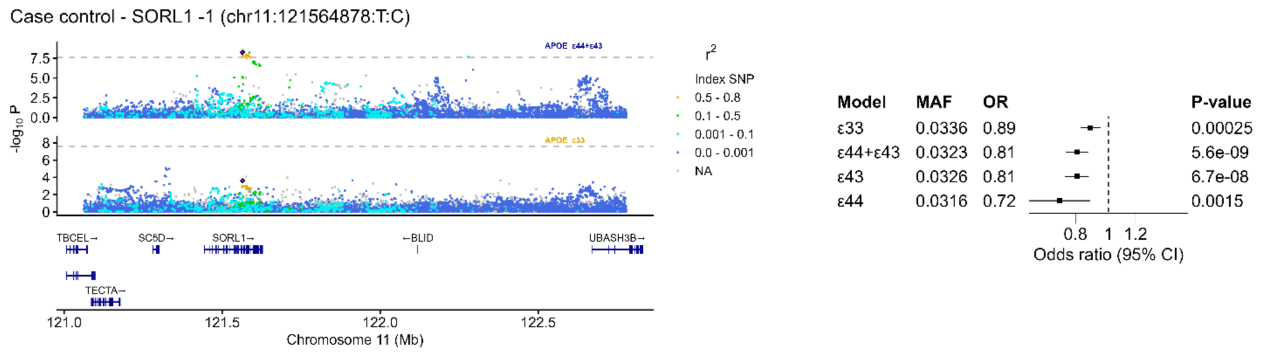


**Supplementary Figure 14: Loci and forest plot for *SORL1*.**

Loci plots for *SORL1* in both the *APOE* ε33 and *APOE* ε44+ε43 strata and forest plots for the lead SNPs for *SORL1* in the four *APOE* strata ε33, ε44+ε43, ε43 and ε44. In the forest plot each *APOE* strata is shown to visualize potential dominant or additive interaction.

### Supplementary Figure 15

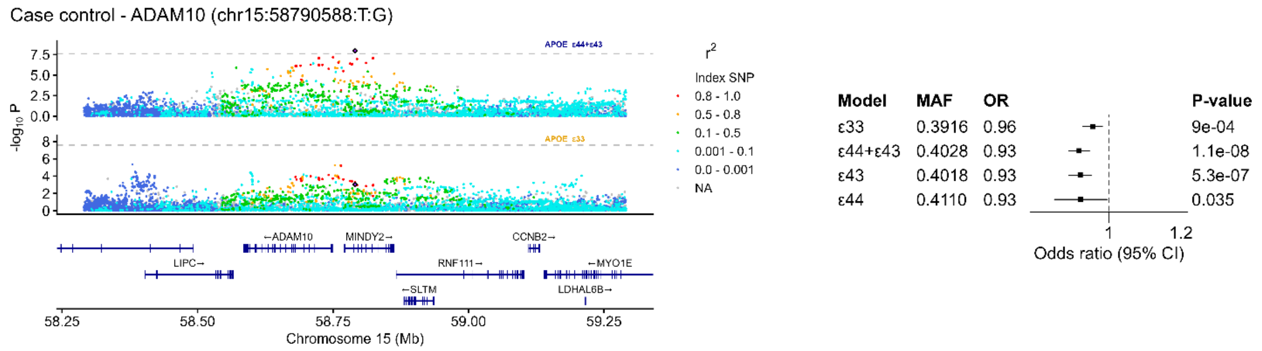


**Supplementary Figure 15: Loci and forest plot for *ADAM10*.**

Loci plots for *ADAM10* in both the *APOE* ε33 and *APOE* ε44+ε43 strata and forest plots for the lead SNPs for *ADAM10* in the four *APOE* strata ε33, ε44+ε43, ε43 and ε44. In the forest plot each *APOE* strata is shown to visualize potential dominant or additive interaction.

### Supplementary Figure 16

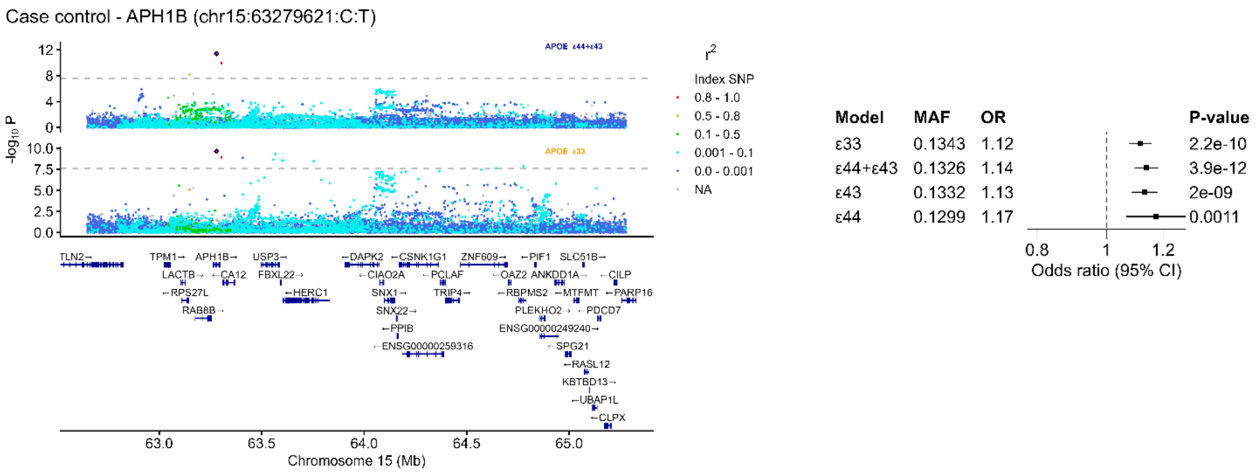


**Supplementary Figure 16: Loci and forest plot for *APH1B*.**

Loci plots for *APH1B* in both the *APOE* ε33 and *APOE* ε44+ε43 strata and forest plots for the lead SNPs for *APH1B* in the four *APOE* strata ε33, ε44+ε43, ε43 and ε44. In the forest plot each *APOE* strata is shown to visualize potential dominant or additive interaction.

### Supplementary Figure 17

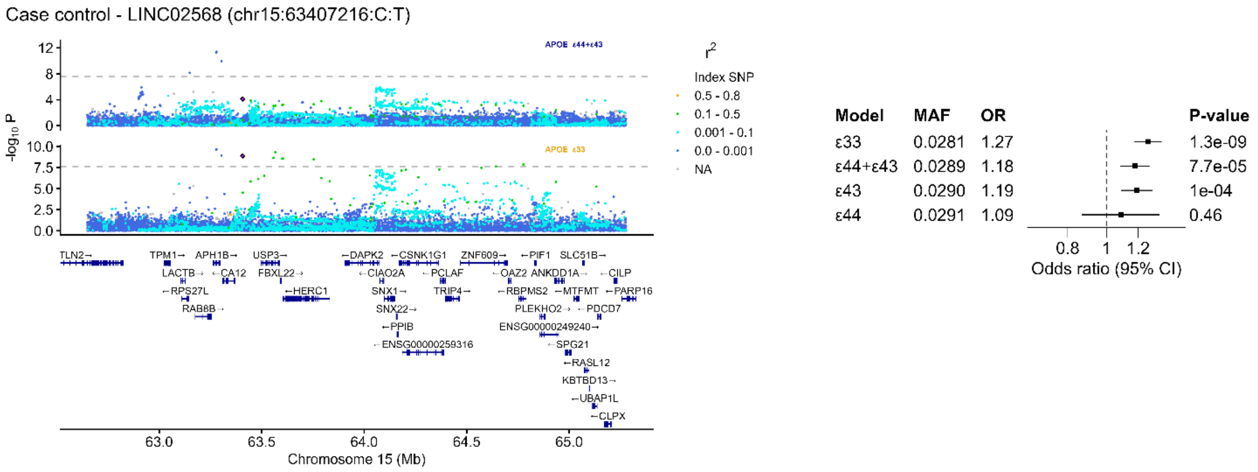


**Supplementary Figure 17: Loci and forest plot for *LINC02568*.**

Loci plots for *LINC02568* in both the *APOE* ε33 and *APOE* ε44+ε43 strata and forest plots for the lead SNPs for *LINC02568* in the four *APOE* strata ε33, ε44+ε43, ε43 and ε44. In the forest plot each *APOE* strata is shown to visualize potential dominant or additive interaction.

### Supplementary Figure 18

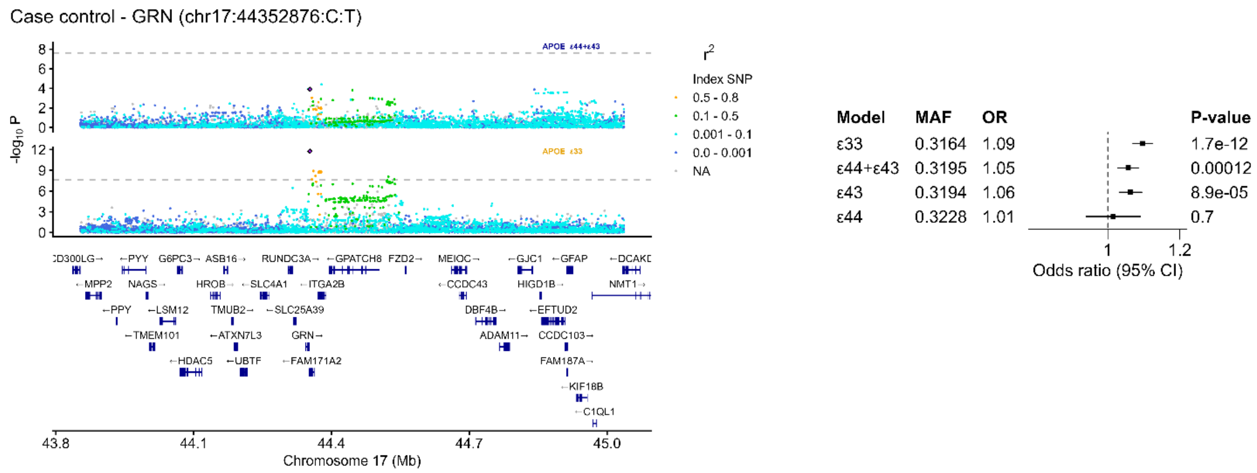


**Supplementary Figure 18: Loci and forest plot for *GRN*.**

Loci plots for *GRN* in both the *APOE* ε33 and *APOE* ε44+ε43 strata and forest plots for the lead SNPs for *GRN* in the four *APOE* strata ε33, ε44+ε43, ε43 and ε44. In the forest plot each *APOE* strata is shown to visualize potential dominant or additive interaction.

### Supplementary Figure 19

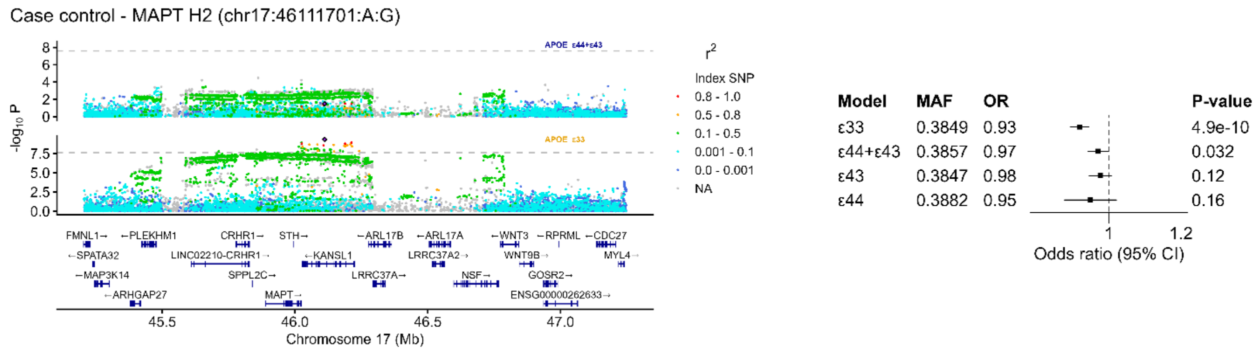


**Supplementary Figure 19: Loci and forest plot for *MAPT*.**

Loci plots for *MAPT* in both the *APOE* ε33 and *APOE* ε44+ε43 strata and forest plots for the lead SNPs for *MAPT* in the four *APOE* strata ε33, ε44+ε43, ε43 and ε44. In the forest plot each *APOE* strata is shown to visualize potential dominant or additive interaction.

### Supplementary Figure 20

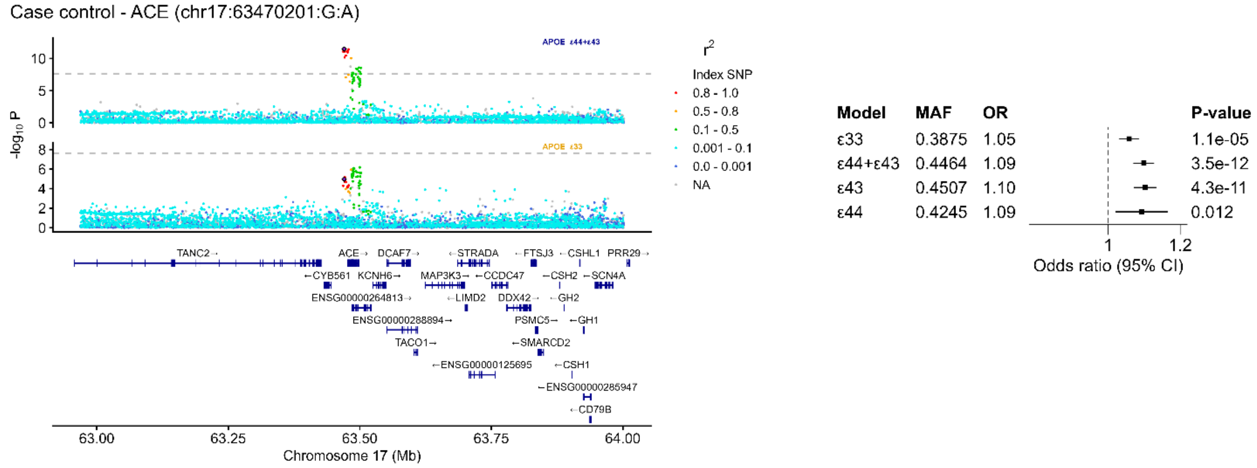


**Supplementary Figure 20: Loci and forest plot for *ACE.***

Loci plots for *ACE* in both the *APOE* ε33 and *APOE* ε44+ε43 strata and forest plots for the lead SNPs for *ACE* in the four *APOE* strata ε33, ε44+ε43, ε43 and ε44. In the forest plot each *APOE* strata is shown to visualize potential dominant or additive interaction.

### Supplementary Figure 21

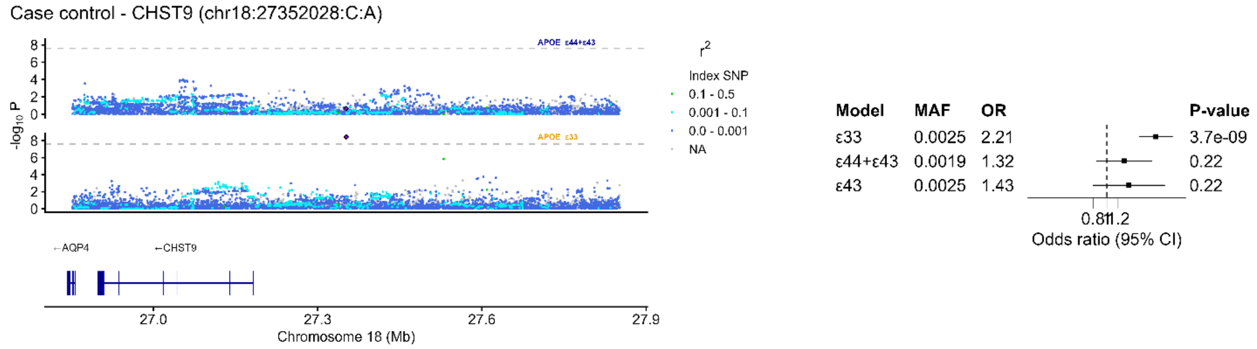


**Supplementary Figure 21: Loci and forest plot for *CHST9.***

Loci plots for *CHST9* in both the *APOE* ε33 and *APOE* ε44+ε43 strata and forest plots for the lead SNPs for *CHST9* in the four *APOE* strata ε33, ε44+ε43, ε43 and ε44. In the forest plot each *APOE* strata is shown to visualize potential dominant or additive interaction.

### Supplementary Figure 22

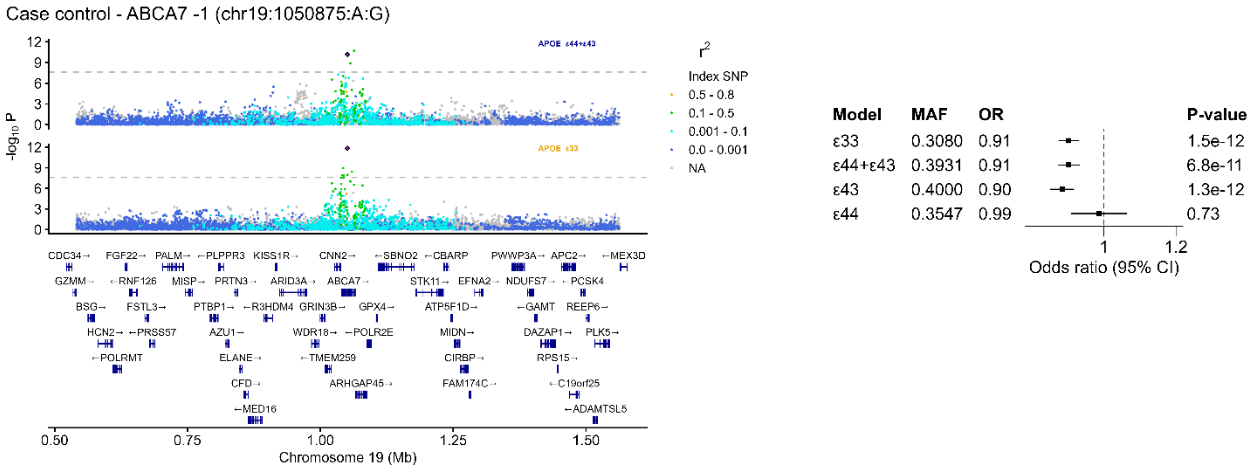


**Supplementary Figure 22: Loci and forest plot for *ABCA7*.**

Loci plots for *ABCA7* in both the *APOE* ε33 and *APOE* ε44+ε43 strata and forest plots for the lead SNPs for *ABCA7* in the four *APOE* strata ε33, ε44+ε43, ε43 and ε44. In the forest plot each *APOE* strata is shown to visualize potential dominant or additive interaction.

### Supplementary Figure 23

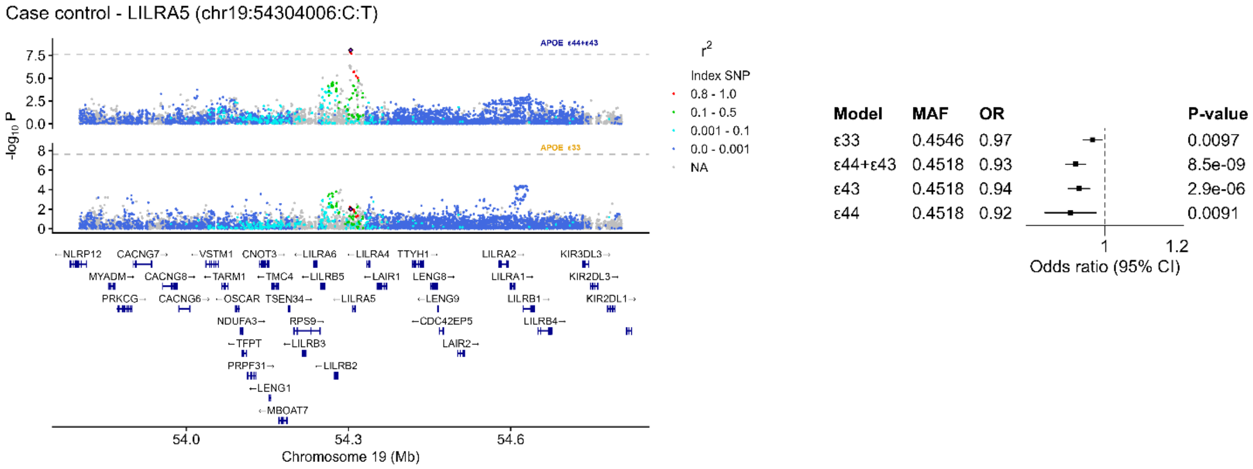


**Supplementary Figure 23: Loci and forest plot for *LILRA5*.**

Loci plots for *LILRA5* in both the *APOE* ε33 and *APOE* ε44+ε43 strata and forest plots for the lead SNPs for *LILRA5* in the four *APOE* strata ε33, ε44+ε43, ε43 and ε44. In the forest plot each *APOE* strata is shown to visualize potential dominant or additive interaction.

### Supplementary Figure 24

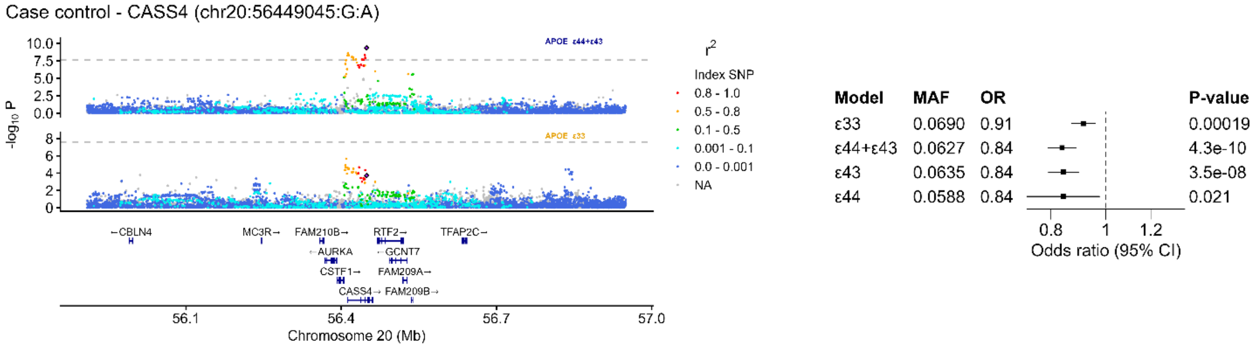


### Supplementary Figure 24: Loci and forest plot for *CASS4*

Loci plots for *CASS4* in both the *APOE* ε33 and *APOE* ε44+ε43 strata and forest plots for the lead SNPs for *CASS4* in the four *APOE* strata ε33, ε44+ε43, ε43 and ε44. In the forest plot each *APOE* strata is shown to visualize potential dominant or additive interaction.

**Supplementary Figure 25**

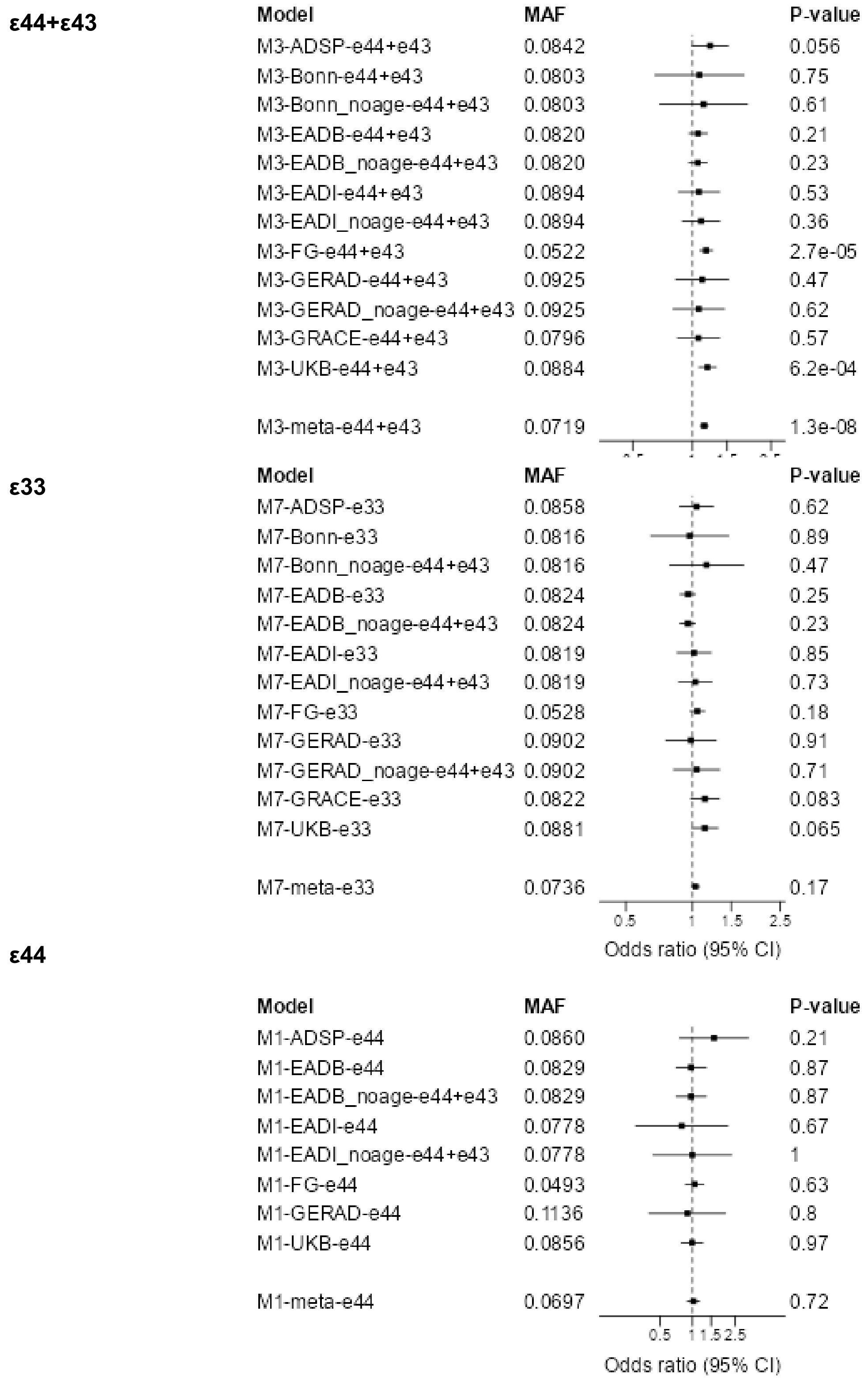


**Supplementary Figure 25: Forest plot across cohorts for *HP1BP3.***

Forest plots across cohorts for the lead SNPs for *HP1BP3* in the three *APOE* strata ε44+ε43, ε33, and ε44. Sensitivity analyses with no age adjustment is marked with noage and are not included in the meta-analysis. The forest plot of the ε44 strata is included to illustrate the robustness across cohorts in this high-risk context.

**Supplementary Figure 26**

**ε44+ε43**

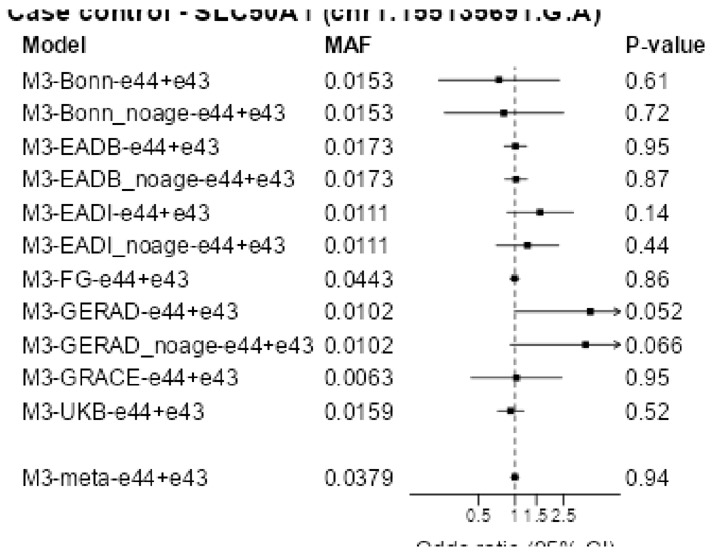


**ε33**

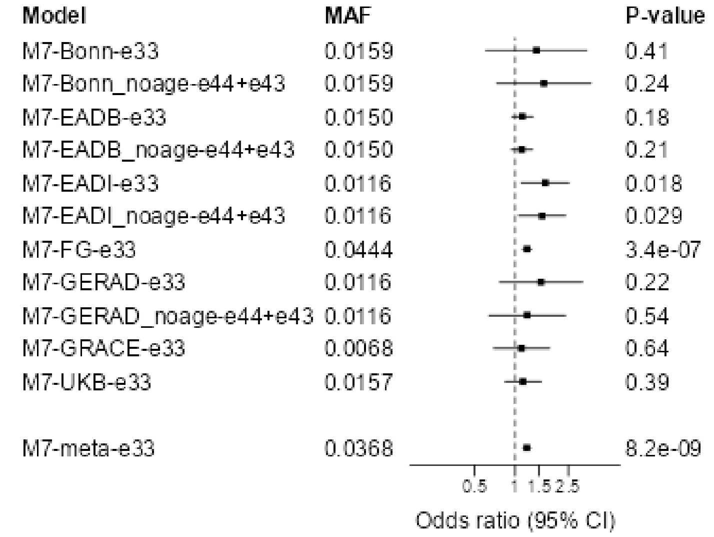


**ε44**

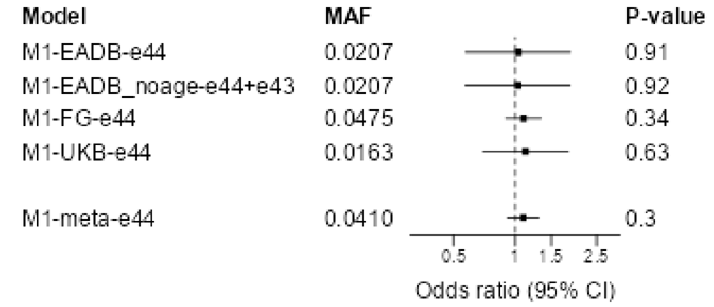


**Supplementary Figure 26: Forest plot across cohorts for *SLC50A1.***

Forest plots across cohorts for the lead SNPs for *SLC50A1* in the three *APOE* strata ε44+ε43, ε33, and ε44. Sensitivity analyses with no age adjustment is marked with noage and are not included in the meta-analysis. The forest plot of the ε44 strata is included to illustrate the robustness across cohorts in this high-risk context.

**Supplementary Figure 27**

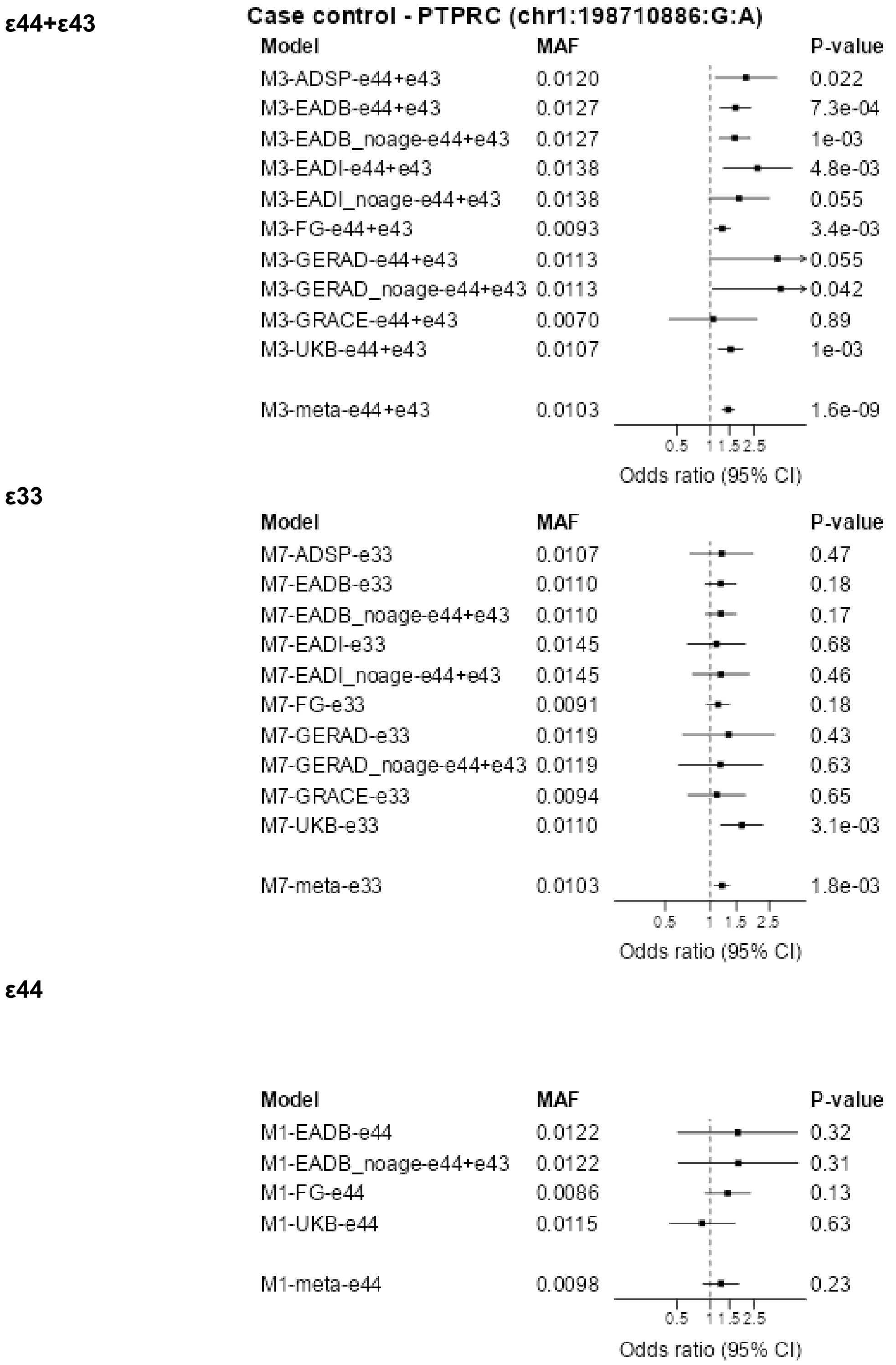


**Supplementary Figure 27 Forest plot across cohorts for *PTPRC*.**

Forest plots across cohorts for the lead SNPs for *PTPRC* in the three *APOE* strata ε44+ε43, ε33, and ε44. Sensitivity analyses with no age adjustment is marked with noage and are not included in the meta-analysis. The forest plot of the ε44 strata is included to illustrate the robustness across cohorts in this high-risk context.

**Supplementary Figure 28**

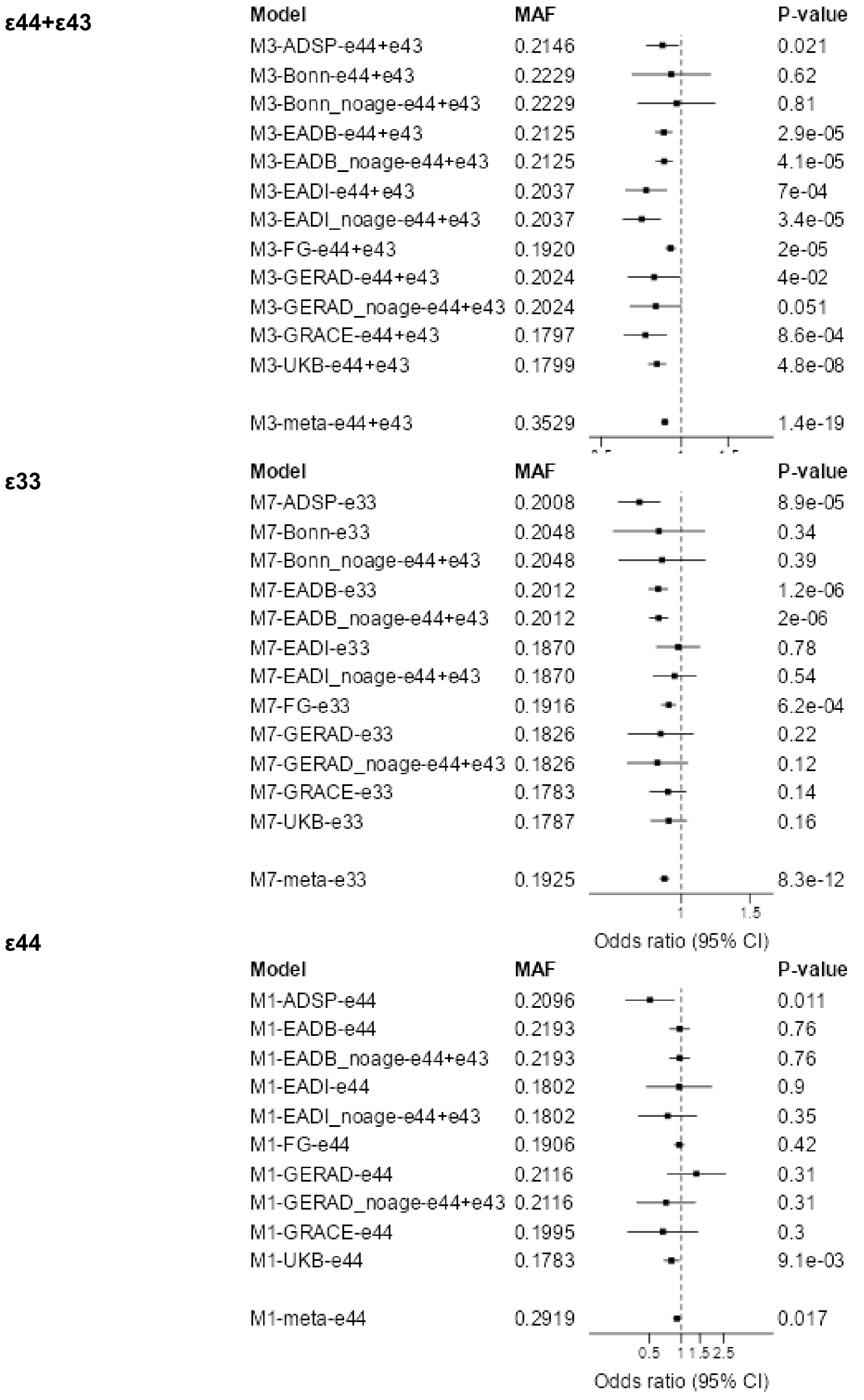


**Supplementary Figure 28: Forest plot across cohorts for *CR1*.**

Forest plots across cohorts for the lead SNPs for *CR1* in the three *APOE* strata ε44+ε43, ε33, and ε44. Sensitivity analyses with no age adjustment is marked with noage and are not included in the meta-analysis. The forest plot of the ε44 strata is included to illustrate the robustness across cohorts in this high-risk context.

**Supplementary Figure 29**

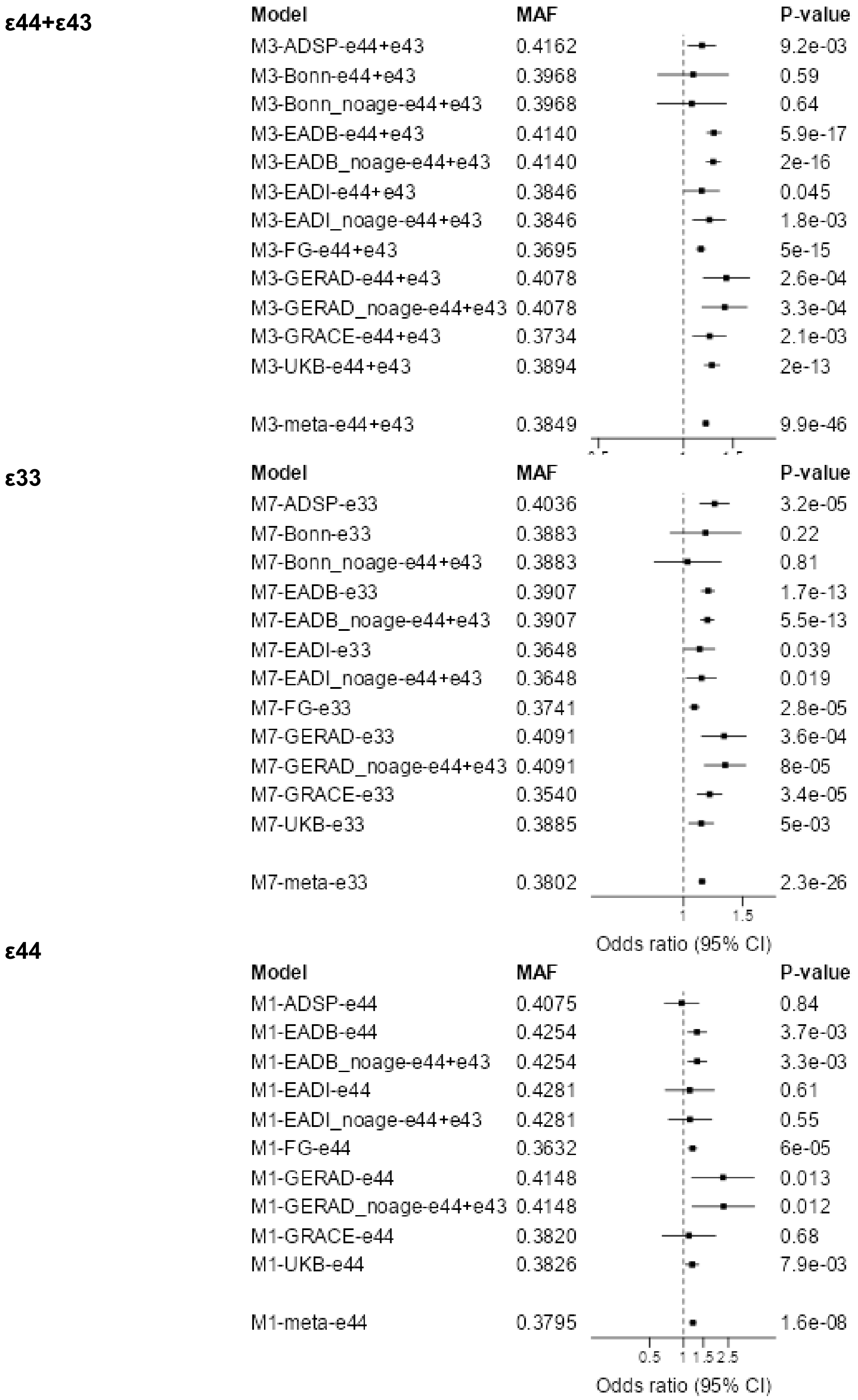


**Supplementary Figure 29: Forest plot across cohorts for *BIN1*.**

Forest plots across cohorts for the lead SNPs for *BIN1* in the three *APOE* strata ε44+ε43, ε33, and ε44. Sensitivity analyses with no age adjustment is marked with noage and are not included in the meta-analysis. The forest plot of the ε44 strata is included to illustrate the robustness across cohorts in this high-risk context.

**Supplementary Figure 30**

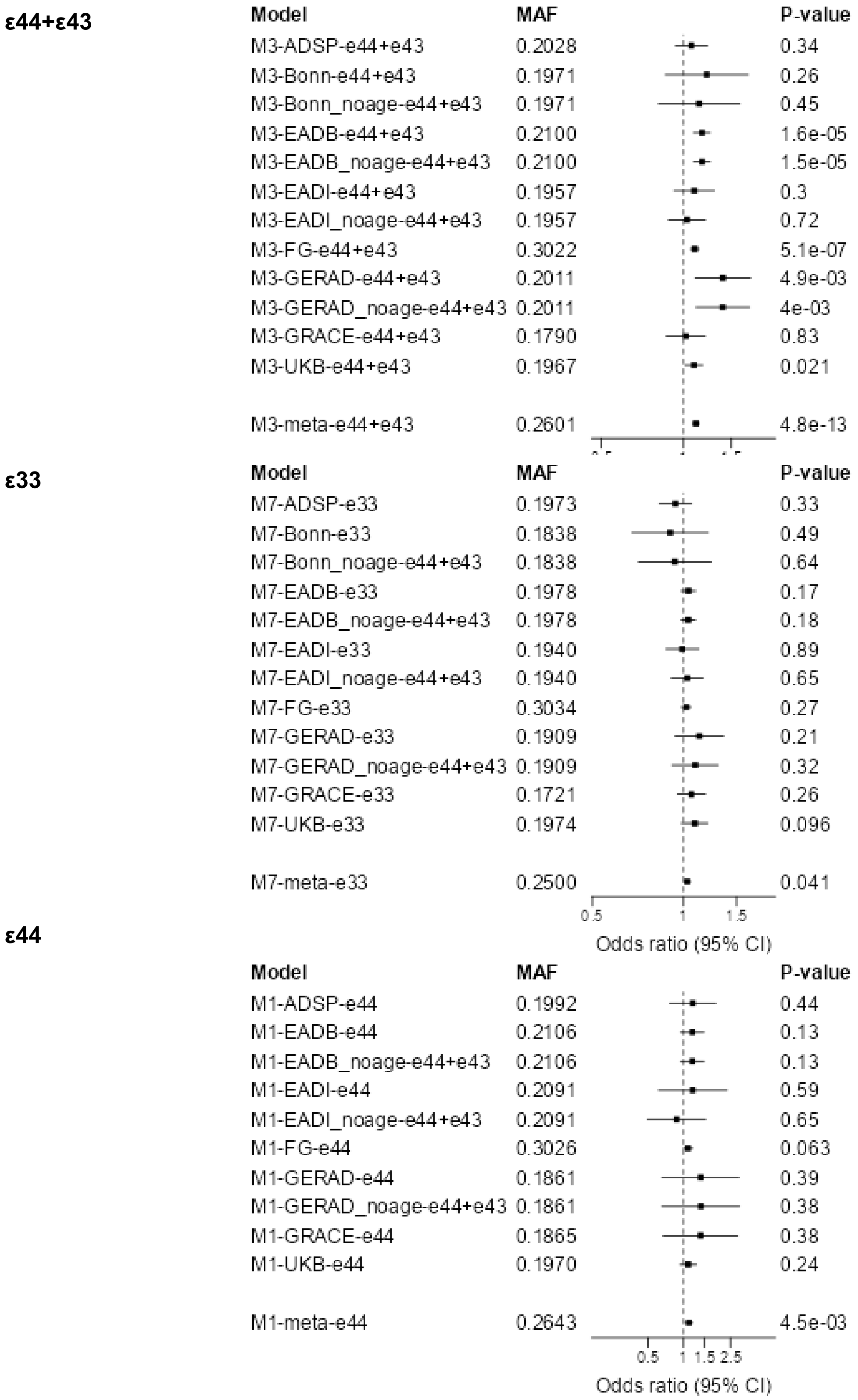


**Supplementary Figure 30: Forest plot across cohorts for *HLA-DRA -1.***

Forest plots across cohorts for the lead SNPs for *HLA-DRA -1* in the three *APOE* strata ε44+ε43, ε33, and ε44. Sensitivity analyses with no age adjustment is marked with noage and are not included in the meta-analysis. The forest plot of the ε44 strata is included to illustrate the robustness across cohorts in this high-risk context.

**Supplementary Figure 31**

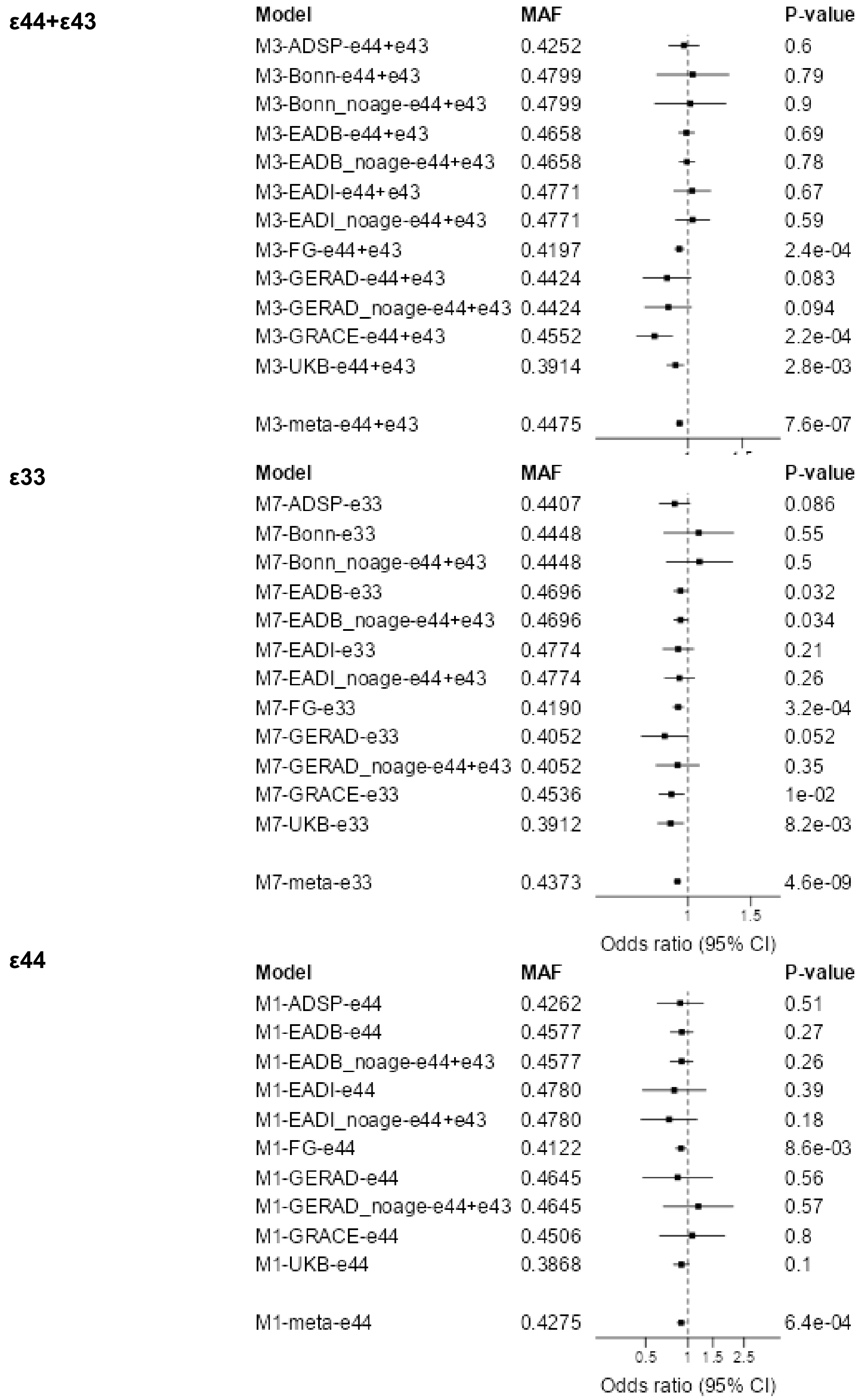


**Supplementary Figure 31: Forest plot across cohorts for *HLA-DRA -2.***

Forest plots across cohorts for the lead SNPs for *HLA-DRA -2* in the three *APOE* strata ε44+ε43, ε33, and ε44. Sensitivity analyses with no age adjustment is marked with noage and are not included in the meta-analysis. The forest plot of the ε44 strata is included to illustrate the robustness across cohorts in this high-risk context.

**Supplementary Figure 32**

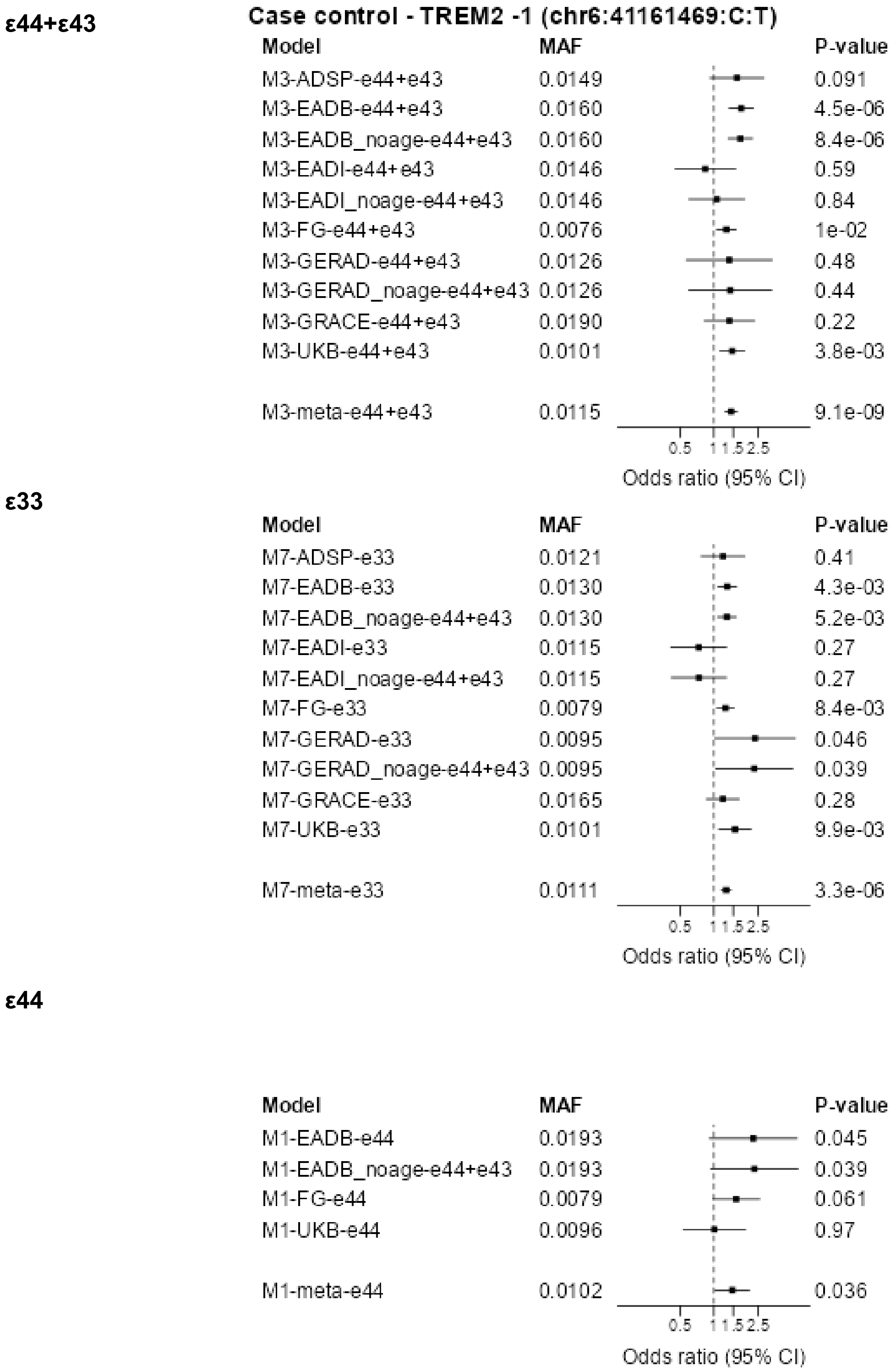


**Supplementary Figure 32: Forest plot across cohorts for *TREM2 -1***

Forest plots across cohorts for the lead SNPs for *TREM2 -1* in the three *APOE* strata ε44+ε43, ε33, and ε44. Sensitivity analyses with no age adjustment is marked with noage and are not included in the meta-analysis. The forest plot of the ε44 strata is included to illustrate the robustness across cohorts in this high-risk context.

**Supplementary Figure 33**

**ε44+ε43**

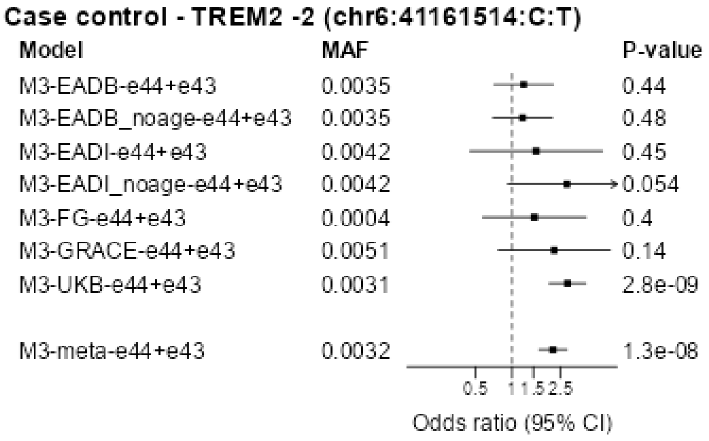


**ε33**

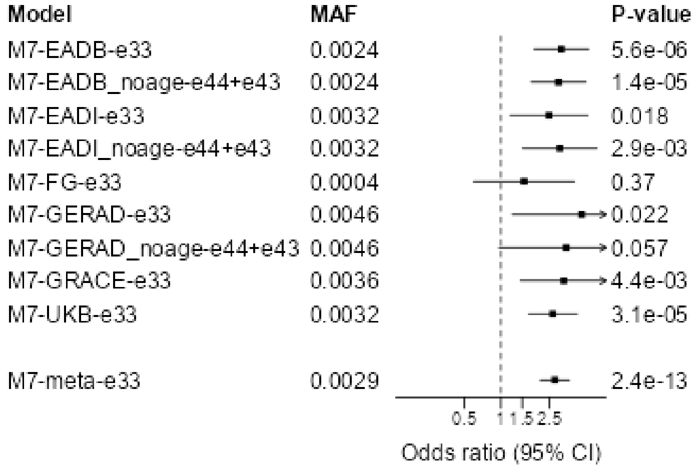


**Supplementary Figure 33: Forest plot across cohorts for *TREM2 -2***

Forest plots across cohorts for the lead SNPs for *TREM2 -2* in the three *APOE* strata ε44+ε43, ε33, and ε44. Sensitivity analyses with no age adjustment is marked with noage and are not included in the meta-analysis. The forest plot of the ε44 strata is included to illustrate the robustness across cohorts in this high-risk context.

**Supplementary Figure 34**

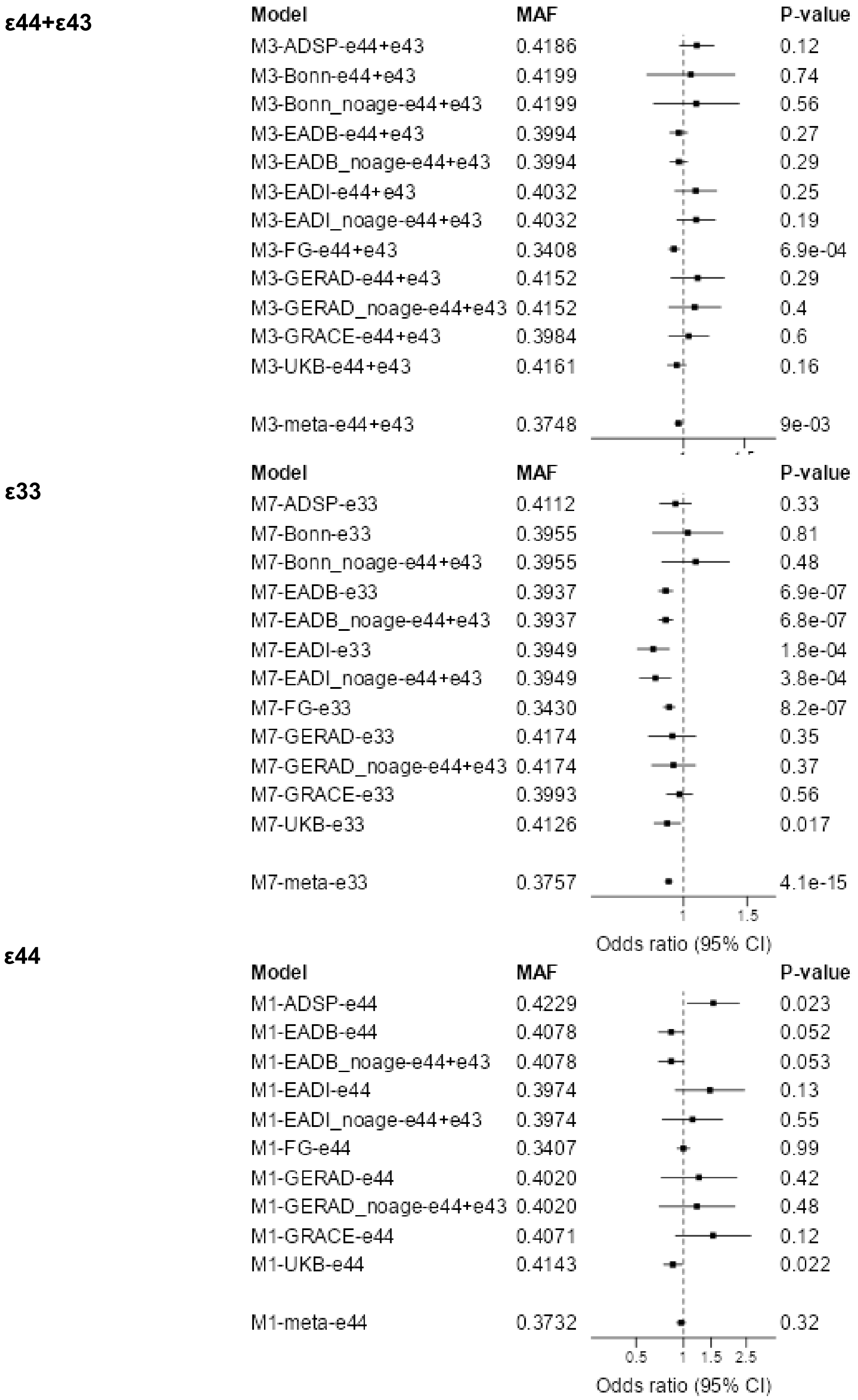


**Supplementary Figure 34: Forest plot across cohorts for *TMEM106B*.**

Forest plots across cohorts for the lead SNPs for *TMEM106B* in the three *APOE* strata ε44+ε43, ε33, and ε44. Sensitivity analyses with no age adjustment is marked with noage and are not included in the meta-analysis. The forest plot of the ε44 strata is included to illustrate the robustness across cohorts in this high-risk context.

**Supplementary Figure 35**

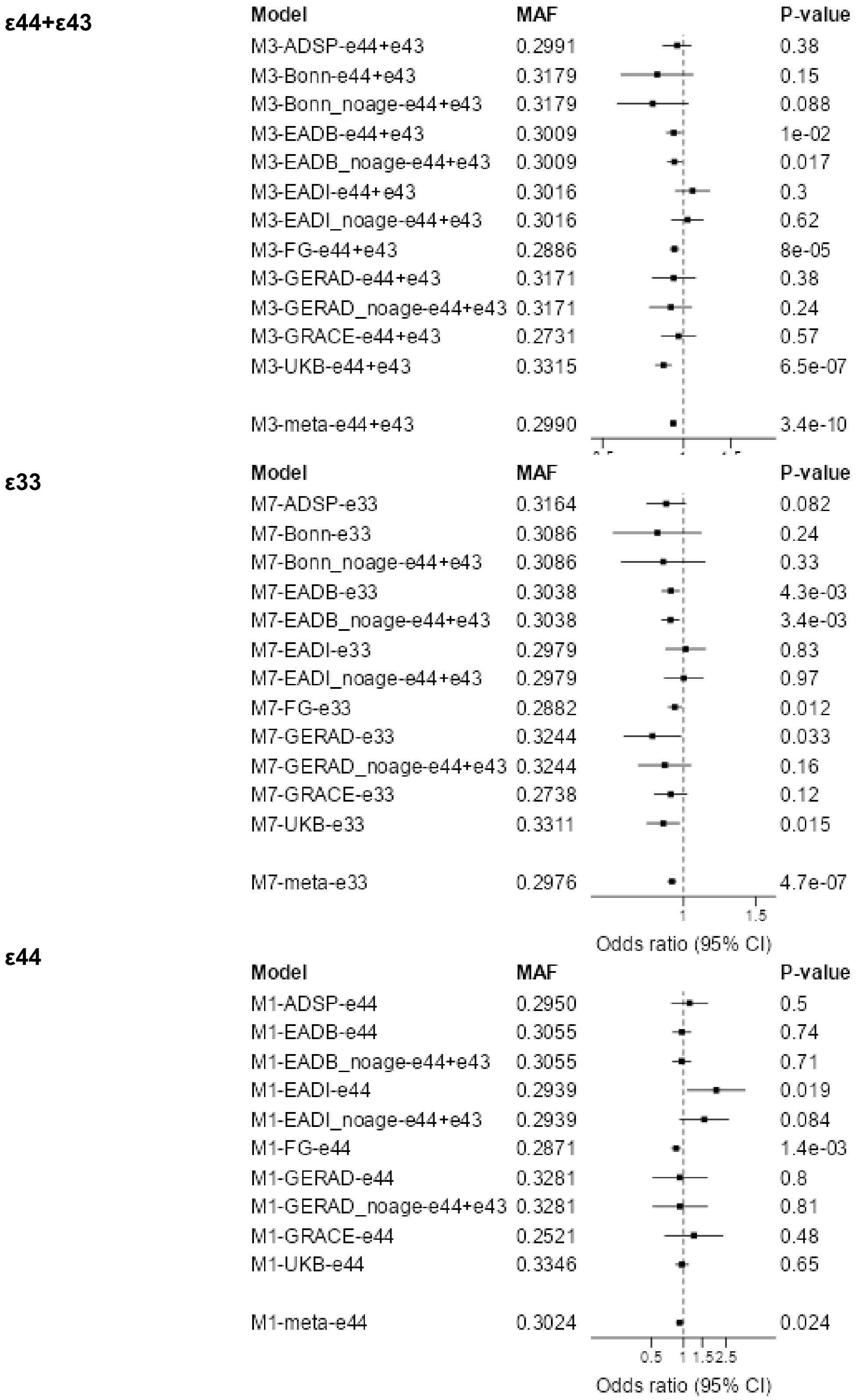


**Supplementary Figure 35: Forest plot across cohorts for *PLIRA*.**

Forest plots across cohorts for the lead SNPs for *PLIRA* in the three *APOE* strata ε44+ε43, ε33, and ε44. Sensitivity analyses with no age adjustment is marked with noage and are not included in the meta-analysis. The forest plot of the ε44 strata is included to illustrate the robustness across cohorts in this high-risk context.

**Supplementary Figure 36**

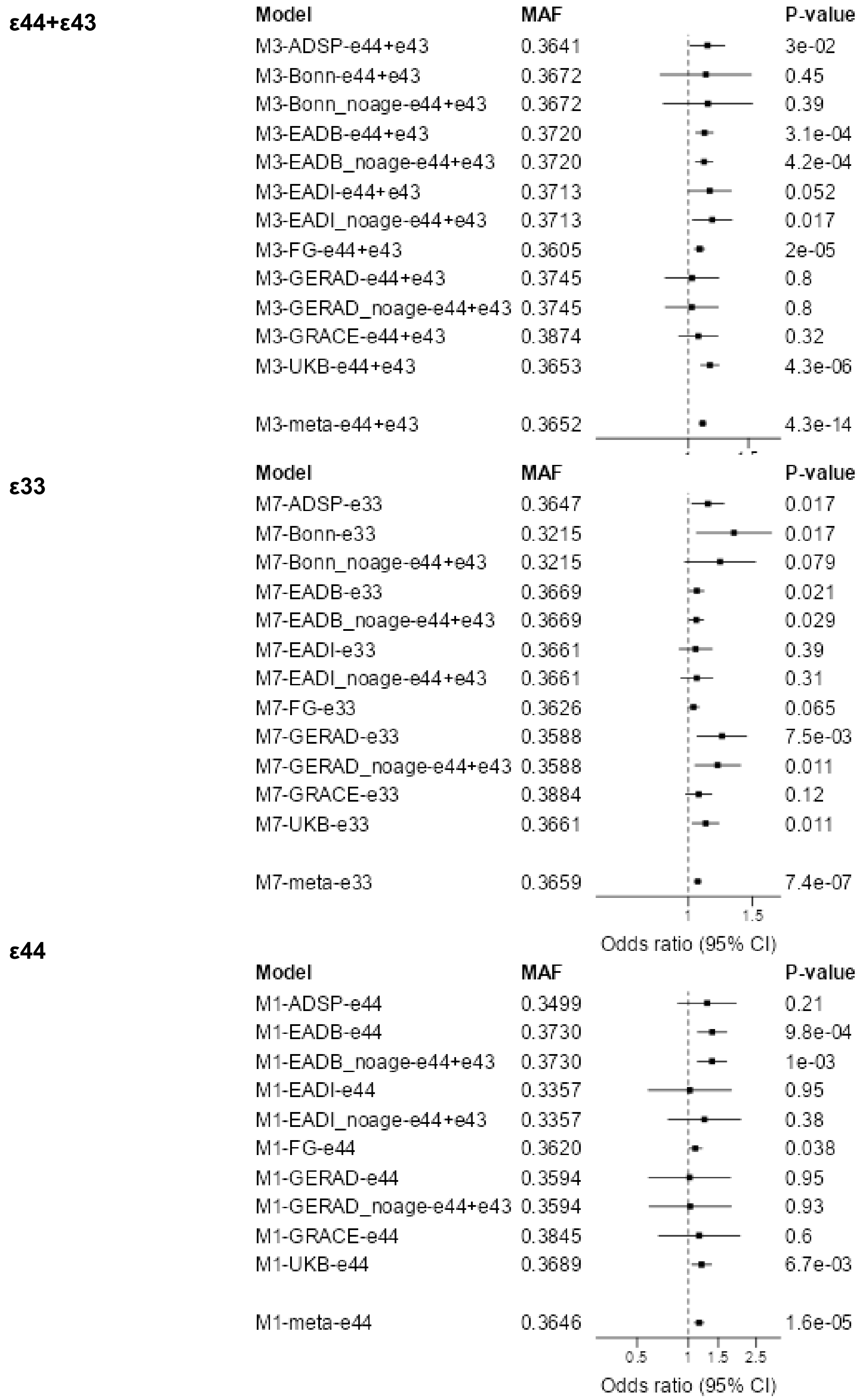


**Supplementary Figure 36: Forest plot across cohorts for *PTK2B*.**

Forest plots across cohorts for the lead SNPs for *PTK2B* in the three *APOE* strata ε44+ε43, ε33, and ε44. Sensitivity analyses with no age adjustment is marked with noage and are not included in the meta-analysis. The forest plot of the ε44 strata is included to illustrate the robustness across cohorts in this high-risk context.

**Supplementary Figure 37**

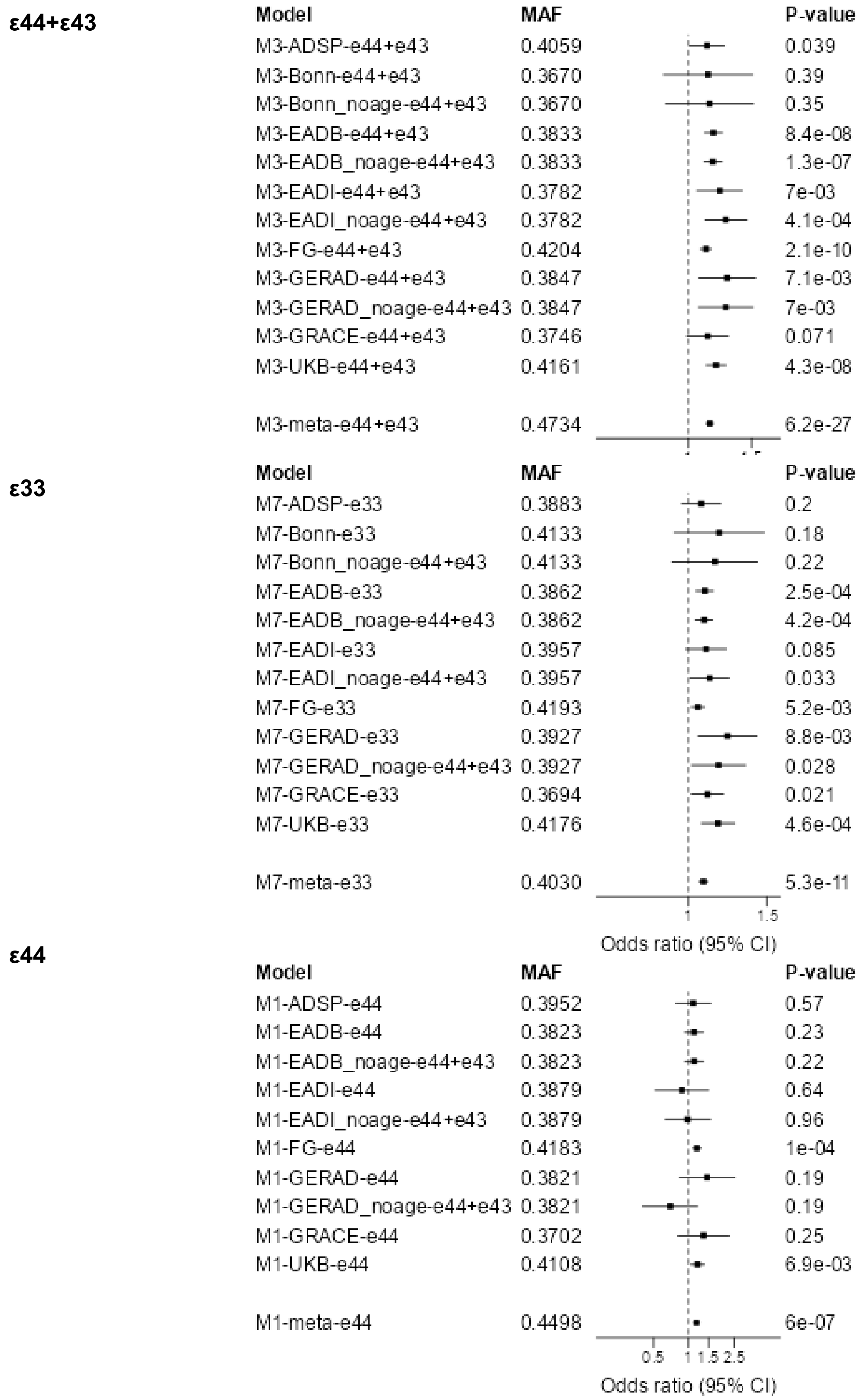


**Supplementary Figure 37: Forest plot across cohorts for *CLU*.**

Forest plots across cohorts for the lead SNPs for *CLU* in the three *APOE* strata ε44+ε43, ε33, and ε44. Sensitivity analyses with no age adjustment is marked with noage and are not included in the meta-analysis. The forest plot of the ε44 strata is included to illustrate the robustness across cohorts in this high-risk context.

**Supplementary Figure 38**

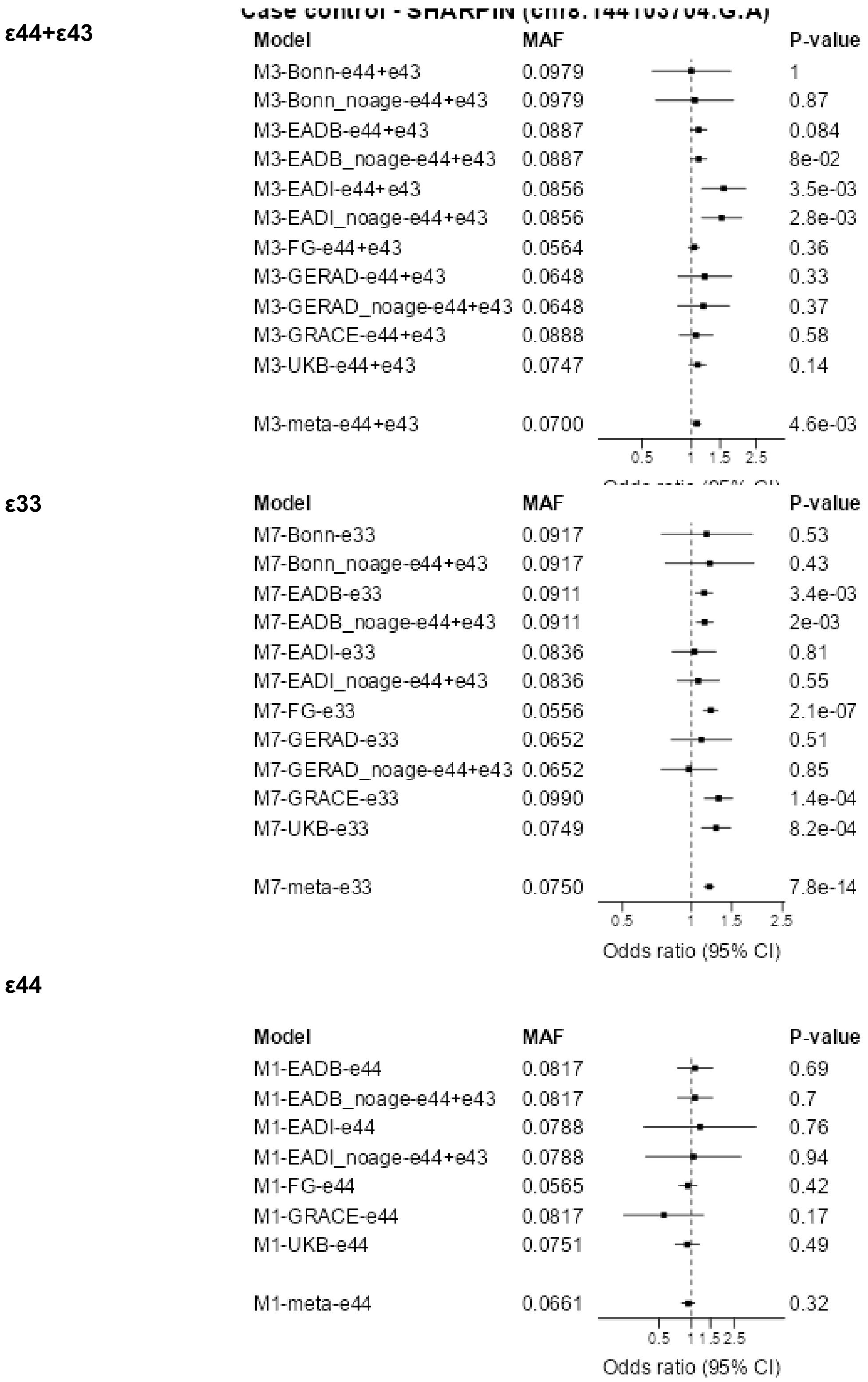


**Supplementary Figure 38: Forest plot across cohorts for *SHARPIN.***

Forest plots across cohorts for the lead SNPs for *SHARPIN* in the three *APOE* strata ε44+ε43, ε33, and ε44. Sensitivity analyses with no age adjustment is marked with noage and are not included in the meta-analysis. The forest plot of the ε44 strata is included to illustrate the robustness across cohorts in this high-risk context.

**Supplementary Figure 39**

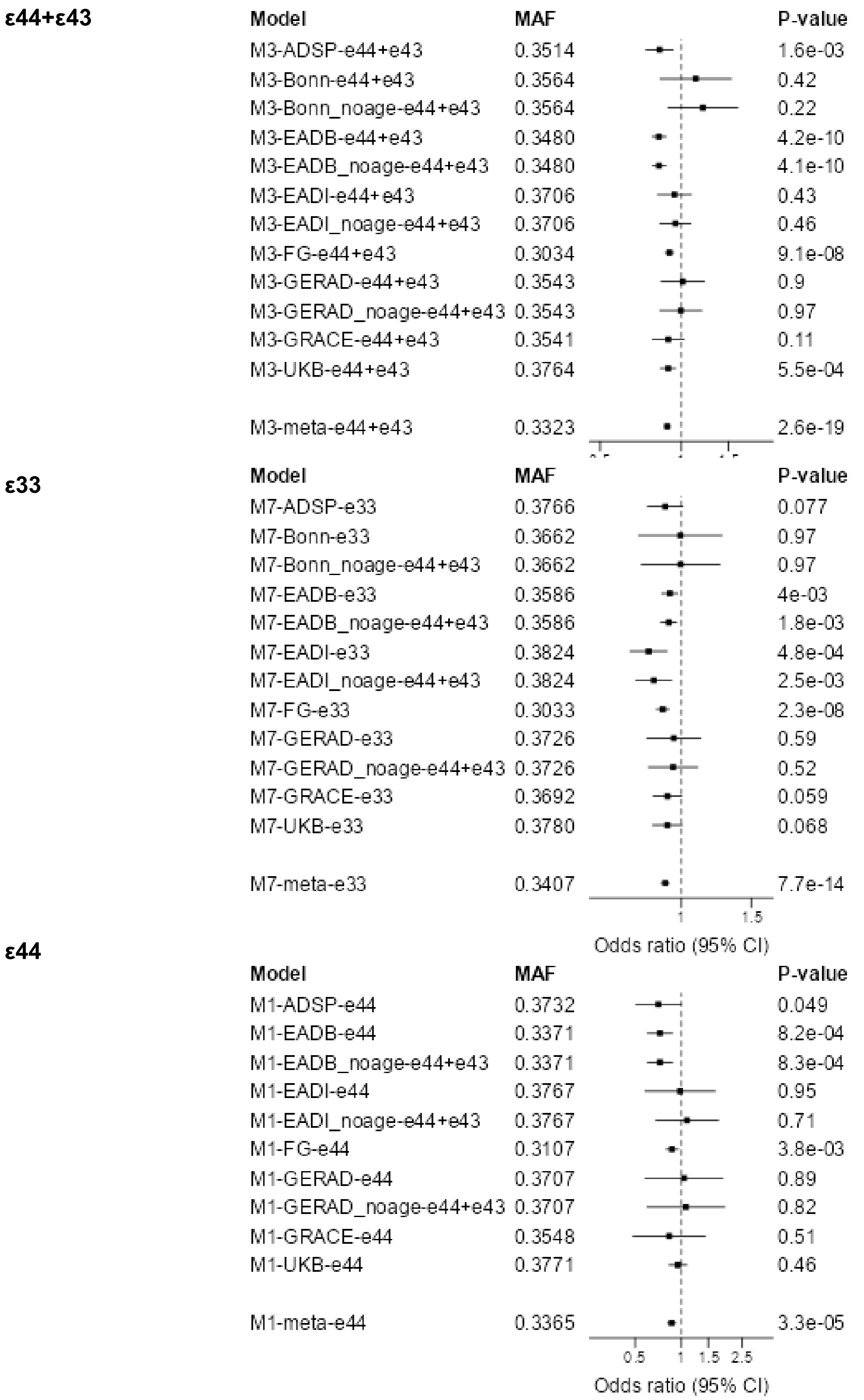


**Supplementary Figure 39: Forest plot across cohorts for *MS4A6A*.**

Forest plots across cohorts for the lead SNPs for *MS4A6A* in the three *APOE* strata ε44+ε43, ε33, and ε44. Sensitivity analyses with no age adjustment is marked with noage and are not included in the meta-analysis. The forest plot of the ε44 strata is included to illustrate the robustness across cohorts in this high-risk context.

**Supplementary Figure 40**

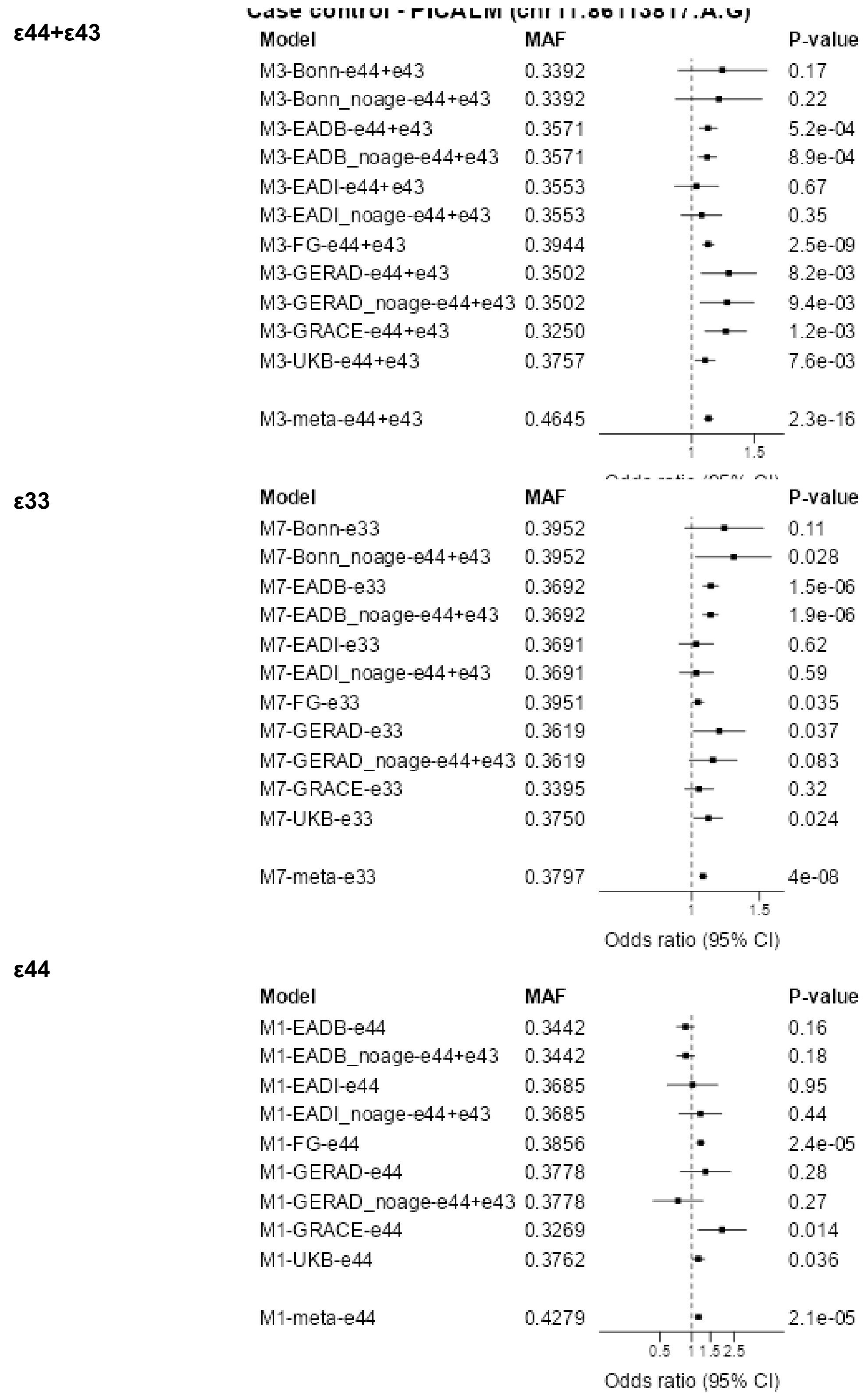


**Supplementary Figure 40: Forest plot across cohorts *PICALM***

Forest plots across cohorts for the lead SNPs for *PICALM* in the three *APOE* strata ε44+ε43, ε33, and ε44. Sensitivity analyses with no age adjustment is marked with noage and are not included in the meta-analysis. The forest plot of the ε44 strata is included to illustrate the robustness across cohorts in this high-risk context.

**Supplementary Figure 41**

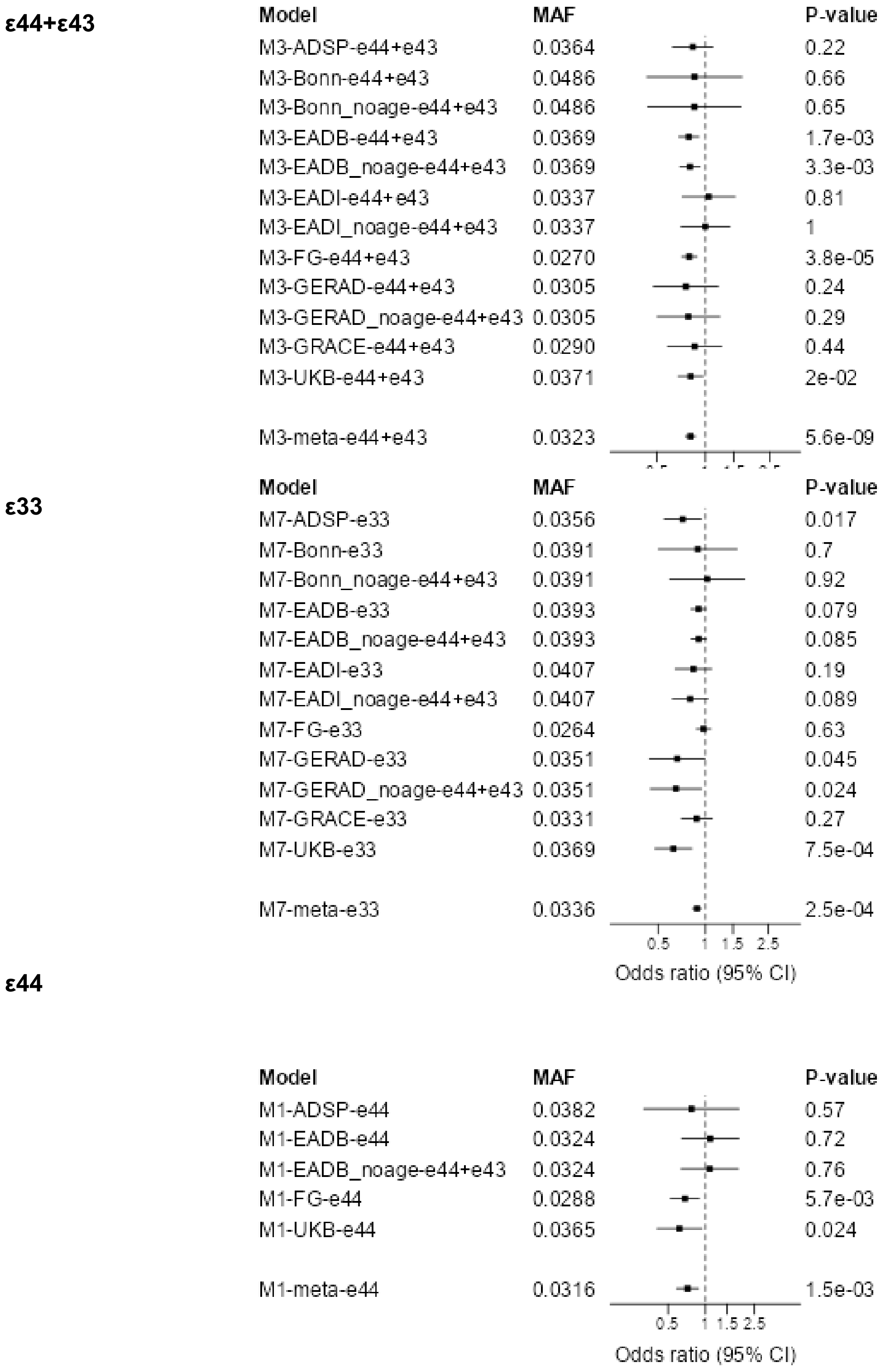


**Supplementary Figure 41: Forest plot across cohorts *SORL1*.**

Forest plots across cohorts for the lead SNPs for *SORL1* in the three *APOE* strata ε44+ε43, ε33, and ε44. Sensitivity analyses with no age adjustment is marked with noage and are not included in the meta-analysis. The forest plot of the ε44 strata is included to illustrate the robustness across cohorts in this high-risk context.

**Supplementary Figure 42**

**ε44+ε43**

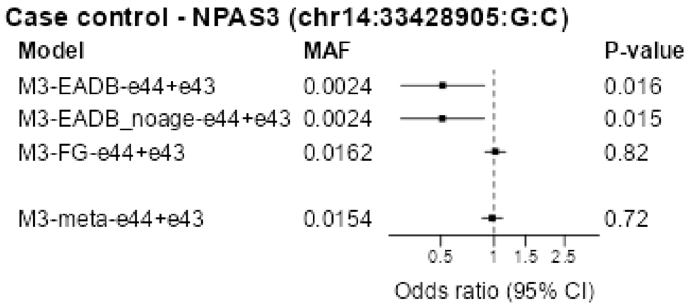


**ε33**

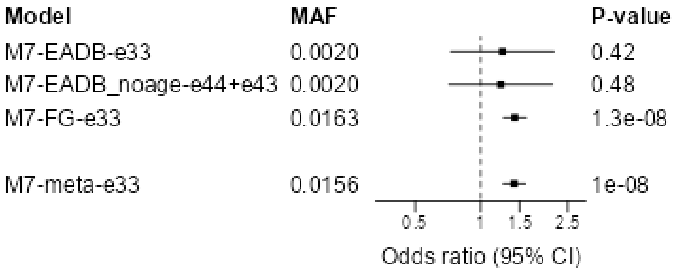


**ε44**

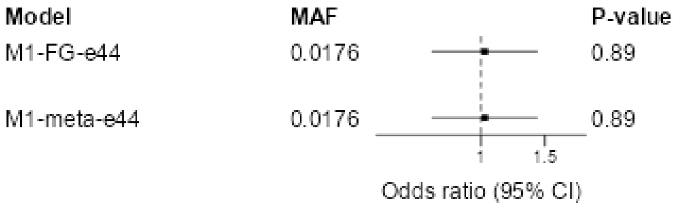


**Supplementary Figure 42: Forest plot across cohorts for *NPAS3.***

Forest plots across cohorts for the lead SNPs for *NPAS3* in the three *APOE* strata ε44+ε43, ε33, and ε44. Sensitivity analyses with no age adjustment is marked with noage and are not included in the meta-analysis. The forest plot of the ε44 strata is included to illustrate the robustness across cohorts in this high-risk context.

**Supplementary Figure 43**

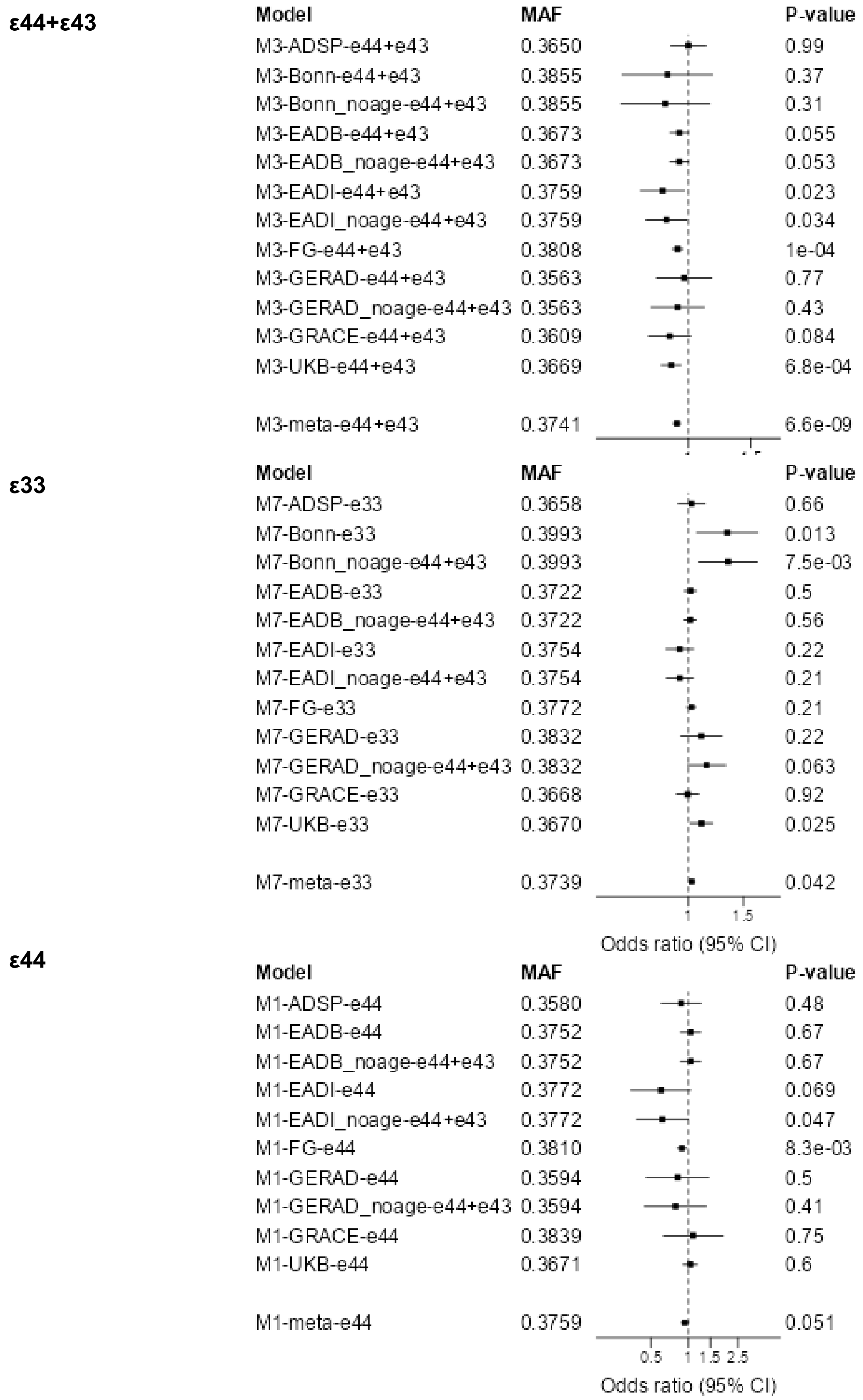


**Supplementary Figure 43: Forest plot across cohorts for *DDHD1*.**

Forest plots across cohorts for the lead SNPs for *DDHD1* in the three *APOE* strata ε44+ε43, ε33, and ε44. Sensitivity analyses with no age adjustment is marked with noage and are not included in the meta-analysis. The forest plot of the ε44 strata is included to illustrate the robustness across cohorts in this high-risk context.

**Supplementary Figure 44**

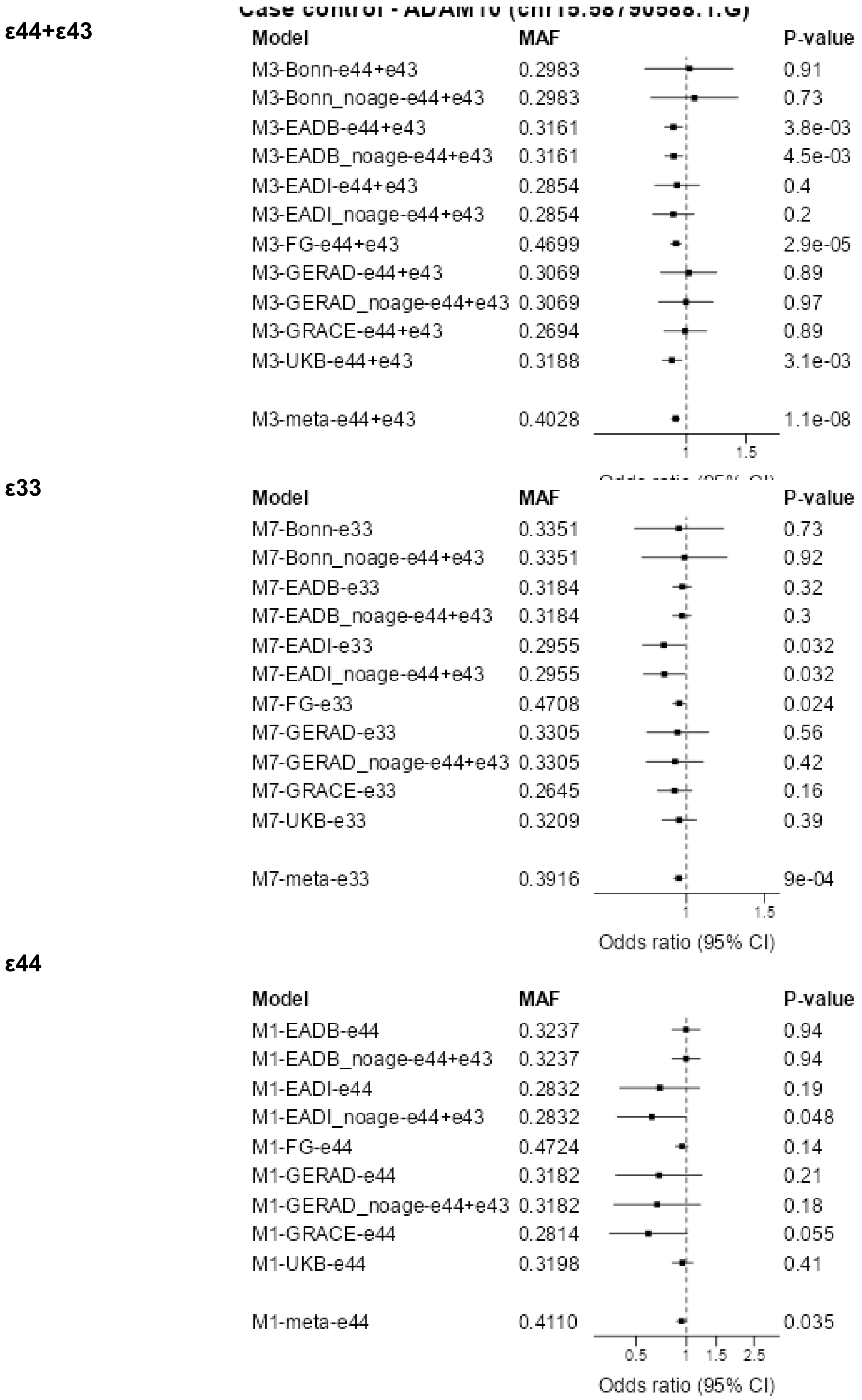


**Supplementary Figure 44: Forest plot across cohorts for *ADAM10.***

Forest plots across cohorts for the lead SNPs for *ADAM10* in the three *APOE* strata ε44+ε43, ε33, and ε44. Sensitivity analyses with no age adjustment is marked with noage and are not included in the meta-analysis. The forest plot of the ε44 strata is included to illustrate the robustness across cohorts in this high-risk context.

**Supplementary Figure 45**

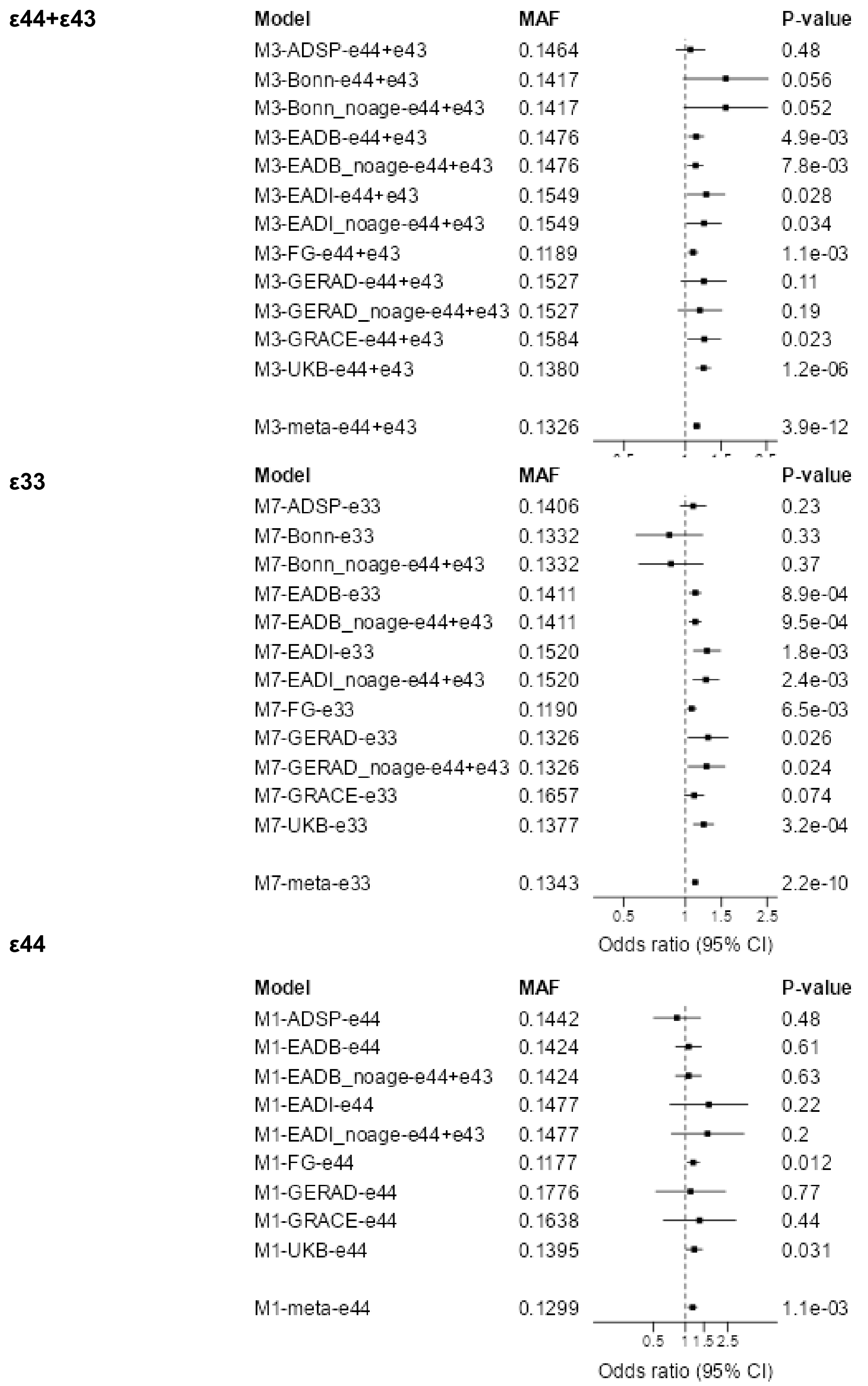


**Supplementary Figure 45: Forest plot across cohorts for *APH1B*.**

Forest plots across cohorts for the lead SNPs for *APH1B* in the three *APOE* strata ε44+ε43, ε33, and ε44. Sensitivity analyses with no age adjustment is marked with noage and are not included in the meta-analysis. The forest plot of the ε44 strata is included to illustrate the robustness across cohorts in this high-risk context.

**Supplementary Figure 46**

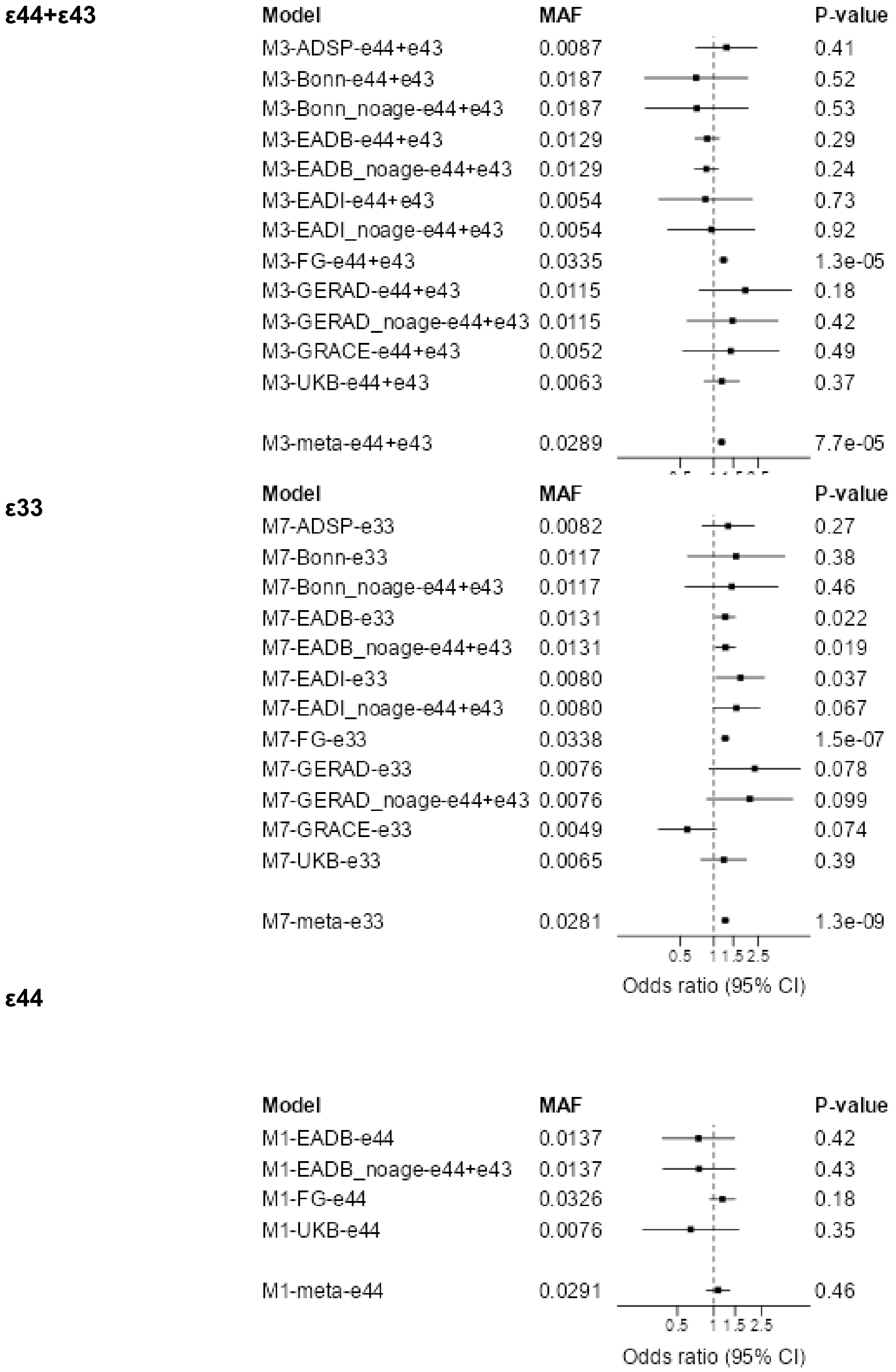


**Supplementary Figure 46: Forest plot across cohorts for *LINC02568.***

Forest plots across cohorts for the lead SNPs for *LINC02568* in the three *APOE* strata ε44+ε43, ε33, and ε44. Sensitivity analyses with no age adjustment is marked with noage and are not included in the meta-analysis. The forest plot of the ε44 strata is included to illustrate the robustness across cohorts in this high-risk context.

**Supplementary Figure 47**

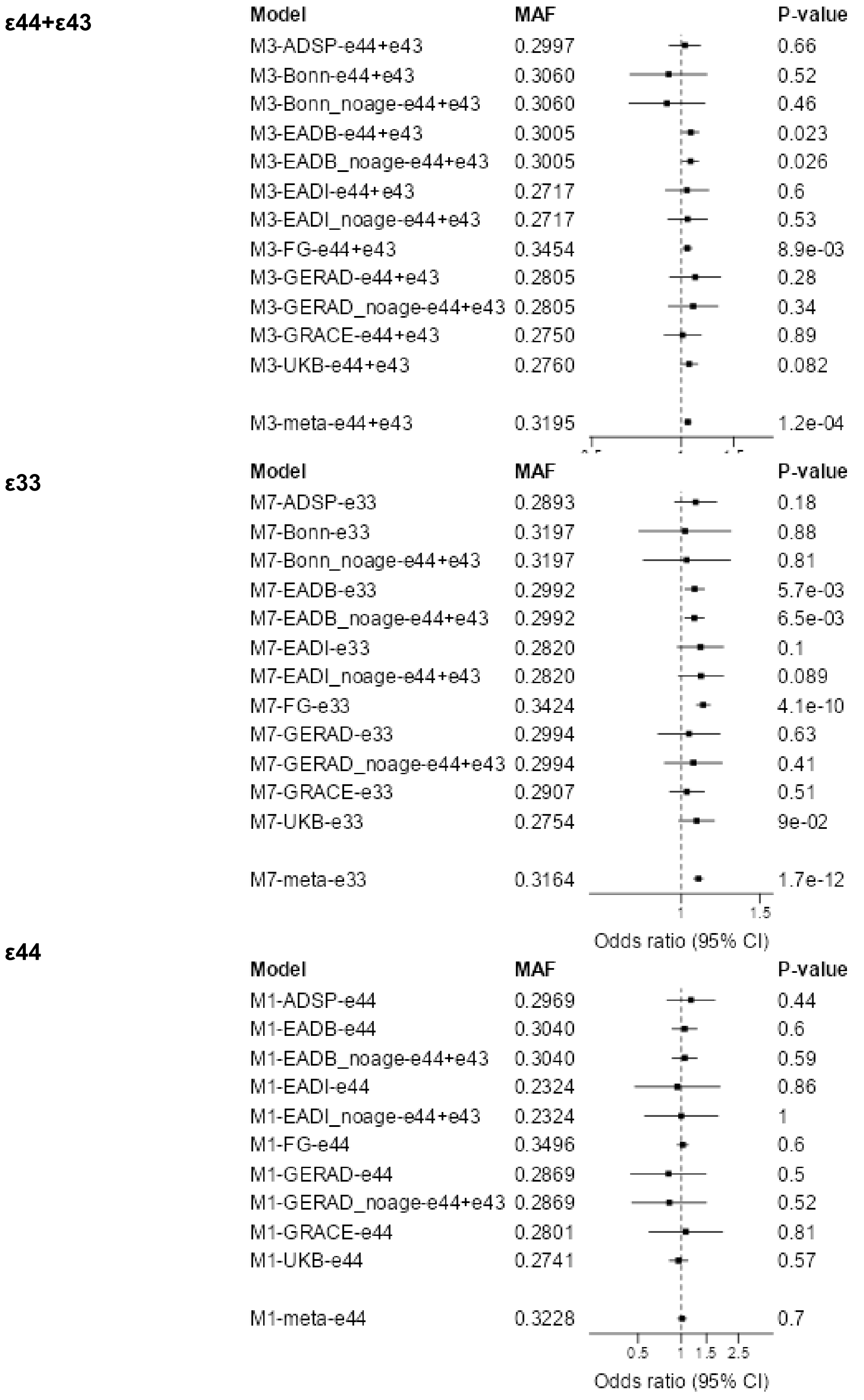


**Supplementary Figure 47: Forest plot across cohorts for *GRN*.**

Forest plots across cohorts for the lead SNPs for *GRN* in the three *APOE* strata ε44+ε43, ε33, and ε44. Sensitivity analyses with no age adjustment is marked with noage and are not included in the meta-analysis. The forest plot of the ε44 strata is included to illustrate the robustness across cohorts in this high-risk context.

**Supplementary Figure 48**

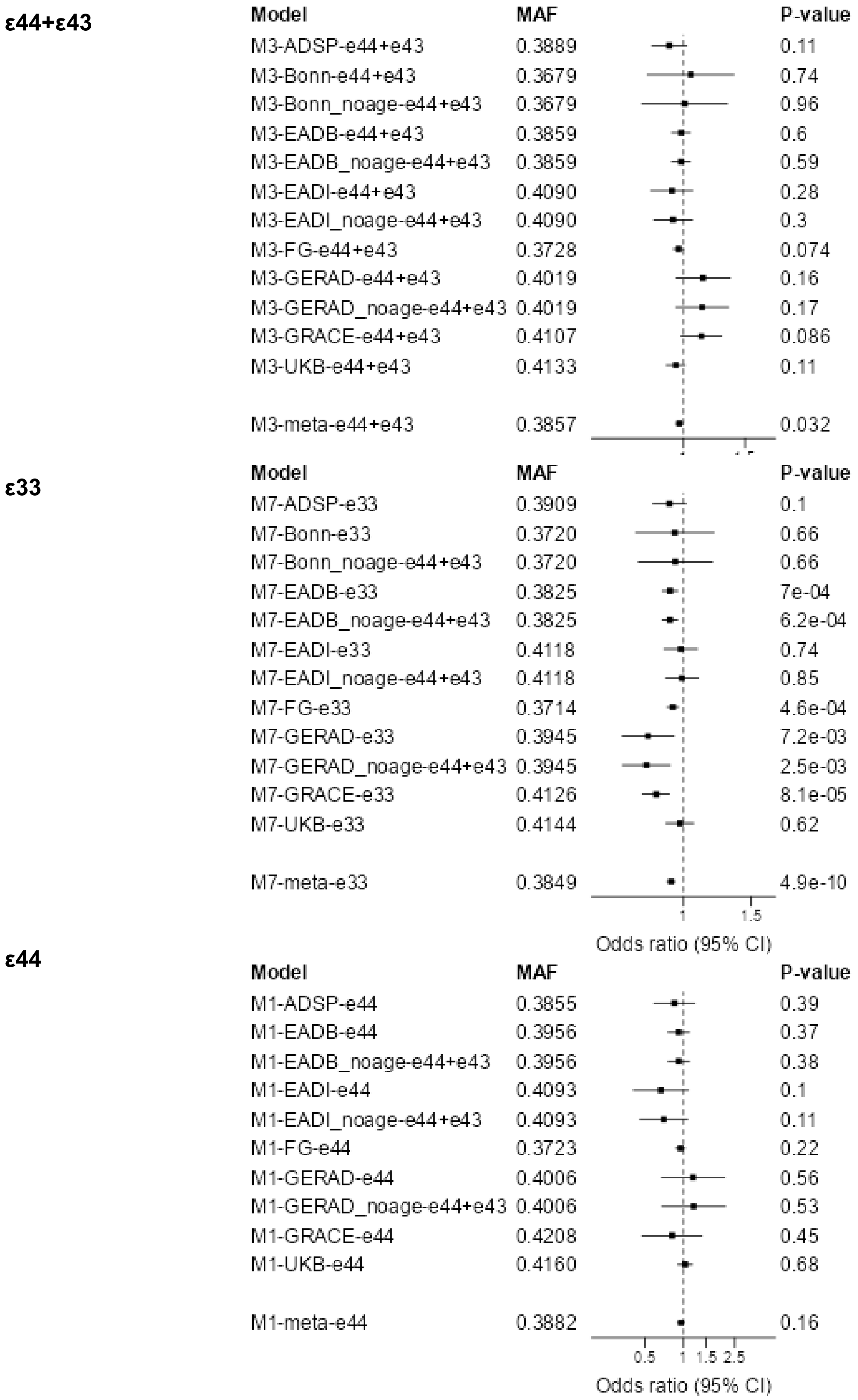


**Supplementary Figure 48: Forest plot across cohorts for *MAPT*.**

Forest plots across cohorts for the lead SNPs for *MAPT* in the three *APOE* strata ε44+ε43, ε33, and ε44. Sensitivity analyses with no age adjustment is marked with noage and are not included in the meta-analysis. The forest plot of the ε44 strata is included to illustrate the robustness across cohorts in this high-risk context.

**Supplementary Figure 49**

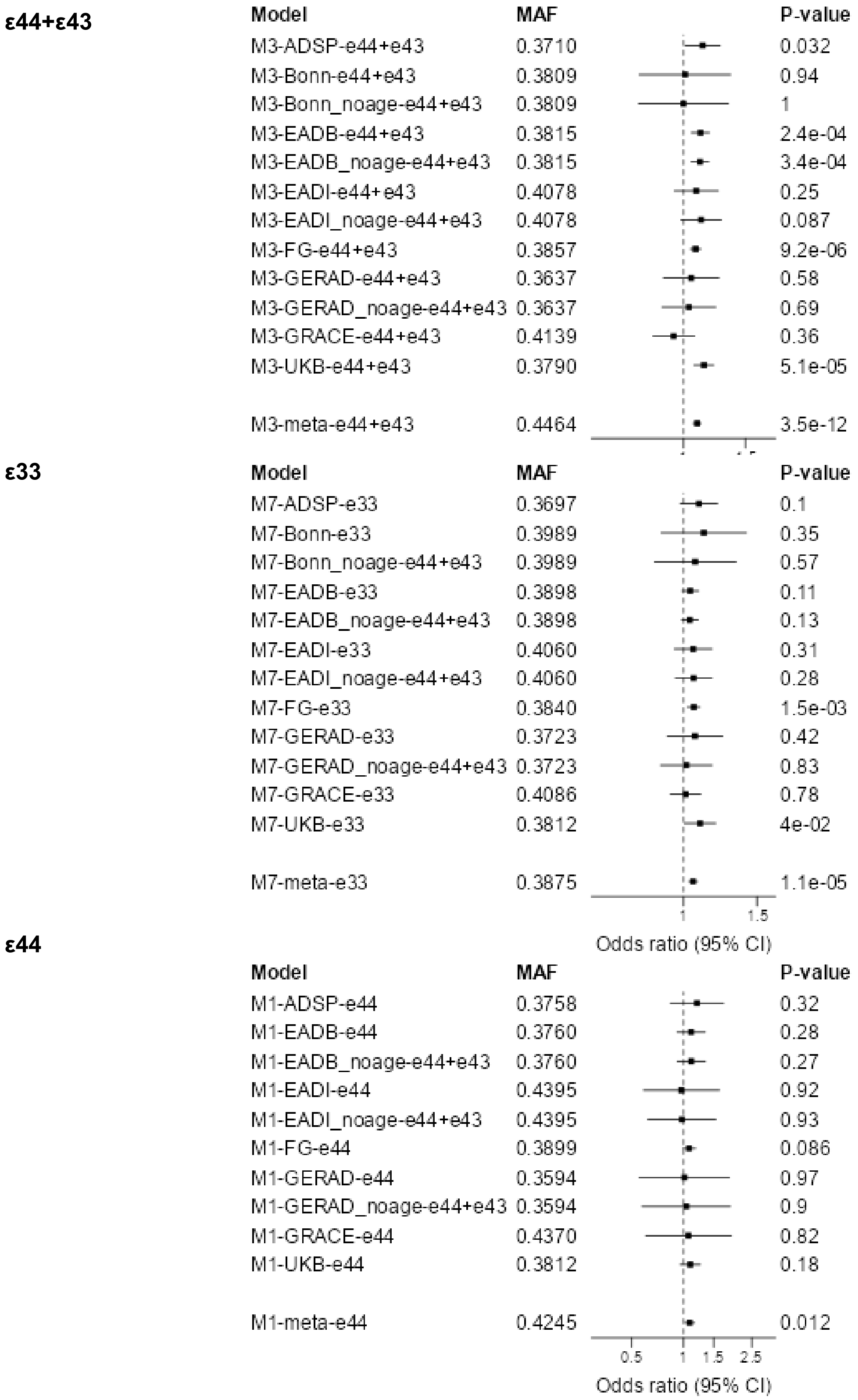


**Supplementary Figure 49: Forest plot across cohorts for *ACE*.**

Forest plots across cohorts for the lead SNPs for *ACE* in the three *APOE* strata ε44+ε43, ε33, and ε44. Sensitivity analyses with no age adjustment is marked with noage and are not included in the meta-analysis. The forest plot of the ε44 strata is included to illustrate the robustness across cohorts in this high-risk context.

**Supplementary Figure 50**

**ε44+ε43**

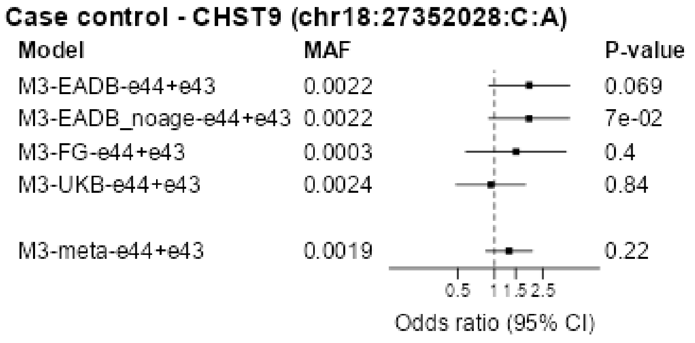


**ε33**

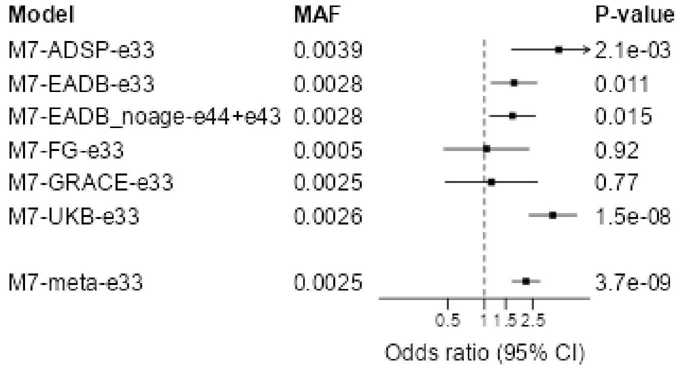


**Supplementary Figure 50: Forest plot across cohorts for *CHST9.***

Forest plots across cohorts for the lead SNPs for *CHST9* in the three *APOE* strata ε44+ε43, ε33, and ε44. Sensitivity analyses with no age adjustment is marked with noage and are not included in the meta-analysis. The forest plot of the ε44 strata is included to illustrate the robustness across cohorts in this high-risk context.

**Supplementary Figure 51**

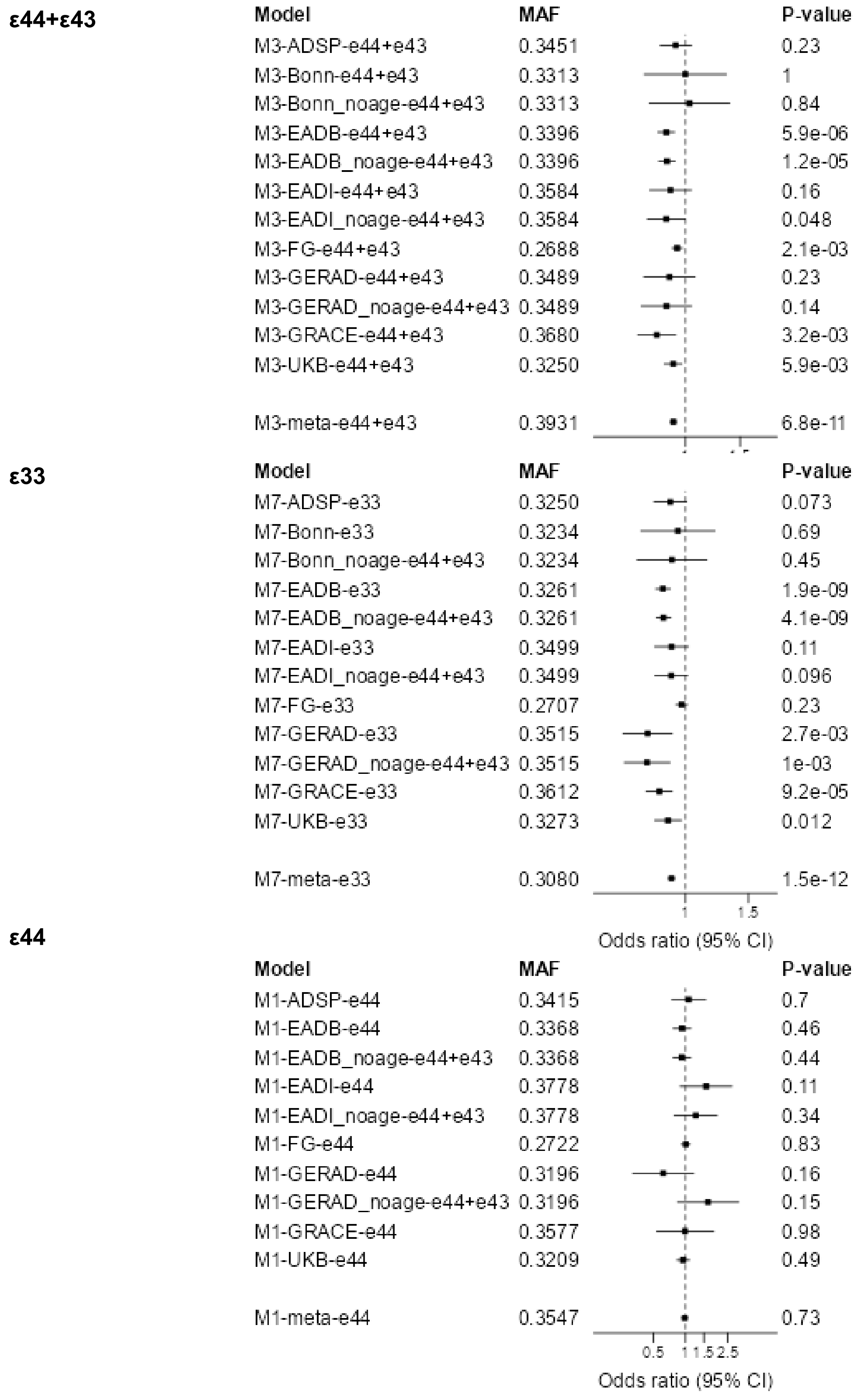


**Supplementary Figure 51: Forest plot across cohorts for *ABCA7*.**

Forest plots across cohorts for the lead SNPs for *ABCA7* in the three *APOE* strata ε44+ε43, ε33, and ε44. Sensitivity analyses with no age adjustment is marked with noage and are not included in the meta-analysis. The forest plot of the ε44 strata is included to illustrate the robustness across cohorts in this high-risk context.

**Supplementary Figure 52**

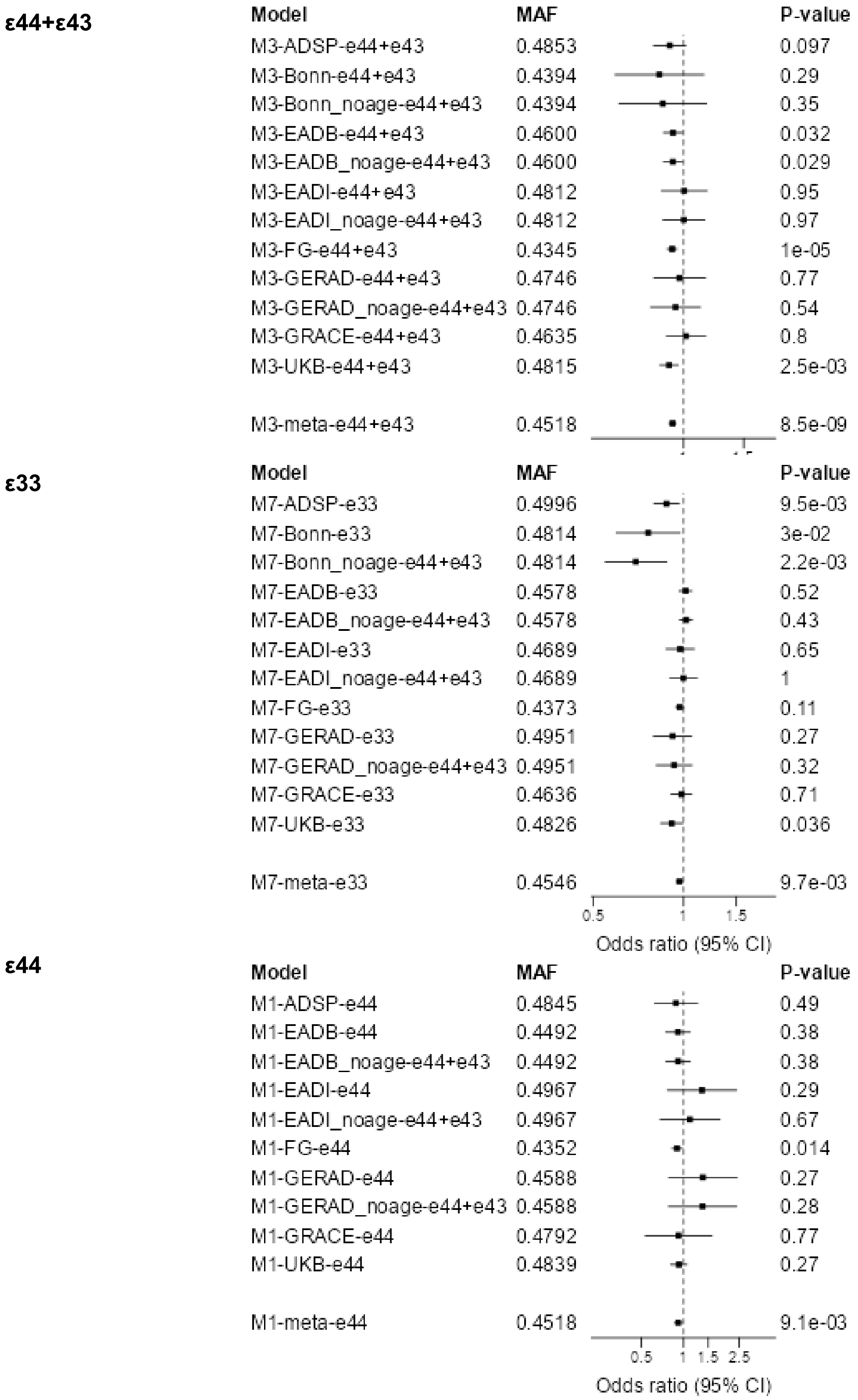


**Supplementary Figure 52: Forest plot across cohorts for *LILRA5.***

Forest plots across cohorts for the lead SNPs for *LILRA5* in the three *APOE* strata ε44+ε43, ε33, and ε44. Sensitivity analyses with no age adjustment is marked with noage and are not included in the meta-analysis. The forest plot of the ε44 strata is included to illustrate the robustness across cohorts in this high-risk context.

**Supplementary Figure 53**

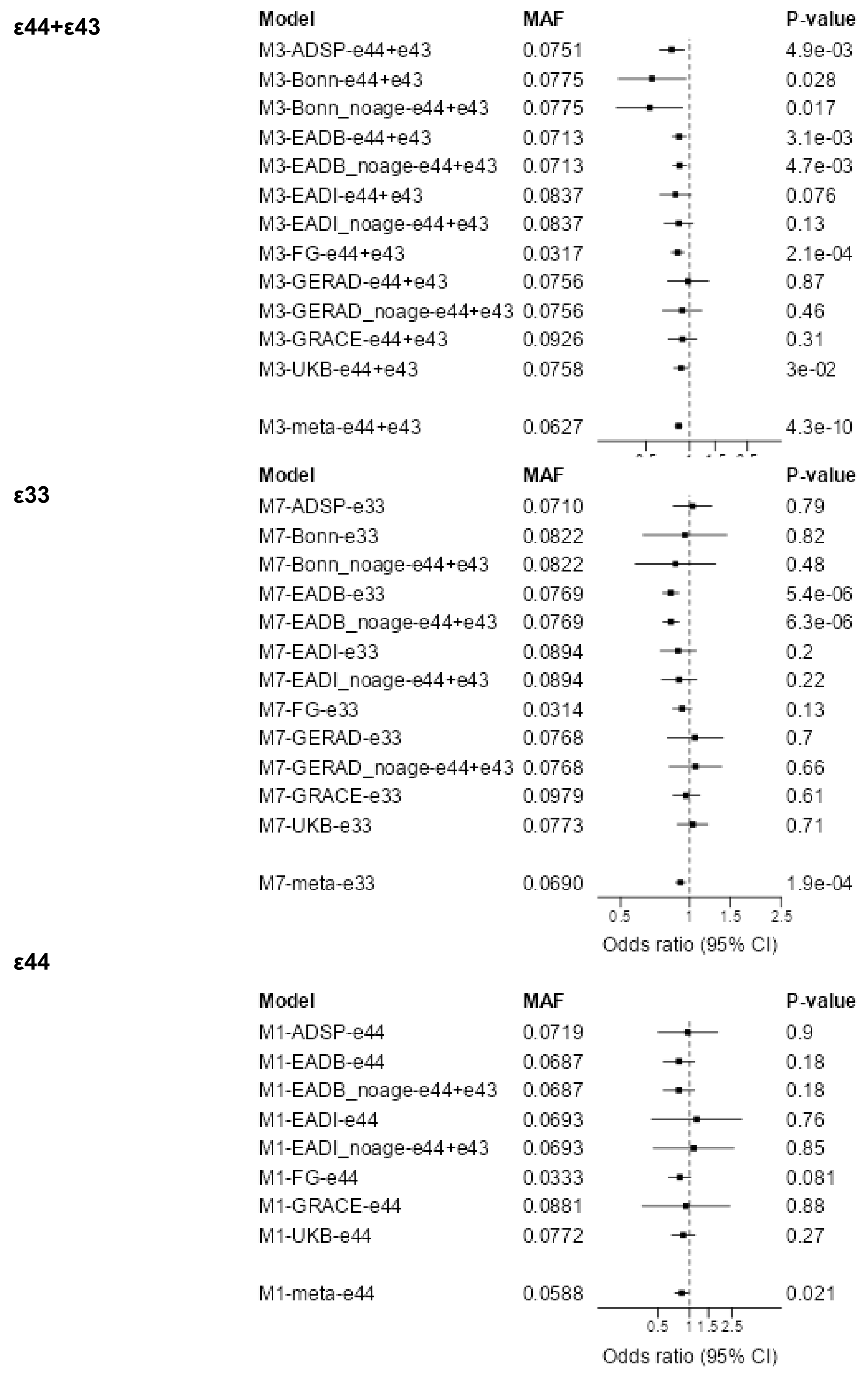


**Supplementary Figure 53: Forest plot across cohorts for *CASS4***

Forest plots across cohorts for the lead SNPs for *CASS4* in the three *APOE* strata ε44+ε43, ε33, and ε44. Sensitivity analyses with no age adjustment is marked with noage and are not included in the meta-analysis. The forest plot of the ε44 strata is included to illustrate the robustness across cohorts in this high-risk context.

### Supplementary Figure 54

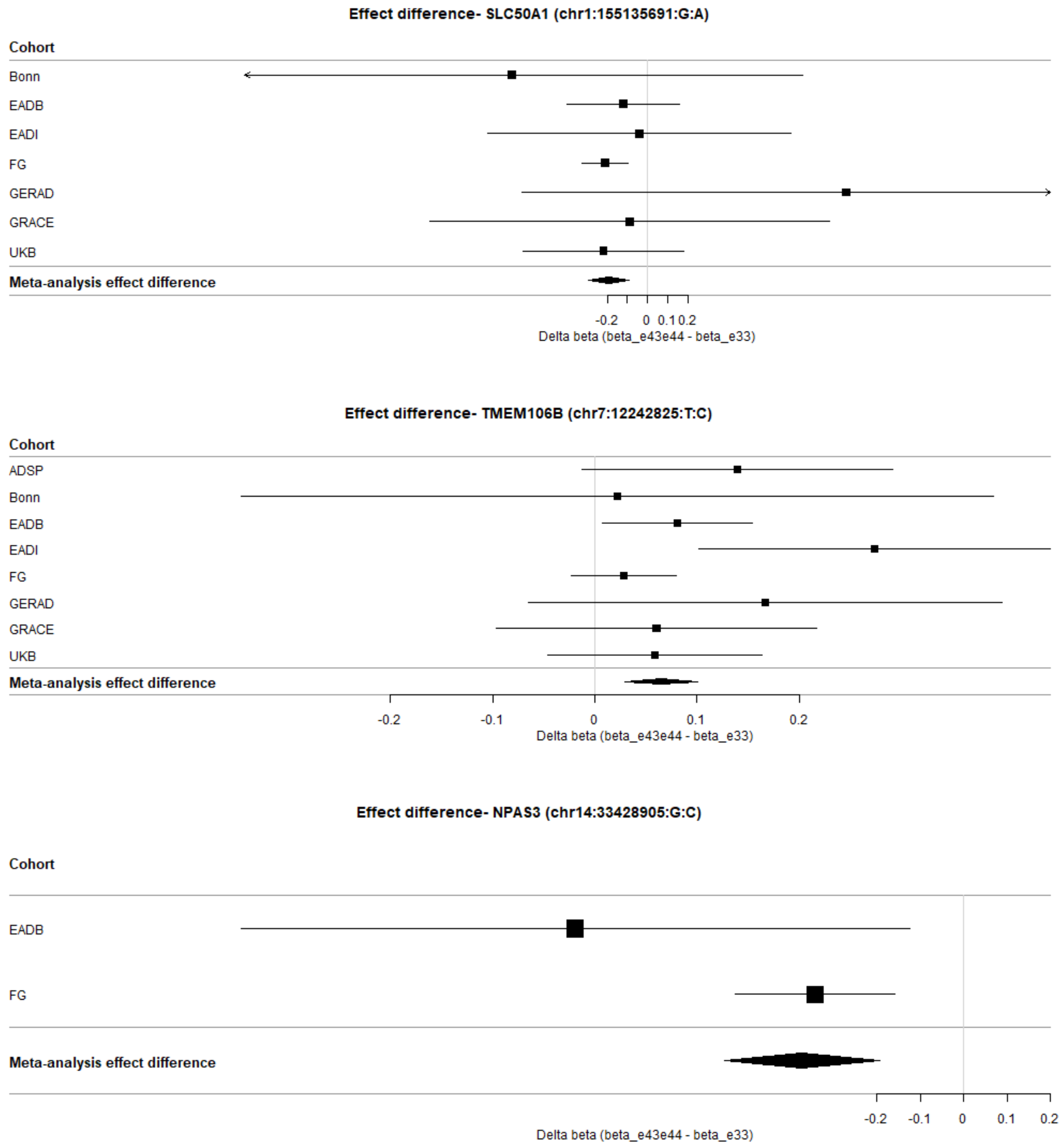


**Supplementary Figure 54: Forest plot for the effect difference between *APOE* strata ε44+ε43 and ε33 for loci with attenuated effect with the *APOE* ε4 allele.**

Forest plots for *SLC50A1*, *TMEM106B* and *NPAS3* of the effect difference between *APOE* strata ε44+ε43 and ε33. Note that the effect difference has been calculated with respect to the TOPMed reference allele. Only loci with dominant interaction are shown.

### Supplementary Figure 55

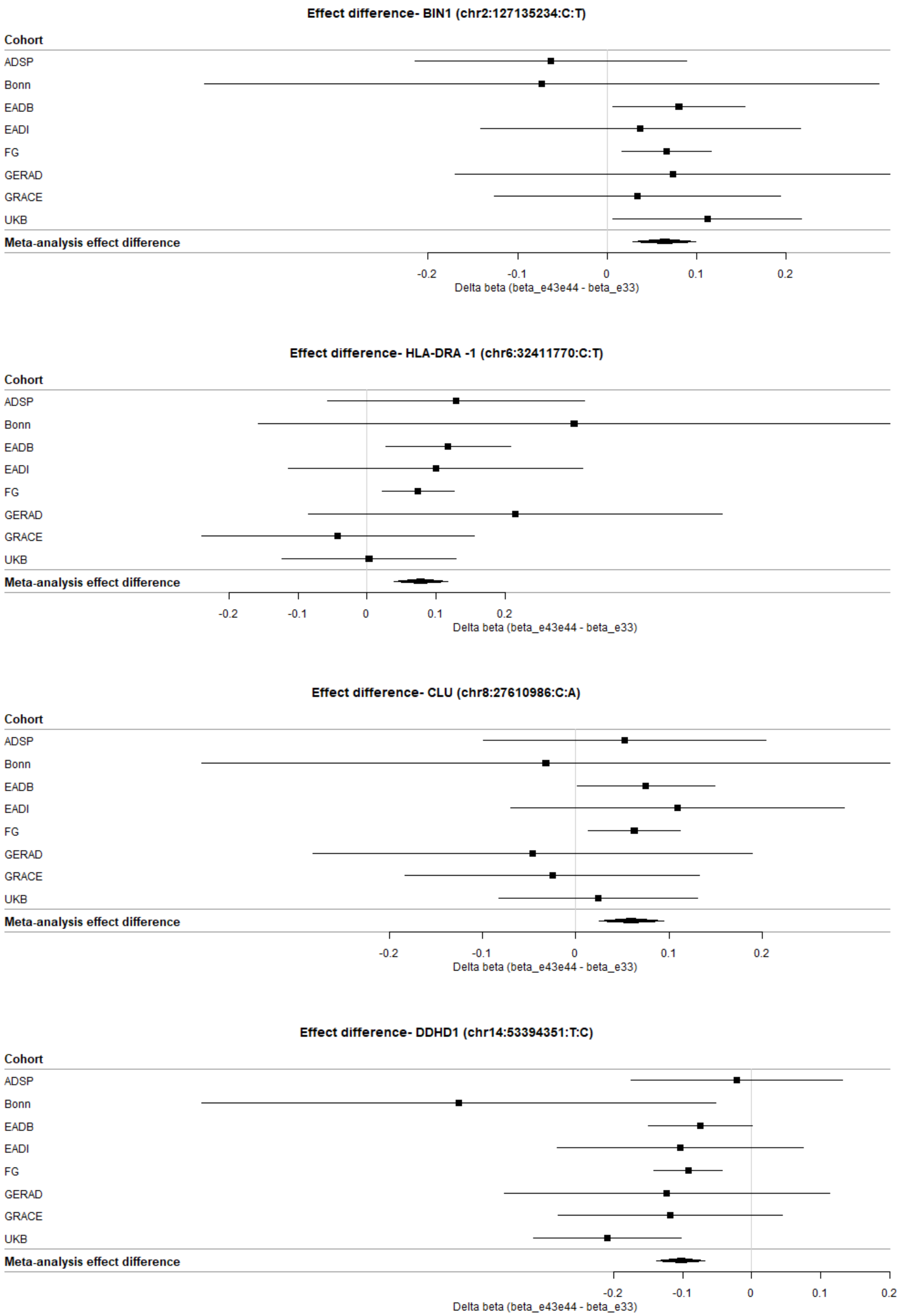


**Supplementary Figure 55: Forest plot for the effect difference between *APOE* strata ε44+ε43 and ε33 for loci with augmented effect with the *APOE* ε4 allele.**

Forest plots for *BIN1*, *HLA-DRA -1*, *CLU* and *DDHD1* of the effect difference between *APOE* strata ε44+ε43 and ε33. Note that the effect difference has been calculated with respect to the TOPMed reference allele. Only loci with dominant interaction are shown.

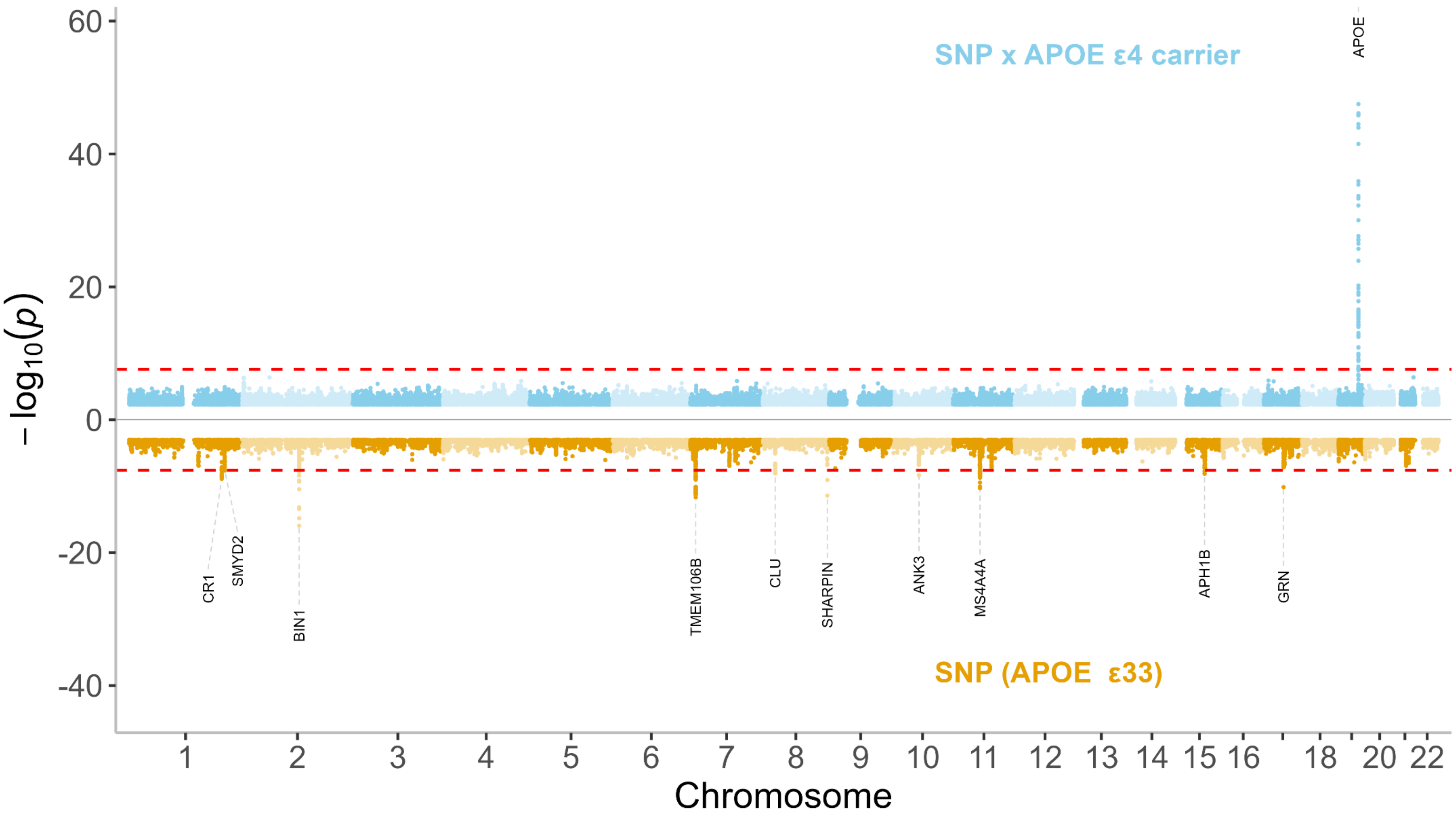


**Supplementary figure 56: Miami plot of the SNP term and interaction terms in the interaction model.**

The Manhattan plot for the interaction term (SNP x *APOE* ε4 carrier) is shown in light blue in the upper part of the figure and for the SNP term (the reference level of the interaction model corresponding to the *APOE* ε33 strata) in orange in the lower part of the figure. Genome wide significant loci are annotated with the nearest gene. Two-sided raw P-values were derived from a fixed-effect meta-analysis. The red dashed lines show the genome-wide significant level ($P=2.5\times10^{-8}$). *APOE*: Apolipoprotein E gene.

### Supplementary Figure 57

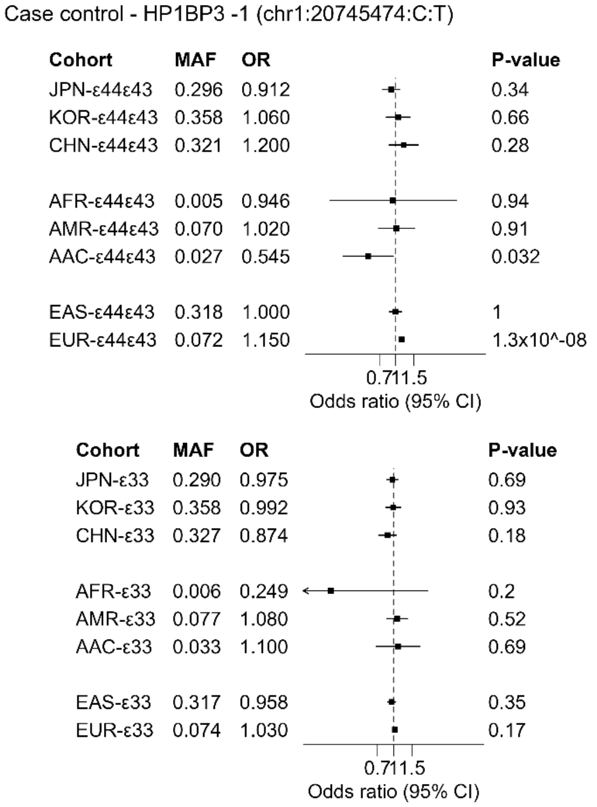


**Supplementary Figure 57: Evaluation of *HP1BP3* SNP across cohorts of different ancestry.**

Forest plots for the lead SNP for *HP1BP3* across cohorts of different ancestry and fixed-effect meta-analysis estimates of the East Asian and European cohorts respectively in the two *APOE* strata ε44+ε43 and ε33. AAC: American Asian, AFR: American African, AMR: Admixed American, CHN: Hong Kong Chinese, EAS: East Asian, EUR: European, JPN: Japanese, KOR: Korean.

### Supplementary Figure 58

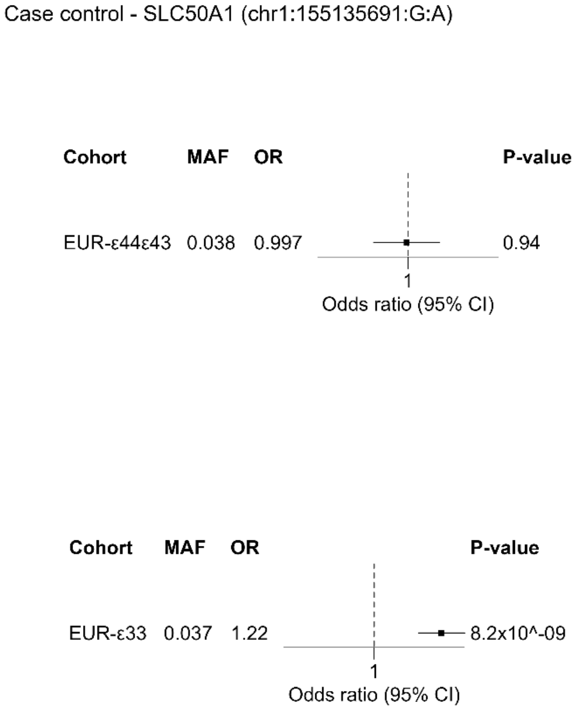


**Supplementary Figure 58: Evaluation of *SLC50A1* SNP across cohorts of different ancestry.**

Forest plots for the lead SNP for *SLC50A1* across cohorts of different ancestry and fixed-effect meta-analysis estimates of the East Asian and European cohorts respectively in the two *APOE* strata ε44+ε43 and ε33. AAC: American Asian, AFR: American African, AMR: Admixed American, CHN: Hong Kong Chinese, EAS: East Asian, EUR: European, JPN: Japanese, KOR: Korean.

### Supplementary Figure 59

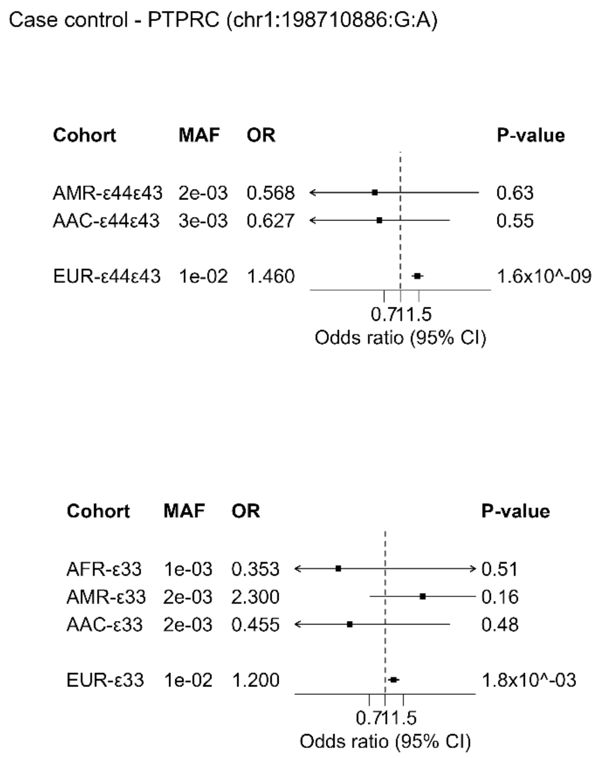


**Supplementary Figure 59: Evaluation of *PTPRC* SNP across cohorts of different ancestry.**

Forest plots for the lead SNP for *PTPRC* across cohorts of different ancestry and fixed-effect meta-analysis estimates of the East Asian and European cohorts respectively in the two *APOE* strata ε44+ε43 and ε33. AAC: American Asian, AFR: American African, AMR: Admixed American, CHN: Hong Kong Chinese, EAS: East Asian, EUR: European, JPN: Japanese, KOR: Korean.

**Supplementary Figure 60**

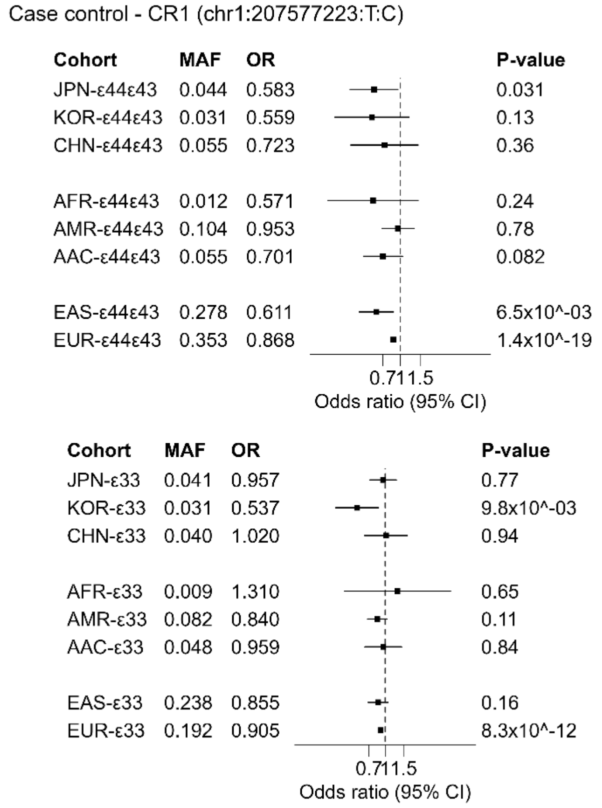


**Supplementary Figure 60: Evaluation of *CR1* SNP across cohorts of different ancestry.**

Forest plots for the lead SNP for *CR1* across cohorts of different ancestry and fixed-effect meta-analysis estimates of the East Asian and European cohorts respectively in the two *APOE* strata ε44+ε43 and ε33. AAC: American Asian, AFR: American African, AMR: Admixed American, CHN: Hong Kong Chinese, EAS: East Asian, EUR: European, JPN: Japanese, KOR: Korean.

**Supplementary Figure 61**

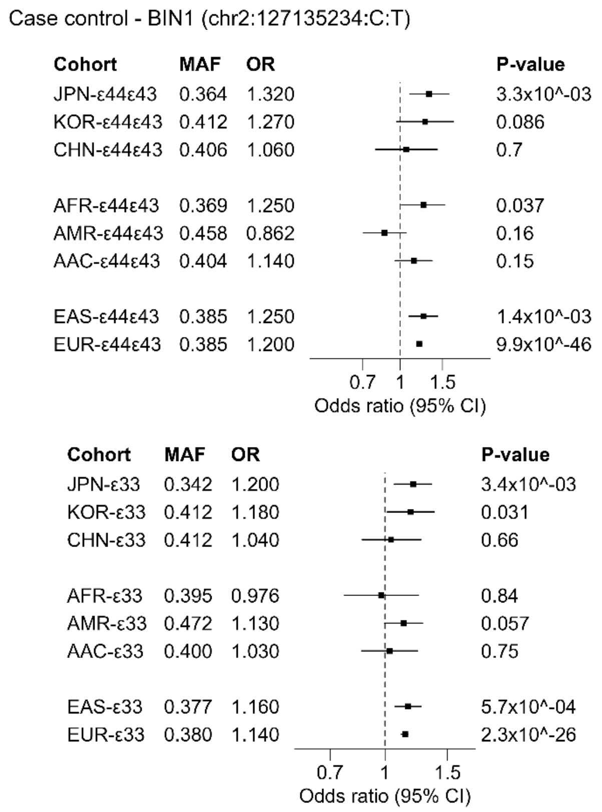


**Supplementary Figure 61: Evaluation of *BIN1* SNP across cohorts of different ancestry.**

Forest plots for the lead SNP for *BIN1* across cohorts of different ancestry and fixed-effect meta-analysis estimates of the East Asian and European cohorts respectively in the two *APOE* strata ε44+ε43 and ε33. AAC: American Asian, AFR: American African, AMR: Admixed American, CHN: Hong Kong Chinese, EAS: East Asian, EUR: European, JPN: Japanese, KOR: Korean.

### Supplementary Figure 62

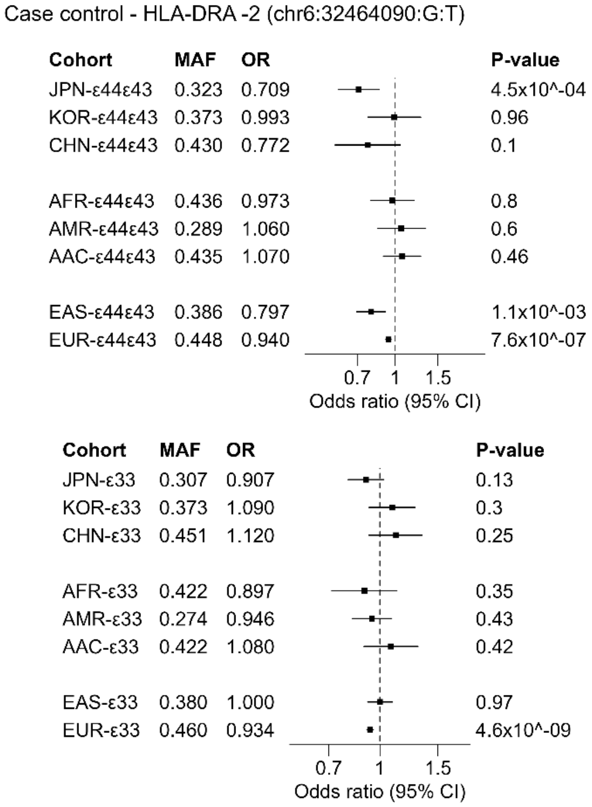


**Supplementary Figure 62: Evaluation of *HLA-DRA -2* SNP across cohorts of different ancestry.**

Forest plots for the lead SNP for *HLA-DRA -2* across cohorts of different ancestry and fixed-effect meta-analysis estimates of the East Asian and European cohorts respectively in the two *APOE* strata ε44+ε43 and ε33. AAC: American Asian, AFR: American African, AMR: Admixed American, CHN: Hong Kong Chinese, EAS: East Asian, EUR: European, JPN: Japanese, KOR: Korean.

### Supplementary Figure 63

Case control - TREM2 -1 (chr6:41161469:C:T)

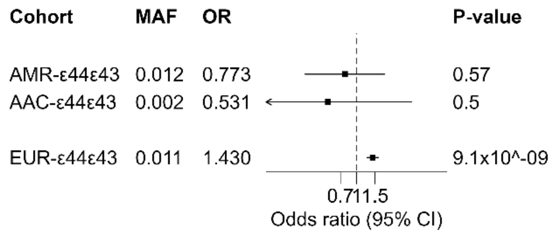


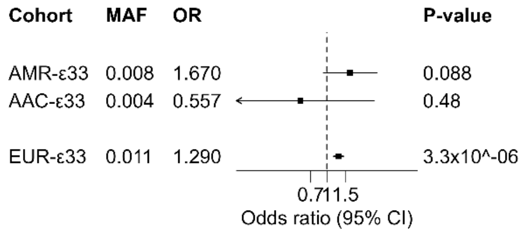


**Supplementary Figure 63: Evaluation of *TREM2 -1* SNP across cohorts of different ancestry.**

Forest plots for the lead SNP for *TREM -1* across cohorts of different ancestry and fixed-effect meta-analysis estimates of the East Asian and European cohorts respectively in the two *APOE* strata ε44+ε43 and ε33. AAC: American Asian, AFR: American African, AMR: Admixed American, CHN: Hong Kong Chinese, EAS: East Asian, EUR: European, JPN: Japanese, KOR: Korean.

### Supplementary Figure 64

Case control - TREM2 -2 (chr6:41161514:C:T)

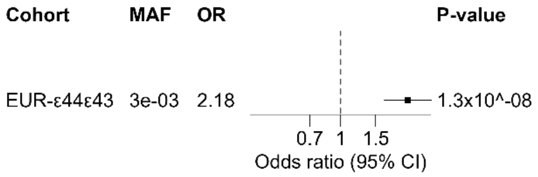


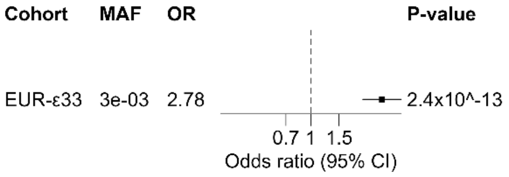


**Supplementary Figure 64: Evaluation of *TREM2 -2* SNP across cohorts of different ancestry.**

Forest plots for the lead SNP for *TREM2 -2* across cohorts of different ancestry and fixed-effect meta-analysis estimates of the East Asian and European cohorts respectively in the two *APOE* strata ε44+ε43 and ε33. AAC: American Asian, AFR: American African, AMR: Admixed American, CHN: Hong Kong Chinese, EAS: East Asian, EUR: European, JPN: Japanese, KOR: Korean.

### Supplementary Figure 65

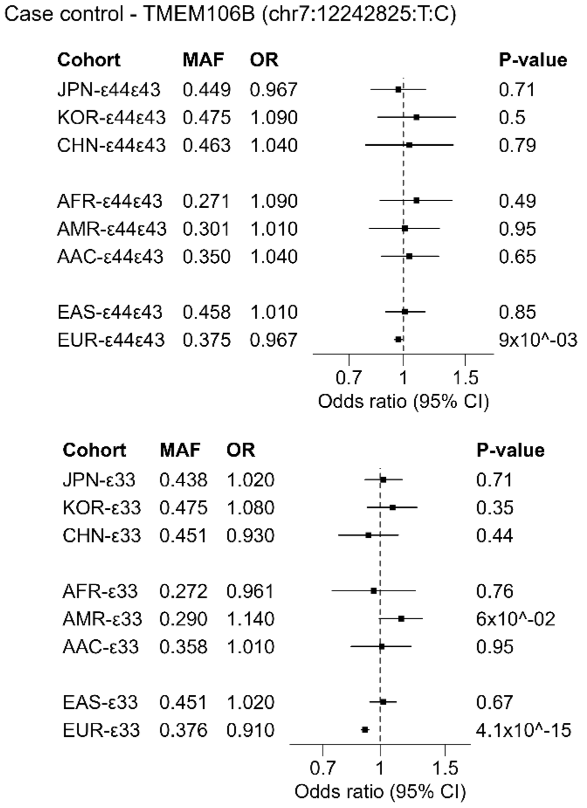


**Supplementary Figure 65: Evaluation of *TMEM106B* SNP across cohorts of different ancestry.**

Forest plots for the lead SNP for *TMEM106B* across cohorts of different ancestry and fixed-effect meta-analysis estimates of the East Asian and European cohorts respectively in the two *APOE* strata ε44+ε43 and ε33. AAC: American Asian, AFR: American African, AMR: Admixed American, CHN: Hong Kong Chinese, EAS: East Asian, EUR: European, JPN: Japanese, KOR: Korean.

### Supplementary Figure 66

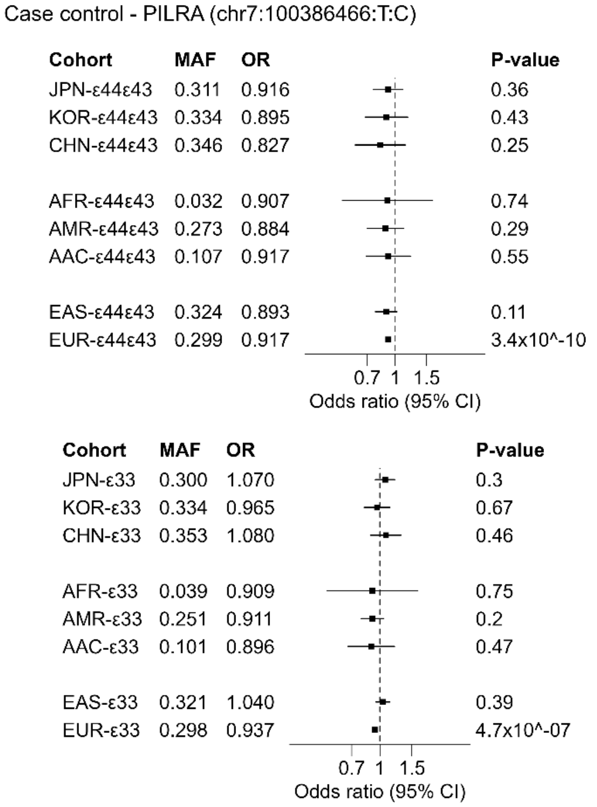


**Supplementary Figure 66: Evaluation of *PILRA* SNP across cohorts of different ancestry.**

Forest plots for the lead SNP for *PILRA* across cohorts of different ancestry and fixed-effect meta-analysis estimates of the East Asian and European cohorts respectively in the two *APOE* strata ε44+ε43 and ε33. AAC: American Asian, AFR: American African, AMR: Admixed American, CHN: Hong Kong Chinese, EAS: East Asian, EUR: European, JPN: Japanese, KOR: Korean.

### Supplementary Figure 67

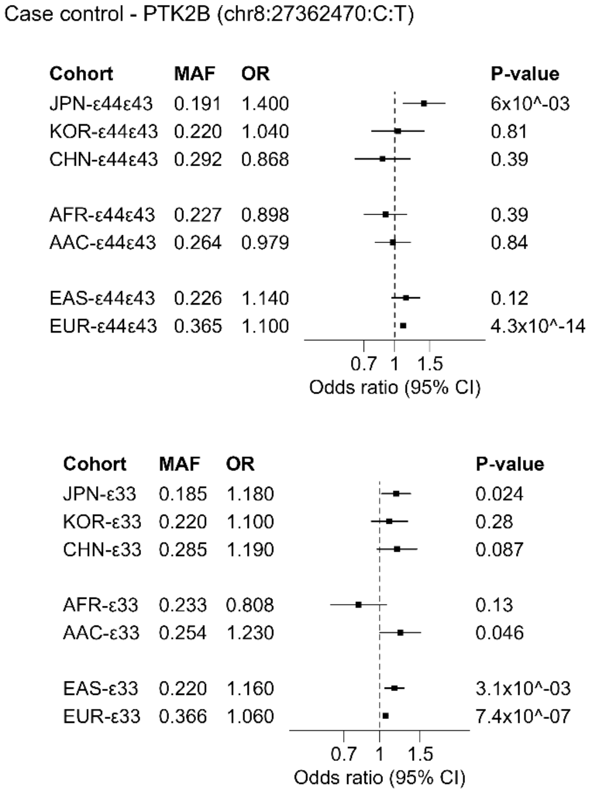


**Supplementary Figure 67: Evaluation of *PTK2B* SNP across cohorts of different ancestry.**

Forest plots for the lead SNP for *PTK2B* across cohorts of different ancestry and fixed-effect meta-analysis estimates of the East Asian and European cohorts respectively in the two *APOE* strata ε44+ε43 and ε33. AAC: American Asian, AFR: American African, AMR: Admixed American, CHN: Hong Kong Chinese, EAS: East Asian, EUR: European, JPN: Japanese, KOR: Korean.

### Supplementary Figure 68

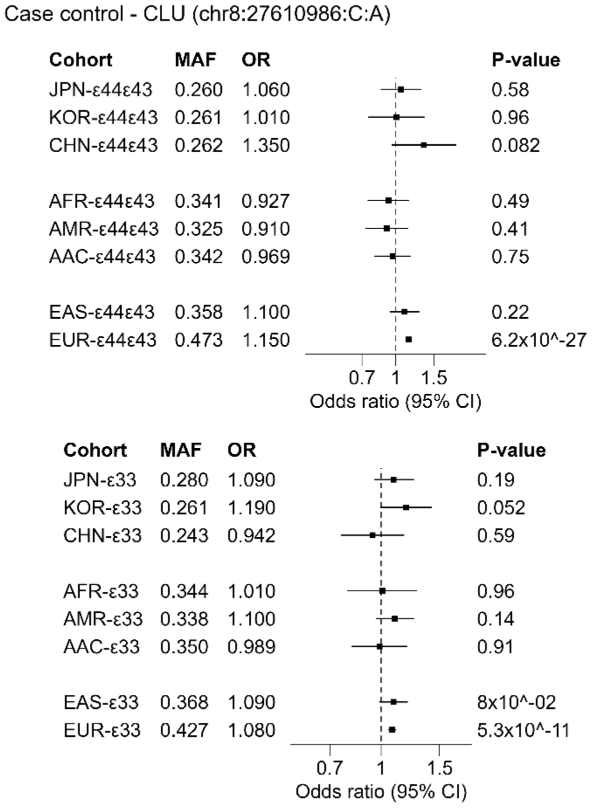


**Supplementary Figure 68: Evaluation of *CLU* SNP across cohorts of different ancestry.**

Forest plots for the lead SNP for *CLU* across cohorts of different ancestry and fixed-effect meta-analysis estimates of the East Asian and European cohorts respectively in the two *APOE* strata ε44+ε43 and ε33. AAC: American Asian, AFR: American African, AMR: Admixed American, CHN: Hong Kong Chinese, EAS: East Asian, EUR: European, JPN: Japanese, KOR: Korean.

### Supplementary Figure 69

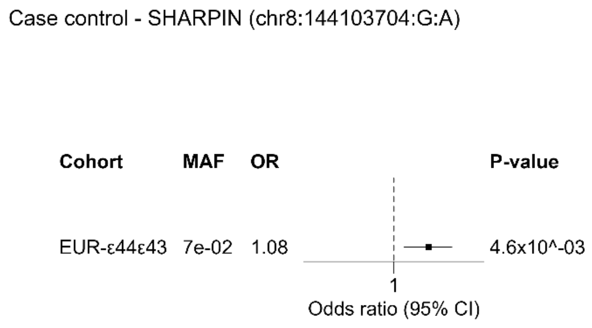


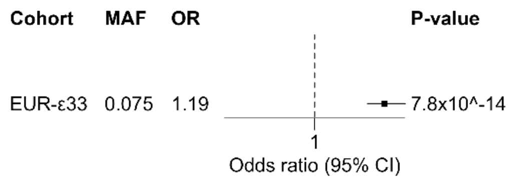


**Supplementary Figure 69: Evaluation of *SHARPIN* SNP across cohorts of different ancestry.**

Forest plots for the lead SNP for *SHARPIN* across cohorts of different ancestry and fixed-effect meta-analysis estimates of the East Asian and European cohorts respectively in the two *APOE* strata ε44+ε43 and ε33. AAC: American Asian, AFR: American African, AMR: Admixed American, CHN: Hong Kong Chinese, EAS: East Asian, EUR: European, JPN: Japanese, KOR: Korean.

### Supplementary Figure 70

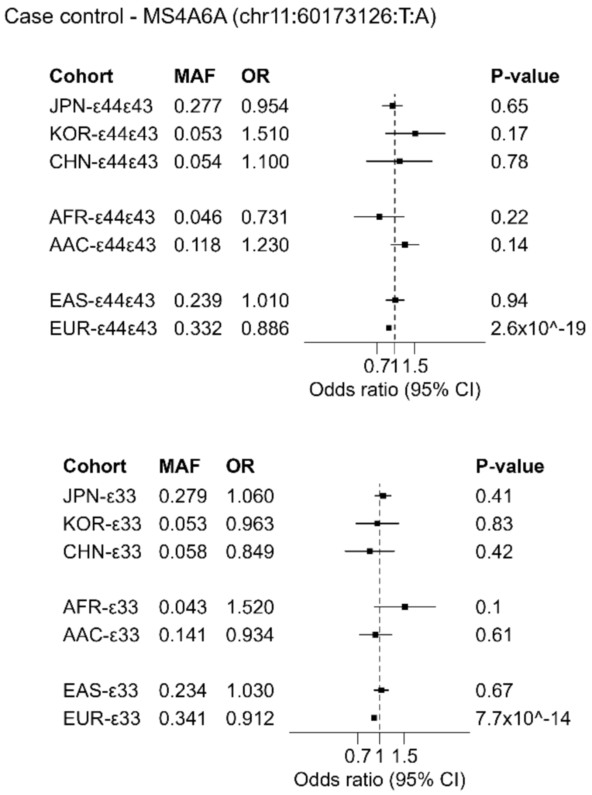


**Supplementary Figure 70: Evaluation of *MS4A6A* SNP across cohorts of different ancestry.**

Forest plots for the lead SNP for *MS4A6A* across cohorts of different ancestry and fixed-effect meta-analysis estimates of the East Asian and European cohorts respectively in the two *APOE* strata ε44+ε43 and ε33. AAC: American Asian, AFR: American African, AMR: Admixed American, CHN: Hong Kong Chinese, EAS: East Asian, EUR: European, JPN: Japanese, KOR: Korean.

**Supplementary Figure 71**

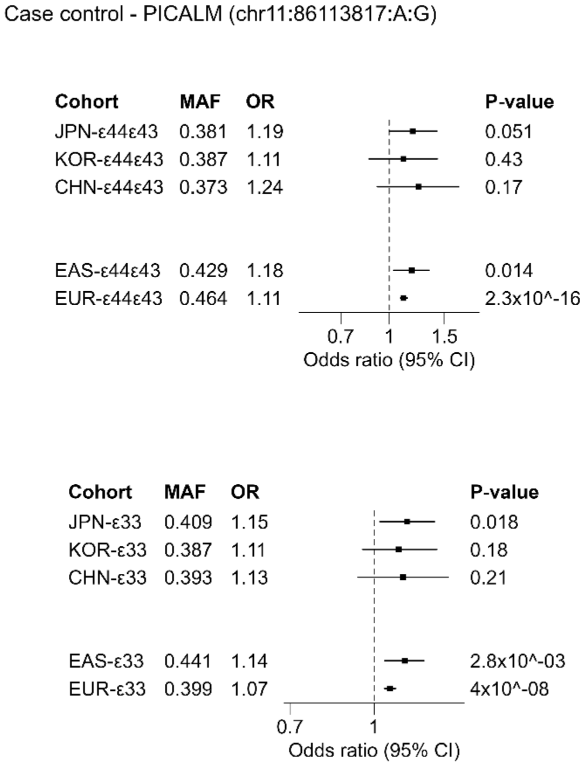


**Supplementary Figure 71: Evaluation of *PICALM* SNP across cohorts of different ancestry.**

Forest plots for the lead SNP for *PICALM* across cohorts of different ancestry and fixed-effect meta-analysis estimates of the East Asian and European cohorts respectively in the two *APOE* strata ε44+ε43 and ε33. AAC: American Asian, AFR: American African, AMR: Admixed American, CHN: Hong Kong Chinese, EAS: East Asian, EUR: European, JPN: Japanese, KOR: Korean.

### Supplementary Figure 72

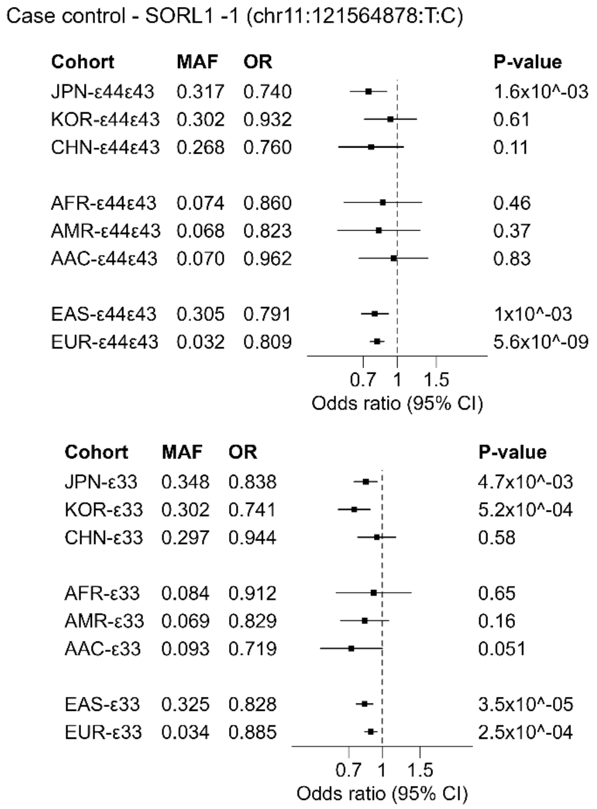


**Supplementary Figure 72: Evaluation of *SORL1* SNP across cohorts of different ancestry.**

Forest plots for the lead SNP for *SORL1* across cohorts of different ancestry and fixed-effect meta-analysis estimates of the East Asian and European cohorts respectively in the two *APOE* strata ε44+ε43 and ε33. AAC: American Asian, AFR: American African, AMR: Admixed American, CHN: Hong Kong Chinese, EAS: East Asian, EUR: European, JPN: Japanese, KOR: Korean.

### Supplementary Figure 73

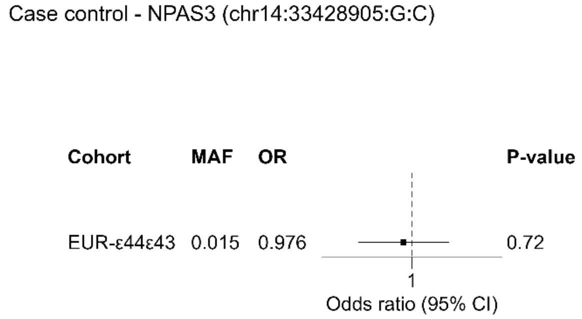


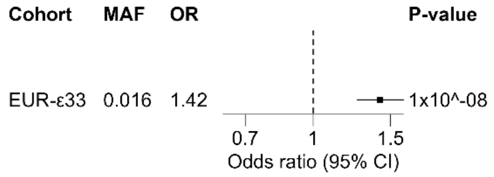


**Supplementary Figure 73: Evaluation of *NPAS3* SNP across cohorts of different ancestry.**

Forest plots for the lead SNP for *NPAS3* across cohorts of different ancestry and fixed-effect meta-analysis estimates of the East Asian and European cohorts respectively in the two *APOE* strata ε44+ε43 and ε33. AAC: American Asian, AFR: American African, AMR: Admixed American, CHN: Hong Kong Chinese, EAS: East Asian, EUR: European, JPN: Japanese, KOR: Korean.

### Supplementary Figure 74

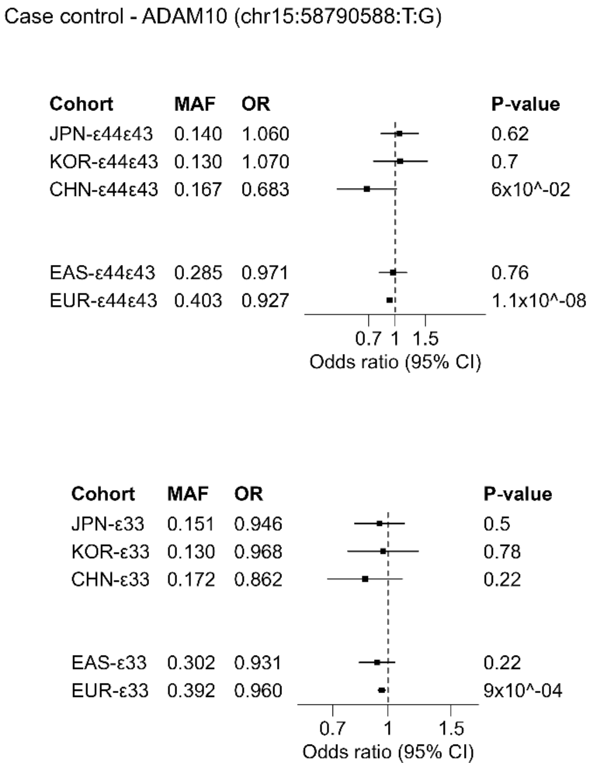


**Supplementary Figure 74: Evaluation of *ADAM10* SNP across cohorts of different ancestry.**

Forest plots for the lead SNP for *ADAM10* across cohorts of different ancestry and fixed-effect meta-analysis estimates of the East Asian and European cohorts respectively in the two *APOE* strata ε44+ε43 and ε33. AAC: American Asian, AFR: American African, AMR: Admixed American, CHN: Hong Kong Chinese, EAS: East Asian, EUR: European, JPN: Japanese, KOR: Korean.

**Supplementary Figure 75**

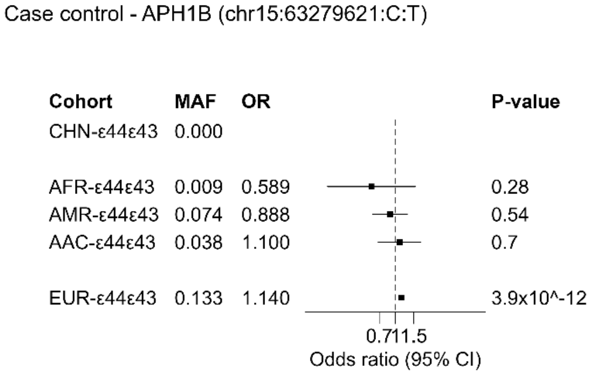


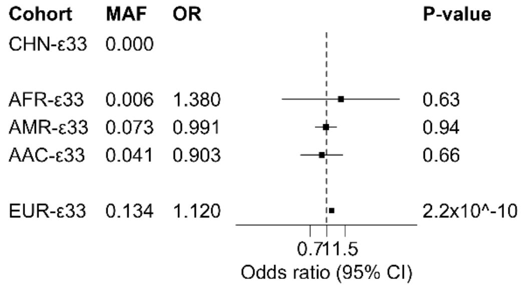


**Supplementary Figure 75: Evaluation of *APH1B* SNP across cohorts of different ancestry.**

Forest plots for the lead SNP for *APH1B* across cohorts of different ancestry and fixed-effect meta-analysis estimates of the East Asian and European cohorts respectively in the two *APOE* strata ε44+ε43 and ε33. AAC: American Asian, AFR: American African, AMR: Admixed American, CHN: Hong Kong Chinese, EAS: East Asian, EUR: European, JPN: Japanese, KOR: Korean.

### Supplementary Figure 76

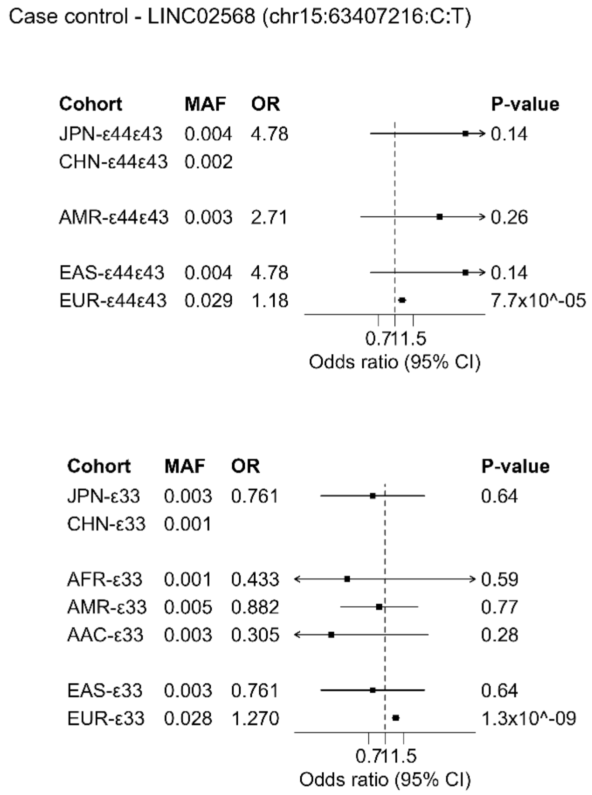


**Supplementary Figure 76: Evaluation of *LINC02568* SNP across cohorts of different ancestry.**

Forest plots for the lead SNP for *LINC02568* across cohorts of different ancestry and fixed-effect meta-analysis estimates of the East Asian and European cohorts respectively in the two *APOE* strata ε44+ε43 and ε33. AAC: American Asian, AFR: American African, AMR: Admixed American, CHN: Hong Kong Chinese, EAS: East Asian, EUR: European, JPN: Japanese, KOR: Korean.

### Supplementary Figure 77

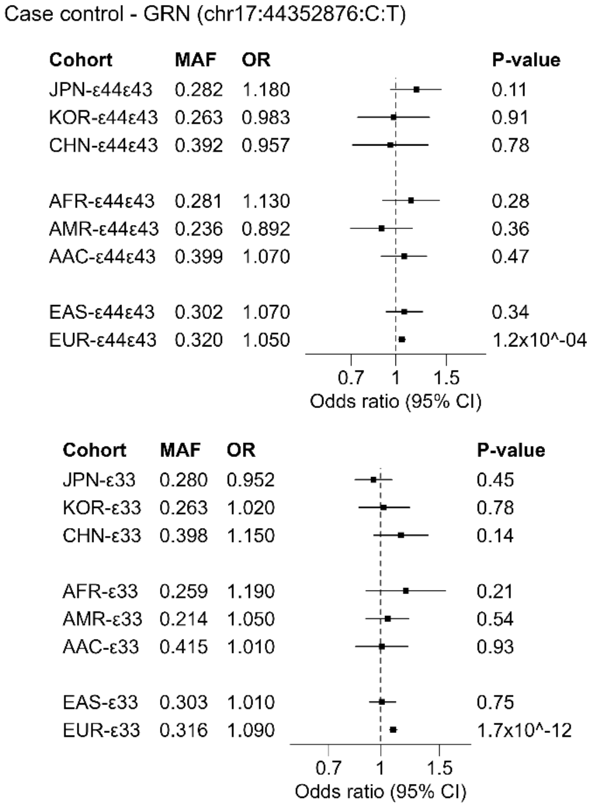


**Supplementary Figure 77: Evaluation of *GRN* SNP across cohorts of different ancestry.**

Forest plots for the lead SNP for *GRN* across cohorts of different ancestry and fixed-effect meta-analysis estimates of the East Asian and European cohorts respectively in the two *APOE* strata ε44+ε43 and ε33. AAC: American Asian, AFR: American African, AMR: Admixed American, CHN: Hong Kong Chinese, EAS: East Asian, EUR: European, JPN: Japanese, KOR: Korean.

**Supplementary Figure 78**

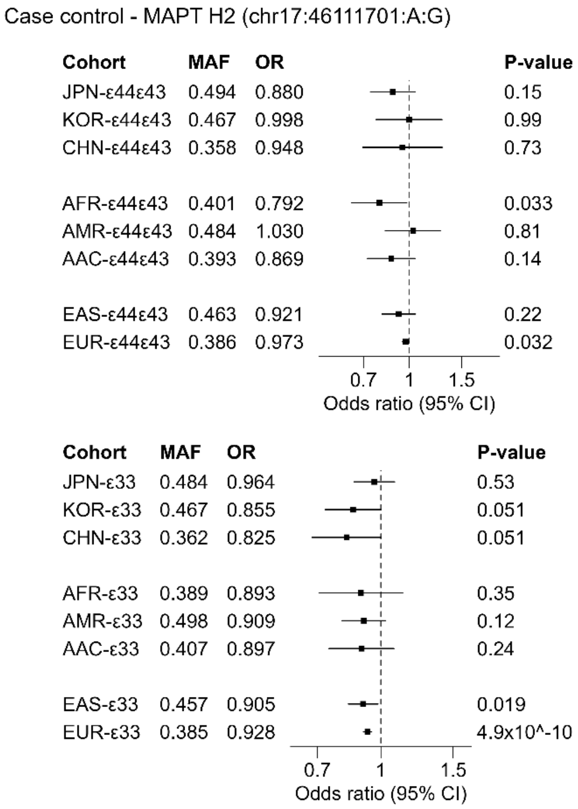


**Supplementary Figure 78: Evaluation of *MAPT* SNP across cohorts of different ancestry.**

Forest plots for the lead SNP for *MAPT* across cohorts of different ancestry and fixed-effect meta-analysis estimates of the East Asian and European cohorts respectively in the two *APOE* strata ε44+ε43 and ε33. AAC: American Asian, AFR: American African, AMR: Admixed American, CHN: Hong Kong Chinese, EAS: East Asian, EUR: European, JPN: Japanese, KOR: Korean.

### Supplementary Figure 79

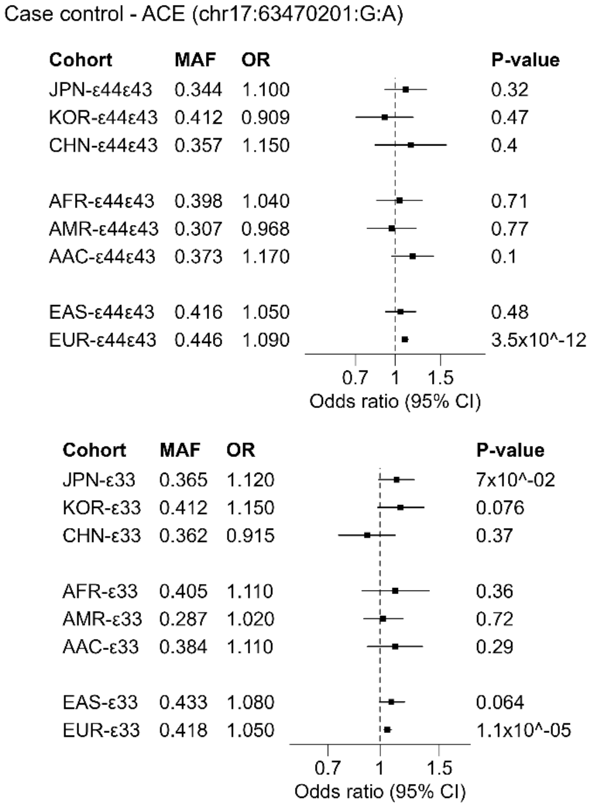


**Supplementary Figure 79: Evaluation of *ACE* SNP across cohorts of different ancestry.**

Forest plots for the lead SNP for *ACE* across cohorts of different ancestry and fixed-effect meta-analysis estimates of the East Asian and European cohorts respectively in the two *APOE* strata ε44+ε43 and ε33. AAC: American Asian, AFR: American African, AMR: Admixed American, CHN: Hong Kong Chinese, EAS: East Asian, EUR: European, JPN: Japanese, KOR: Korean.

### Supplementary Figure 80

Case control - CHST9 (chr18:27352028:C:A)

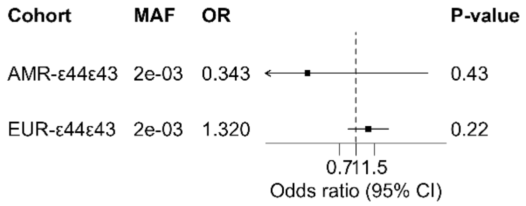


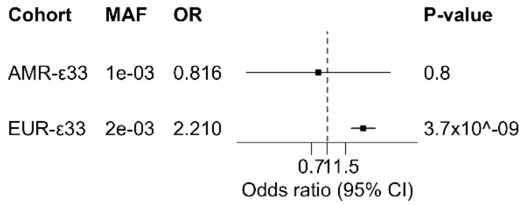


**Supplementary Figure 80: Evaluation of *CHST9* SNP across cohorts of different ancestry.**

Forest plots for the lead SNP for *CHST9* across cohorts of different ancestry and fixed-effect meta-analysis estimates of the East Asian and European cohorts respectively in the two *APOE* strata ε44+ε43 and ε33. AAC: American Asian, AFR: American African, AMR: Admixed American, CHN: Hong Kong Chinese, EAS: East Asian, EUR: European, JPN: Japanese, KOR: Korean.

### Supplementary Figure 81

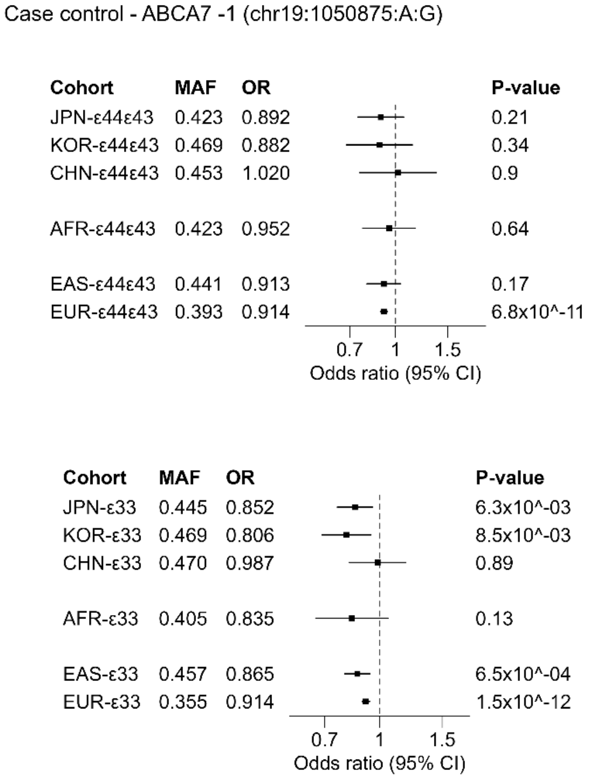


**Supplementary Figure 81: Evaluation of *ABCA7* SNP across cohorts of different ancestry.**

Forest plots for the lead SNP for *ABCA7* across cohorts of different ancestry and fixed-effect meta-analysis estimates of the East Asian and European cohorts respectively in the two *APOE* strata ε44+ε43 and ε33. AAC: American Asian, AFR: American African, AMR: Admixed American, CHN: Hong Kong Chinese, EAS: East Asian, EUR: European, JPN: Japanese, KOR: Korean.

### Supplementary Figure 82

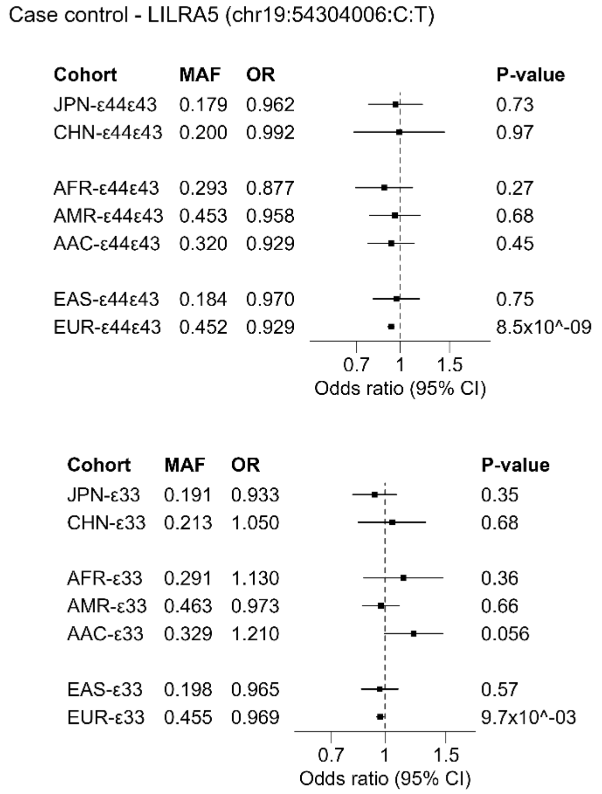


**Supplementary Figure 82: Evaluation of *LILRA5* SNP across cohorts of different ancestry.**

Forest plots for the lead SNP for *LILRA5* across cohorts of different ancestry and fixed-effect meta-analysis estimates of the East Asian and European cohorts respectively in the two *APOE* strata ε44+ε43 and ε33. AAC: American Asian, AFR: American African, AMR: Admixed American, CHN: Hong Kong Chinese, EAS: East Asian, EUR: European, JPN: Japanese, KOR: Korean.

**Supplementary Figure 83**

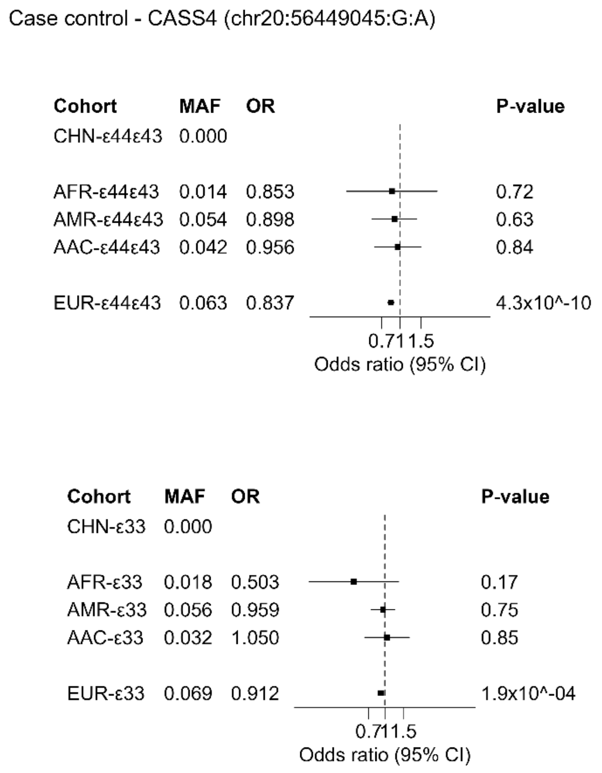


**Supplementary Figure 83: Evaluation of *CASS4* SNP across cohorts of different ancestry.**

Forest plots for the lead SNP for *CASS4* across cohorts of different ancestry and fixed-effect meta-analysis estimates of the East Asian and European cohorts respectively in the two *APOE* strata ε44+ε43 and ε33. AAC: American Asian, AFR: American African, AMR: Admixed American, CHN: Hong Kong Chinese, EAS: East Asian, EUR: European, JPN: Japanese, KOR: Korean.

### Supplementary Figure 84

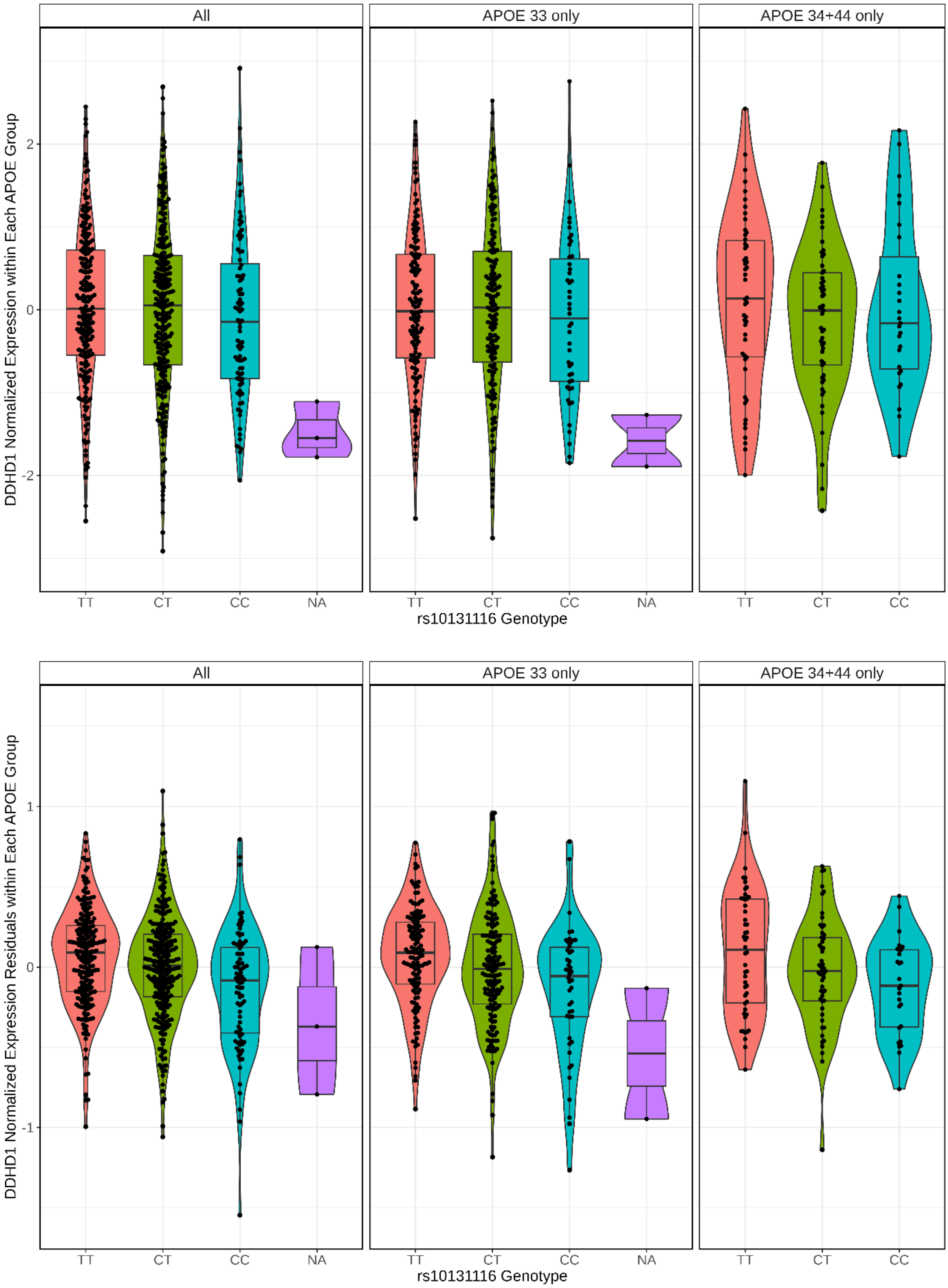


**Supplementary Figure 84: rs10131116 association with *DDHD1* expression**

eQTL violin plots showing rs10131116 association with normalized *DDHD1* expression (top) and residuals of normalized expressions of *DDHD1* in the ROSMAP DLPFC data set shown in full dataset (left), and *APOE* ε33 strata (middle) and *APOE* ε44+ε43 strata (right).
