## Supplementary note for "*APOE* stratified genome-wide association studies provide novel insights into the genetic etiology of Alzheimers’s disease"

### 1. GWAS Cohort descriptions and acknowledgements

#### 1.1 European Alzheimer & Dementia Biobank (EADB-TOPMed)

This consortium groups together 20,464 Alzheimer's disease (AD) cases and 22,244 controls after quality controls from 16 European countries (Austria, Belgium, Bulgaria, Czech Republic, Denmark, Finland, France, Germany, Greece, Italy, Portugal, Spain, Sweden, Switzerland, The Netherlands and the UK). These samples were genotyped in three independent centers (France, Germany and the Netherlands) leading to define three nodes: EADB-France, EADB-Germany and EADB-Netherlands. In addition, EADB also included Australian partners.

We thank the numerous participants, researchers, and staff from many studies who collected and contributed to the data. We thank the high performance computing service of the University of Lille. We thank all the CEA-CNRGH staff who contributed to sample preparation and genotyping for their excellent technical assistance.

This work was supported by a grant (European Alzheimer DNA BioBank, EADB) from the EU Joint Programme – Neurodegenerative Disease Research (JPND). Inserm UMR1167 is also funded by Inserm, Institut Pasteur de Lille, the Lille Métropole Communauté Urbaine, the French government's LABEX DISTALZ program (development of innovative strategies for a transdisciplinary approach to Alzheimer's disease).

**EADB-France.** In the France node, samples were collected from nine countries (39 centers/studies), and after quality controls (QCs), we obtained 13,867 AD cases and 15,310 controls. All these samples were genotyped at the Centre National de Recherche en Génomique Humaine (CNRGH, Evry, France).
<u>Belgium:</u> The participants were part of a large prospective cohort (PMID: 29589097) of Belgian AD patients and healthy elderly control individuals. The patients were ascertained at the memory clinic of Middelheim and Hoge Beuken (Hospital Network Antwerp, Belgium) and at the memory clinic of the University Hospitals of Leuven, Belgium. The control individuals were the partners of the patients or volunteers from the Belgian community. The study protocols were approved by the ethics committees of the Antwerp University Hospital and the participating neurological centers at the different hospitals of the BELNEU consortium and by the University of Antwerp.
<u>Czech Republic:</u> The Czech Brain Aging Study (CBAS) (PMID: 31857299) is a longitudinal memory-clinic–based study recruiting subjects at risk of dementia (subjects referred for cognitive complaints-SCD, MCI). The CBAS+ study is a cross-sectional study of patients in the early stages of dementia. All subjects signed informed consent and both studies were approved by the local ethics committee.
<u>Denmark:</u> The Copenhagen General Population Study (CGPS) is a prospective study of the Danish general population initiated in 2003 and still recruiting. Individuals were selected randomly based on the national Danish Civil Registration System to reflect the adult Danish population aged 20-100. Data were obtained from a self-administered questionnaire reviewed together with an investigator at the day of attendance, a physical examination, and from blood samples including DNA extraction (PMID: 33022702). The study is approved by the regional ethics committee and by the Steering Committee of the CGPS.
<u>Finland:</u> *The ADGEN cohort (PMID: 25807283):* clinic-based collection of AD patients from Eastern and Northern Finland examined in the Department of Neurology in Kuopio University Hospital and the Department of Neurology in Oulu University Hospital. All the patients were diagnosed with probable AD according to the criteria of the National Institute of Neurological and Communicative Disorders and Stroke and the Alzheimer's disease and Related Disorders Association (NINCDS–ADRDA). The study was approved by the ethics committee of Kuopio University Hospital, Finland (420/2016). *The FINGER study* (PMID: 25771249): a Finnish multi-domain lifestyle RCT enrolling 1,260 older adults with an increased risk of dementia from the general population. The intensive lifestyle intervention lasted for two years, and follow-up extends currently up to seven years. The FINGER study was approved by the coordinating ethics committee of the Hospital

District of Helsinki and Uusimaa (94/13/03/00/2009 and HUS/1204/2017), and all the participants gave written informed consent.

France: *The BALTAZAR multicenter (23 memory centers) prospective study* (PMID: 29458036): 1,040 participants from September 2010 to April 2015. They were classified as AD cases (n = 501) according to DSM IV-TR and NINCDS–ADRDA criteria as well as amnestic mild cognitive impairment (MCI) cases (a MCI, n = 417) and non-amnestic MCI cases (na MCI, n = 122) according to Petersen's criteria. A comprehensive battery of cognitive tests was performed, including MMSE, verbal fluency, and FCSRT. All the participants or their legal guardians gave written informed consent. The study was approved by the Paris ethics committee (CPP Ile de France IV Saint Louis Hospital). MEMENTO: a clinic-based study (PMID: 28851447) aimed at better understanding the natural history of AD, dementia, and related diseases. Between 2011 and 2014, 2,323 individuals presenting either recently diagnosed MCI or isolated cognitive complaints were enrolled in 26 memory centers in France. This study was performed in accordance with the guidelines of the Declaration of Helsinki. The MEMENTO study protocol has been approved by the local ethics committee (Comité de Protection des Personnes Sud-Ouest et Outre Mer III; approval number 2010-A01394-35). All the participants provided written informed consent. The CNRMAJ-Rouen study (PMID: 26242991): early onset AD patients (n = 870). The patients or their legal guardians provided written informed consent. This study was approved by the ethics committee of CPP Ile de France II.

Italy: The AD cases and controls were collated through Italy in different centers: Brescia, Cagliari, Florence, Milan, Rome, Pertugia, San Giovani Rotondo and Torino. AD cases were diagnosed according to DSM III-R,IV and NINCDS–ADRDA criteria. Controls were defined a minima as subjects without DMS-III-R dementia criteria and with integrity of their cognitive functions (MMS>25).

Spain: *The Dementia Genetic Spanish Consortium (DEGESCO)* is a national consortium comprising 23 research centers and hospitals across the country, that holds the institutional coverage of The Network Center for Biomedical Research in Neurodegenerative Diseases (CIBERNED). Created in 2013, DEGESCO's objective is the promotion and conduction of genetic studies aimed at understanding the genetic architecture of neurodegenerative dementias in the Spanish population and participates in coordinated actions in national and international frameworks. All DNA samples are in compliance with the Law of Biomedical Research (Law 14/2007) and the Royal Decree on Biobanks (RD 1716/2011). Patients included in the present study met clinical criteria for probable or possible disease established by the National Institute of Neurological and Communication Disorders and Stroke and the Alzheimer Disease and Related Disorders Association (NINCDS-ADRDA). Cognitively healthy controls were unrelated individuals who had a documented MMSE in the normal range. Contributing centers in the France node genotyping were Centro de Biología Molecular Severo Ochoa (CSIC-UAM (Madrid), the Institute Biodonostia, University of Basque Contry (EHU-UPV, San Sebastián), Institut de Biomedicina de Valencia CSIC (València), and Sant Pau Biomedical Research Institute (Barcelona).

Sweden: *Uppsala.* The Swedish AD patients were ascertained at the Memory Disorder Unit at Uppsala University Hospital. For all patients, the diagnosis was established according to the National Institute on Neurological Disorders and Stroke, and the Alzheimer's Disease and Related Disorders Association (NINDS-ADRDA) guidelines (PMID: 6610841). Healthy control subjects were recruited from the same geographic region following advertisements in local newspapers and displayed no signs of dementia upon Mini Mental State Examination (MMSE). *Karolinska Institutet, Stockholm*. AD patients were ascertained at the Memory Clinic at Karolinska University Hospital - Huddinge, Stockholm Sweden. For all patients, the diagnosis was established according to the National Institute on Neurological Disorders and Stroke, and the Alzheimer's Disease and Related Disorders Association (NINDS-ADRDA) guidelines (PMID: 6610841) and if available supported by CSF beta-amyloid and phospho-tau biomarker analyses. As controls, DNA from participants in the *Swedish National Study on Aging and Care in Kungsholmen* (SNAC-K) data was collected. The original SNAC-K population consisted of 4590 living and eligible persons who lived on the island of Kungsholmen in Central Stockholm, belonged to pre-specified age strata, and were randomly selected to take part in the study. Between 2001 and 2004, 3363 persons participated in the baseline assessment. They belonged to the age cohorts 60, 66, 72, 78, 81, 84, 87, 90, 93, and 96 years and 99 years and older. The examination consists of three parts: a nurse interview, a medical examination, and a neuropsychological testing session. Altogether, the examination takes about six hours. The participants are reexamined each time they reach the next age cohort. All parts of the SNAC-K project have been approved by the ethical committee at Karolinska

Institutet or the regional ethical review board. Informed consent was collected from all the participants or, if the person was severely cognitively impaired, from their next of kin.
The UK: MRC. The sample set comprises individuals with AD and healthy controls recruited across the MRC Centre for Neuropsychiatric Genetics and Genomics, Cardiff University, Cardiff, UK; Institute of Psychiatry, London, UK; University of Cambridge, Cambridge, UK. The collection of the samples was through multiple channels, including specialist NHS services and clinics, research registers and Join Dementia Research (JDR) platform. The participants were assessed at home or in research clinics along with an informant, usually a spouse, family member or close friend, who provided information about and on behalf of the individual with dementia. Established measures were used to ascertain the disease severity: Bristol activities of daily living (BADL), Clinical Dementia Rating scale (CDR), Neuropsychiatric Inventory (NPI) and Global Deterioration Scale (GlDS). Individuals with dementia completed the Addenbrooke's Cognitive Examination (ACE-r), Geriatric Depression Scale (GeDS) and National Adult Reading Test (NART) too. Control participants were recruited from GP surgeries and by means of self-referral (including existing studies and Joint Dementia Research platform). For all other recruitment, all AD cases met criteria for either probable (NINCDS-ADRDA, DSM-IV) or definite (CERAD) AD. All elderly controls were screened for dementia using the Mini Mental State Examination (MMSE) or ADAS-cog, were determined to be free from dementia at neuropathological examination or had a Braak score of 2.5 or lower. Control samples were chosen to match case samples for age, gender, ethnicity and country of origin. Informed consent was obtained for all study participants, and the relevant independent ethical committees approved study protocols. SOTON, University of Southampton, Southampton, UK. All AD cases met criteria for either probable (NINCDS-ADRDA, DSM-IV) or definite (CERAD) AD. All elderly controls were screened for dementia using the MMSE or ADAS-cog, were determined to be free from dementia at neuropathological examination or had a Braak score of 2.5 or lower. Nottingham and Manchester, University of Nottingham, Nottingham, UK and Manchester Brain Bank. All AD cases met criteria for either probable (NINCDS-ADRDA, DSM-IV) or definite (CERAD) AD. All elderly controls were screened for dementia using the MMSE or ADAS-cog, were determined to be free from dementia at neuropathological examination or had a Braak score of 2.5 or lower. KCL, London Neurodegenerative Diseases Brain Bank. All AD cases met criteria for either probable (NINCDS-ADRDA, DSM-IV) or definite (CERAD) AD. All elderly controls were screened for dementia using the MMSE or ADAS-cog, were determined to be free from dementia at neuropathological examination or had a Braak score of 2.5 or lower. PRION, All AD cases met criteria for either probable (NINCDS-ADRDA, DSM-IV) or definite (CERAD) AD. All elderly controls were screened for dementia using the MMSE or ADAS-cog, were determined to be free from dementia at neuropathological examination or had a Braak score of 2.5 or lower. CFAS Wales, The Cognitive Function and Ageing Study Wales (CFAS-Wales) is a longitudinal population-based study of people aged 65 years and over in rural and urban areas of Wales that aims to investigate physical and cognitive health in older age and examine the interactions between health, social networks, activity, and participation. Individuals aged 65 years and over were randomly sampled from general medical practice lists between 2011 and 2013, stratified by age to ensure equal numbers in two age groups, 65-74 years and 75 and over. The baseline sample included 3593 older people and included those living in care homes as well as those living at home. Those who provided written consent to join the study were interviewed in their own homes by trained interviewers and could choose to have the interview conducted through the medium of either English or Welsh. Participants were followed up 2 years later. All AD cases met criteria for either probable (NINCDS-ADRDA, DSM-IV) or definite (CERAD) AD. All elderly controls were screened for dementia using the MMSE or CAMCOG, and were determined to be free from dementia. UCL-DRC. the UCL Alzheimer's disease cohort of the Dementia Research Centre (UCL - EOAD DRC) included patients seen at the Cognitive Disorders Clinics at The National Hospital for Neurology and Neurosurgery (Queen Square), or affiliated hospitals. Individuals were assessed clinically and diagnosed as having probable Alzheimer's disease based on contemporary clinical criteria in use at the time, including imaging and neuropsychological testing where appropriate.
**EADB-Germany.** In the German node, samples were collected from seven countries (11 centers/studies) and after QCs, we obtained 4,159 AD cases and 4,545 controls. All these samples were genotyped at Life&brain (Bonn, Germany).
Germany: *DELCODE (the multicenter DZNE-Longitudinal Cognitive Impairment and Dementia Study)*. This is an observational longitudinal memory clinic-based multicenter study in Germany comprising 400 subjects

with Subjective cognitive decline (SCD), 200 mild cognitive impairment (MCI) patients, 100 AD dementia patients, 200 control subjects without subjective or objective cognitive decline, and 100 first-degree relatives of patients with a documented diagnosis of AD dementia. All patient groups (SCD, MCI, AD) are referrals, including self-referrals, to the participating memory centers. The control group and the relatives of AD dementia patients are recruited by standardized public advertisement. Ten university-based memory centers are participating, all being collaborators of local DZNE sites. All patient groups (SCD, MCI, AD) were assessed clinically at the respective memory centers before entering DELCODE. The assessments include medical history, psychiatric and neurological examination, neuropsychological testing, blood laboratory work-up, cerebrospinal fluid (CSF) biomarkers, and routine MRI, all according to the local standards. The Consortium to Establish a Registry for Alzheimer's Disease (CERAD) neuropsychological test battery was applied at all memory centers to measure cognitive function. German age, sex, and education-adjusted norms of the CERAD neuropsychological battery are available online (www.memoryclinic.ch). Detail description is of recruitment protocol is reported elsewhere. *The VOGEL study*: The VOGEL study is a prospective, observational, long-term follow-up study with three time points of investigation within 6–8 years. This cohort includes dementia and healthy subjects. Residents of the city of Würzburg born between 1936 and 1941 were recruited. Every participant underwent physical, psychiatric, and laboratory examinations and performed intense neuropsychological testing as well as VSEP and NIRS according to the published procedures. A total of 604 subjects were included. *The Heidelberg/Mannheim memory clinic sample*: This cohort includes 61 subjects from whom 40 MCI patients were recruited and assessed between 2012 and 2016. Some of those patients converted to dementia by AD or other dementias. *The PAGES study*: This study includes 301 subjects. AD patients were recruited at the memory clinic of the Department of Psychiatry, University of Munich, Germany. Participants in whom dementia associated with AD was diagnosed fulfilled the criteria for probable AD according to the NINCDS–ADRDA. The control group included participants who were randomly selected from the general population of Munich. Controls who had central nervous system diseases or psychotic disorders or who had first-degree relatives with psychotic disorders were excluded. *The Technische Universität München study*: This cohort includes 359 healthy, AD, and other dementias patients recruited from the Centre for Cognitive Disorders. All the participants provided written informed consent. A biobank was submitted to the ethics committee of the Technical University of Munich, School of Medicine (Munich, Germany), which raised no objections and approved the biobank (reference number 347-14*). The Göttingen Universität study*: This study includes 111 in- and outpatients with a healthy or AD dementia status from the Department of Psychiatry of the University of Göttingen. The study's ethical statement was provided locally at the Göttingen University Medical Centre. *The German Dementia Competence Network (DCN) cohort*: Individuals from the DCN cohort were recruited from 14 university hospital memory clinics across Germany between 2003 and 2005 (PMID: 19339779). The study was approved by the respective ethics committees, and written informed consent was obtained from all the participants prior to inclusion. *The German Study on Aging, Cognition, and Dementia (AgeCoDe)*: The AgeCoDe study is a general practice (GP) registry-based longitudinal study in elderly individuals that recruited patients aged 75 years and above in six German cities from 2003 to 2004 (PMID: 17848793). The study was approved by the respective ethics committees, and written informed consent was obtained from all the participants prior to inclusion.

<u>Greece</u>: the HELIAD study, comprising 49 AD cases and 1,150 controls. HELIAD is a population-based, multidisciplinary, collaborative study designed to estimate, in the Greek population over the age of 64 years, the prevalence and incidence of MCI, AD, other forms of dementia, and other neuropsychiatric conditions of aging and to investigate associations between nutrition and cognitive dysfunction or age-related neuropsychiatric diseases. The participants were selected through random sampling from the records of two Greek municipalities, Larissa and Marousi. All the participants signed informed consent in Greek.

This study was supported by the following grants: IIRG-09-133014 from the Alzheimer's Association; 189 10276/8/9/2011 from the ESPA-EU program Excellence Grant (ARISTEIA), which is co-funded by the European Social Fund and Greek National Resources; and DY2b/oik.51657/14.4.2009 from the Ministry for Health and Social Solidarity(Greece).

<u>Portugal:</u> the Lisbon study from Portugal, totalling 78 AD cases and 74 controls. This cohort was recruited in 2008–2009 to investigate the connections between oxidative stress and lipid dyshomeostasis in AD. The

project includes 190 subjects and was approved by the local ethics committee, and all the participants provided written informed consent. This study includes healthy and dementia-by-AD subjects.
Spain: Those samples are part of DEGESCO. DEGESCO Centers from whom DNA samples were genotyped in the German node (1,778 cases and 470 controls) were the Alzheimer Research Center and Memory Clinic, Fundació ACE, Institut Català de Neurociències Aplicades (Barcelona), the Neurology Service at University Hospital Marqués de Valdecilla (Santander), the Alzheimer's disease and other cognitive disorders, Neurology Department, at Hospital Clínic, IDIBAPS (Barcelona), the Molecular Genetics Laboratory, at the Hospital Universitario Central de Asturias (Oviedo), and Fundació Docència i Recerca Mútua de Terrassa and Movement Disorders Unit, Department of Neurology, University Hospital Mútua de Terrassa (Barcelona).
Switzerland: Two datasets from Switzerland and Austria were combined, totaling 182 AD cases and 388 controls. The Lausanne study: This study includes 137 community-dwelling participants aged 55+ years with cognitive impairment (memory clinic patients with MCI, dementia) or normal cognition (recruited by advertisement, word of mouth). The study's ethical statement was provided locally at the Department of Psychiatry, Geneva University Centre, Switzerland. The VITA study: This is a longitudinal study of 606 individuals (Vienna, Austria) who were 75 years old in 2000, followed up every 30–90 months. This cohort includes dementia and healthy subjects. All the participants gave written informed consent. The study conformed to the latest version of the Declaration of Helsinki and was approved by the ethics committee of the City of Vienna, Austria.
**EADB-Netherlands.** In the Dutch node, samples were collected from six organizations in the Netherlands and after QCs, we obtained 2,438 AD cases and 2,389 controls. All these samples were genotyped at the Erasmus Medical University (Rotterdam, The Netherlands). The Medical Ethics Committee (METC) of the local institutes approved the studies. All the participants and/or their legal guardians gave written informed consent for participation in the clinical and genetic studies. Samples from the following institutes were included. 1) *Erasmus Medical Center*: most individuals were selected from population studies from the epidemiology department and accounted for most of the controls, while a smaller subset of samples originated from the neurology department, where AD was diagnosed according to the National Institute of Neurological and Communicative Disorders and Stroke-Alzheimer's Disease and Related Disorders Association (NINCDS-ADRDA) criteria for AD (PMID: 21514250). 2) *The Amsterdam Dementia Cohort (ADC)* (PMID: 29562540): This cohort comprises patients who visit the memory clinic of the VU University Medical Centre, the Netherlands. The diagnosis of probable AD is based on the clinical criteria formulated by the NINCDS–ADRDA and based on the NIA–AA. Diagnosis of MCI was made according to Petersen and NIA-AA. Controls presented with subjective cognitive decline at the memory clinic, but performed within normal limits on all clinical investigations. 3) *The 100-Plus study*: This study includes Dutch-speaking individuals who (i) can provide official evidence for being aged 100 years or older, (ii) self-report to be cognitively healthy, which is confirmed by a proxy, (iii) consent to the donation of a blood sample, (iv) consent to (at least) two home visits from a researcher, and (v) consent to undergo an interview and neuropsychological test battery (PMID: 30362018). 4) *Parelsnoer Institute*: a collaboration between 8 Dutch University Medical Centers in which clinical data and biomaterials from patients suffering from chronic diseases (so called "Pearls") are collected according to harmonized protocols. The Pearl Neurodegenerative Diseases (PMID: 25551191) includes individuals diagnosed with dementia, mild cognitive impairment, and controls with subjective memory complaints. 5) *The Netherlands Brain Bank*: a non-profit organization that collects human brain tissue of donors with a variety of neurological and psychiatric disorders, but also of non-diseased donors. A clinical diagnosis of AD is based on the clinical criteria of probable AD (PMID: 6610841, PMID: 17616482). The selected AD patients for this study all received a definitive diagnosis which was based on autopsy. 6) *Maastricht University Medical Center*: a subset of individuals that were referred to the memory clinic for cognitive complaints were included if they participated in the BioBank-Alzheimer Centrum Limburg (BB-ACL). Diagnosis of MCI was made according to the criteria of Petersen, and diagnosis of AD-type dementia was made according to the criteria of the DSM-4, and the NINCDS-ADRDA (PMID: 6610841).

**Genotyping**. EADB genomic DNA samples were transferred to 3 genotyping centers and samples that passed the DNA QC were genotyped with the Illumina Infinium Global Screening Array. Raw probe intensities were shared with the CNRGH, which performed the genotype calling on all samples using the same custom cluster file. During the genotyping QC process, three genotyping batches were considered: (1) 49 genotyping chips were identified as possibly problematic and thus were considered as a separate batch, (2) a batch of samples was genotyped and processed after all other samples and (3) the main batch including all other samples. Further details are available in *Bellenguez et al 2022*.

**Additional support for EADB cohorts was provided by:** The work for this manuscript was further supported by the CoSTREAM project (www.costream.eu) and funding from the European Union's Horizon 2020 research and innovation programme under grant agreement No 667375. Italian Ministry of Health (Ricerca Corrente); Ministero dell'Istruzione, dell'Università e della Ricerca–MIUR project “Dipartimenti di Eccellenza 2018–2022” to Department of Neuroscience “Rita Levi Montalcini”, University of Torino (IR), and AIRAlzh Onlus-ANCC-COOP (SB); Partly supported by “Ministero della Salute”, I.R.C.C.S. Research Program, Ricerca Corrente 2018-2020, Linea n. 2 “Meccanismi genetici, predizione e terapie innovative delle malattie complesse” and by the “5 x 1000” voluntary contribution to the Fondazione I.R.C.C.S. Ospedale “Casa Sollievo della Sofferenza”; and RF-2018-12366665, Fondi per la ricerca 2019 (Sandro Sorbi). Karolinska Institutet AD cohort: Dr. Graff and co-authors of the Karolinska Institutet AD cohort report grants from Swedish Research Council (VR) 2015-02926, 2018-02754, 2015-06799, Swedish Alzheimer Foundation, Stockholm County Council ALF and resarch school, Karolinska Institutet StratNeuro, Swedish Demensfonden, and Swedish brain foundation, during the conduct of the study. ADGEN: This work was supported by Academy of Finland (grant numbers 307866); Sigrid Jusélius Foundation; the Strategic Neuroscience Funding of the University of Eastern Finland; EADB project in the JPNDCO-FUND program (grant number 301220). CBAS: Supported by the project no. LQ1605 from the National Program of Sustainability II (MEYS CR), Supported by Ministry of Health of the Czech Republic, grant nr. NV19-04-00270 (All rights reserved), Grant Agency of Charles University Grants No. 693018 and 654217; the Ministry of Health, Czech Republic—conceptual development of research organization, University Hospital Motol, Prague, Czech Republic Grant No. 00064203; the Czech Ministry of Health Project AZV Grant No. 16—27611A; and Institutional Support of Excellence 2. LF UK Grant No. 699012. CNRMAJ-Rouen: This study received fundings from the Centre National de Référence Malades Alzheimer Jeunes (CNRMAJ). The Finnish Geriatric Intervention Study for the Prevention of Cognitive Impairment and Disability (FINGER) data collection was supported by grants from the Academy of Finland, La Carita Foundation, Juho Vainio Foundation, Novo Nordisk Foundation, Finnish Social Insurance Institution, Ministry of Education and Culture Research Grants, Yrjö Jahnsson Foundation, Finnish Cultural Foundation South Osthrobothnia Regional Fund, and EVO/State Research Funding grants of University Hospitals of Kuopio, Oulu and Turku, Seinäjoki Central Hospital and Oulu City Hospital, Alzheimer's Research & Prevention Foundation USA, AXA Research Fund, Knut and Alice Wallenberg Foundation Sweden, Center for Innovative Medicine (CIMED) at Karolinska Institutet Sweden, and Stiftelsen Stockholms sjukhem Sweden. FINGER cohort genotyping was funded by EADB project in the JPND CO-FUND (grant number 301220). Research at the Belgian EADB site is funded in part by the Alzheimer Research Foundation (SAO-FRA), The Research Foundation Flanders (FWO), and the University of Antwerp Research Fund. FK is supported by a BOF DOCPRO fellowship of the University of Antwerp Research Fund. SNAC-K is financially supported by the Swedish Ministry of Health and Social Affairs, the participating County Councils and Municipalities, and the Swedish Research Council. BDR Bristol: We would like to thank the South West Dementia Brain Bank (SWDBB) for providing brain tissue for this study. The SWDBB is part of the Brains for Dementia Research programme, jointly funded by Alzheimer’s Research UK and Alzheimer’s Society and is supported by BRACE (Bristol Research into Alzheimer’s and Care of the Elderly) and the Medical Research Council. BDR Manchester: We would like to thank the Manchester Brain Bankfor providing brain tissue for this study. The Manchester Brain Bank is part of the Brains for Dementia Research programme, jointly funded by Alzheimer’s Research UK and Alzheimer’s Society. BDR KCL: Human post-mortem tissue was provided by the London Neurodegenerative Diseases Brain Bank which receives funding from the UK Medical Research Council and as part of the Brains for Dementia Research programme, jointly funded by Alzheimer's Research UK and the Alzheimer's Society. The CFAS Wales study was funded by the ESRC (RES-060-25-0060) and HEFCW as 'Maintaining function and well-being in later life: a longitudinal cohort study', (Principal Investigators: R.T Woods, L.Clare, G.Windle, V. Burholt, J. Philips, C. Brayne, C. McCracken, K. Bennett, F. Matthews). We are grateful to the NISCHR Clinical Research Centre for their assistance in tracing participants and in interviewing and in collecting blood samples, and to general practices in the study areas for their cooperation. MRC: We thank all individuals who participated in this study. Cardiff University was supported by the Alzheimer's Society (AS; grant RF014/164) and the Medical Research Council (MRC; grants G0801418/1, MR/K013041/1, MR/L023784/1) (R. Sims is an AS Research Fellow). Cardiff University was also supported by the European Joint Programme for Neurodegenerative Disease (JPND; grant MR/L501517/1), Alzheimer's Research UK (ARUK; grant ARUK-PG2014-1), the Welsh Assembly Government (grant SGR544:CADR), Brain's for dementia Research and a donation from the Moondance Charitable Foundation. Cardiff University acknowledges the support of the UK Dementia Research Institute, of which J. Williams is an associate director. Cambridge University acknowledges support from the MRC. Patient recruitment for the MRC Prion Unit/UCL Department of Neurodegenerative Disease collection was supported by the UCLH/UCL Biomedical Centre and NIHR Queen Square Dementia Biomedical Research Unit. The University of Southampton acknowledges support from the AS. King's College London was supported by the NIHR Biomedical Research Centre for Mental Health and the Biomedical Research Unit for Dementia at the South London and Maudsley NHS Foundation Trust and by King's College London and the MRC. ARUK and the Big Lottery Fund provided support to Nottingham University. Alfredo Ramirez: Part of the work was funded by the JPND EADB grant (German Federal Ministry of Education and Research (BMBF) grant: 01ED1619A). German Study on Ageing, Cognition and Dementia in Primary Care Patients (AgeCoDe): This study/publication is part of the German Research Network on Dementia (KND), the German Research Network on Degenerative Dementia (KNDD; German Study on Ageing, Cognition and Dementia in Primary Care Patients; AgeCoDe), and the Health Service Research Initiative (Study on Needs, health service use, costs and health-related quality of life in a large sample of oldestold primary care patients (85+; AgeQualiDe)) and was funded by the German Federal Ministry of Education and Research (grants KND: 01GI0102, 01GI0420, 01GI0422, 01GI0423, 01GI0429, 01GI0431, 01GI0433, 01GI0434; grants KNDD: 01GI0710, 01GI0711, 01GI0712, 01GI0713, 01GI0714, 01GI0715, 01GI0716; grants Health Service Research Initiative: 01GY1322A, 01GY1322B, 01GY1322C, 01GY1322D, 01GY1322E, 01GY1322F, 01GY1322G). In addition, this publication was supported by the German Federal Ministry of Education and Research (Grant numbers: PreADAPT project 01ED2007A and DESCARTES project 01EK2102B to Alfredo Ramirez, 01EK2102A to Anja Schneider, and 01EK2102C to Michael Wagner. VITA study: The support of the Ludwig Boltzmann Society and the AFI Germany have supported the VITA study. The former VITA study group should be acknowledged: W. Danielczyk, G. Gatterer, K Jellinger, S Jugwirth, KH Tragl, S Zehetmayer. Vogel Study: This work was financed by a research grant of the ''Vogelstiftung Dr. Eckernkamp''. HELIAD study: This study was supported by the grants: IIRG-09-133014 from the Alzheimer's Association, 189 10276/8/9/2011 from the ESPA-EU program Excellence Grant (ARISTEIA) and the ΔY2β/οικ.51657/14.4.2009 of the Ministry for Health and Social Solidarity (Greece). Biobank Department of Psychiatry, UMG: Prof. Jens Wiltfang is supported by an Ilídio Pinho professorship and iBiMED (UID/BIM/04501/2013), and FCT project PTDC/DTP_PIC/5587/2014 at the University of Aveiro, Portugal. Lausanne study: This work was supported by grants from the Swiss National Research Foundation (SNF 320030_141179). PAGES study: Harald Hampel is an employee of Eisai Inc. During part of this work he was supported by the AXA Research Fund, the "Fondation partenariale Sorbonne Université" and the "Fondation pour la Recherche sur Alzheimer", Paris, France. Mannheim, Germany Biobank: Department of geriatric Psychiatry, Central Institute for Mental Health, Mannheim, University of Heidelberg, Germany. Genotyping for the Swedish Twin Studies of Aging was supported by NIH/NIA grant R01 AG037985. Genotyping in TwinGene was supported by NIH/NIDDK U01 DK066134. WvdF is recipient of Joint Programming for Neurodegenerative Diseases (JPND) grants PERADES (ANR-

13-JPRF-0001) and EADB (733051061). Gothenburg Birth Cohort (GBC) Studies: We would like to thank UCL Genomics for performing the genotyping analyses. The studies were supported by The Stena Foundation, The Swedish Research Council (2015-02830, 2013-8717), The Swedish Research Council for Health, Working Life and Wellfare (2013-1202, 2005-0762, 2008-1210, 2013-2300, 2013- 2496, 2013-0475), The Brain Foundation, Sahlgrenska University Hospital (ALF), The Alzheimer's Association (IIRG-03-6168), The Alzheimer's Association Zenith Award (ZEN-01-3151), Eivind och Elsa K:son Sylvans Stiftelse, The Swedish Alzheimer Foundation. Clinical AD, Sweden: We would like to thank UCL Genomics for performing the genotyping analyses. Barcelona Brain Biobank: Brain Donors of the Neurological Tissue Bank of the Biobanc-Hospital Clinic-IDIBAPS and their families for their generosity. Hospital Clínic de Barcelona Spanish Ministry of Economy and Competitiveness-Instituto de Salud Carlos III and Fondo Europeo de Desarrollo Regional (FEDER), Unión Europea, "Una manera de hacer Europa" grants (PI16/0235 to Dr. R. Sánchez-Valle and PI17/00670 to Dr. A.Antonelli). AA is funded by Departament de Salut de la Generalitat de Catalunya, PERIS 2016-2020 (SLT002/16/00329). Work at JP-T laboratory was possible thanks to funding from Ciberned and generous gifts from Consuelo Cervera Yuste and Juan Manuel Moreno Cervera. Sydney Memory and Ageing Study (Sydney MAS): We gratefully acknowledge and thank the following for their contributions to Sydney MAS: participants, their supporters and the Sydney MAS Research Team (current and former staff and students). Funding was awarded from the Australian National Health and Medical Research Council (NHMRC) Program Grants (350833, 568969, 109308). AddNeuroMed consortium was led by Simon Lovestone, Bruno Vellas, Patrizia Mecocci, Magda Tsolaki, Iwona Kłoszewska, Hilkka Soininen. This work was supported by InnoMed (Innovative Medicines in Europe), an integrated project funded by the European Union of the Sixth Framework program priority (FP6-2004- LIFESCIHEALTH-5). Oviedo: This work was partly supported by Grant from Fondo de Investigaciones Sanitarias-Fondos FEDER EuropeanUnion to Victoria Alvarez PI15/00878. Pascual Sánchez-Juan is supported by CIBERNED and Carlos III Institute of Health, Spain (PI08/0139, PI12/02288, and PI16/01652), jointly funded by Fondo Europeo de Desarrollo Regional (FEDER), Unión Europea, "Una manera de hacer Europa". Project MinE: The ProjectMinE study was supported by the ALS Foundation Netherlands and the MND association (UK) (Project MinE, www.projectmine.com). The SPIN cohort: We are indebted to patients and their families for their participation in the "Sant Pau Initiative on Neurodegeneration cohort", at the Sant Pau Hospital (Barcelona). This is a multimodal research cohort for biomarker discovery and validation that is partially funded by Generalitat de Catalunya (2017 SGR 547 to JC), as well as from the Institute of Health Carlos III-Subdirección General de Evaluación and the Fondo Europeo de Desarrollo Regional (FEDER- "Una manera de Hacer Europa") (grants PI11/02526, PI14/01126, and PI17/01019 to JF; PI17/01895 to AL), and the Centro de Investigación Biomédica en Red Enfermedades Neurodegenerativas programme (Program 1, Alzheimer Disease to AL). We would also like to thank the Fundació Bancària Obra Social La Caixa (DABNI project) to JF and AL; and Fundación BBVA (to AL), for their support in funding this follow-up study. Adolfo López de Munain is supported by Fundación Salud 2000 (PI2013156), CIBERNED and Diputación Foral de Gipuzkoa (Exp.114/17).

#### 1.2 FinnGen

**Current FinnGen Acknowledgment Statement**

We want to acknowledge the participants and investigators of the FinnGen study. The FinnGen project is funded by two grants from Business Finland (HUS 4685/31/2016 and UH 4386/31/2016) and the following industry partners: AbbVie Inc., AstraZeneca UK Ltd, Biogen MA Inc., Bristol Myers Squibb Inc. (and Celgene Corporation & Celgene International II Sàrl), Genentech Inc., Merck Sharp & Dohme LCC, Pfizer Inc., GlaxoSmithKline Intellectual Property Development Ltd., Sanofi US Services Inc., Maze Therapeutics Inc.,

Johnson&Johnson Innovative Medicine Inc., Novartis AG, Boehringer Ingelheim International GmbH and Bayer AG. Following biobanks are acknowledged for delivering biobank samples to FinnGen: Auria Biobank (www.auria.fi/biopankki), THL Biobank (www.thl.fi/biobank), Helsinki Biobank (www.helsinginbiopankki.fi), Biobank Borealis of Northern Finland (https://www.ppshp.fi/Tutkimus-ja-opetus/Biopankki/Pages/Biobank-Borealis-briefly-in-English.aspx), Finnish Clinical Biobank Tampere (www.tays.fi/en-US/Research_and_development/Finnish_Clinical_Biobank_Tampere), Biobank of Eastern Finland (www.ita-suomenbiopankki.fi/en), Central Finland Biobank (www.ksshp.fi/fi-FI/Potilaalle/Biopankki), Finnish Red Cross Blood Service Biobank (www.veripalvelu.fi/verenluovutus/biopankkitoiminta), Terveystalo Biobank (www.terveystalo.com/fi/Yritystietoa/Terveystalo-Biopankki/Biopankki/) and Arctic Biobank (https://www.oulu.fi/en/university/faculties-and-units/faculty-medicine/northern-finland-birth-cohorts-and-arctic-biobank). All Finnish Biobanks are members of BBMRI.fi infrastructure (https://www.bbmri-eric.eu/national-nodes/finland/). Finnish Biobank Cooperative -FINBB (https://finbb.fi/) is the coordinator of BBMRI-ERIC operations in Finland. The Finnish biobank data can be accessed through the Fingenious® services (https://site.fingenious.fi/en/) managed by FINBB.

**FinnGen R11 ethics statement and materials & methods**

Study subjects in FinnGen provided informed consent for biobank research, based on the Finnish Biobank Act. Alternatively, separate research cohorts, collected prior the Finnish Biobank Act came into effect (in September 2013) and start of FinnGen (August 2017), were collected based on study-specific consents and later transferred to the Finnish biobanks after approval by Fimea (Finnish Medicines Agency), the National Supervisory Authority for Welfare and Health. Recruitment protocols followed the biobank protocols approved by Fimea. The Coordinating Ethics Committee of the Hospital District of Helsinki and Uusimaa (HUS) statement number for the FinnGen study is Nr HUS/990/2017.

The FinnGen study is approved by Finnish Institute for Health and Welfare (permit numbers: THL/2031/6.02.00/2017, THL/1101/5.05.00/2017, THL/341/6.02.00/2018, THL/2222/6.02.00/2018, THL/283/6.02.00/2019, THL/1721/5.05.00/2019 and THL/1524/5.05.00/2020), Digital and population data service agency (permit numbers: VRK43431/2017-3, VRK/6909/2018-3, VRK/4415/2019-3), the Social Insurance Institution (permit numbers: KELA 58/522/2017, KELA 131/522/2018, KELA 70/522/2019, KELA 98/522/2019, KELA 134/522/2019, KELA 138/522/2019, KELA 2/522/2020, KELA 16/522/2020), Findata permit numbers THL/2364/14.02/2020, THL/4055/14.06.00/2020, THL/3433/14.06.00/2020, THL/4432/14.06/2020, THL/5189/14.06/2020, THL/5894/14.06.00/2020, THL/6619/14.06.00/2020, THL/209/14.06.00/2021, THL/688/14.06.00/2021, THL/1284/14.06.00/2021, THL/1965/14.06.00/2021, THL/5546/14.02.00/2020, THL/2658/14.06.00/2021, THL/4235/14.06.00/2021, Statistics Finland (permit numbers: TK-53-1041-17 and TK/143/07.03.00/2020 (earlier TK-53-90-20) TK/1735/07.03.00/2021, TK/3112/07.03.00/2021) and Finnish Registry for Kidney Diseases permission/extract from the meeting minutes on 4th July 2019.

The Biobank Access Decisions for FinnGen samples and data utilized in FinnGen Data Freeze 11 include: THL Biobank BB2017_55, BB2017_111, BB2018_19, BB_2018_34, BB_2018_67, BB2018_71, BB2019_7, BB2019_8, BB2019_26, BB2020_1, BB2021_65, Finnish Red Cross Blood Service Biobank 7.12.2017, Helsinki Biobank HUS/359/2017, HUS/248/2020,

HUS/430/2021 §28, §29, HUS/150/2022 §12, §13, §14, §15, §16, §17, §18, §23, §58 and §59, Auria Biobank AB17-5154 and amendment #1 (August 17 2020) and amendments BB_2021-0140, BB_2021-0156 (August 26 2021, Feb 2 2022), BB_2021-0169, BB_2021-0179, BB_2021-0161, AB20-5926 and amendment #1 (April 23 2020) and it´s modification (Sep 22 2021), BB_2022-0262, BB_2022-0256, Biobank Borealis of Northern Finland_2017_1013, 2021_5010, 2021_5018, 2021_5015, 2021_5015 Amendment, 2021_5023, 2021_5023 Amendment, 2021_5017, 2022_6001, 2022_6006 Amendment, BB22-0067, 2022_0262, Biobank of Eastern Finland 1186/2018 and amendment 22§/2020, 53§/2021, 13§/2022, 14§/2022, 15§/2022, 27§/2022, 28§/2022, 29§/2022, 33§/2022, 35§/2022, 36§/2022, 37§/2022, 39§/2022, 7§/2023, Finnish Clinical Biobank Tampere MH0004 and amendments (21.02.2020 & 06.10.2020), 8§/2021, 9§/2021, §9/2022, §10/2022, §12/2022, 13§/2022, §20/2022, §21/2022, §22/2022, §23/2022, 28§/2022, 29§/2022, 30§/2022, 31§/2022, 32§/2022, 38§/2022, 40§/2022, 42§/2022, 1§/2023, Central Finland Biobank 1-2017, BB_2021-0161, BB_2021-0169, BB_2021-0179, BB_2021-0170, BB_2022-0256, and Terveystalo Biobank STB 2018001 and amendment 25th Aug 2020, Finnish Hematological Registry and Clinical Biobank decision 18th June 2021, Arctic biobank P0844: ARC_2021_1001.

#### 1.3 UK Biobank (UKB)

The UK Biobank is comprised of environmental, lifestyle, and genetic data for 500,000 participants. Cases and controls for this analysis were included based on similar criteria to what was used for the FinnGen cohort. AD cases included participants with an ICD10 code of F00 or G30 (UKB codes p130836 and p131036). Controls included participants that did not have an algorithmically defined AD diagnosis as assigned by UKB (UKB code p42020) or any ICD10 code of A81, F01, F02, G31, F03, or I673 (UKB codes p130136, p130838, p130840, p131038, p130842 or p41270). Controls were also filtered for participants currently over 60 years of age to approximate the age distribution of the cases. Cases and controls were filtered for Caucasian ancestry and related individuals removed.

UKB genetic data was genotyped and underwent quality control as described elsewhere (PMID: 30305743). For this manuscript, data imputed using the TOPMed reference panel was used for determining *APOE* genotype and for all subsequent analyses. Data was accessed and analysed on the UKB Research Analysis Platform (RAP) under UK Biobank application ID 33601.

## 1.4 GR@CE

The GR@ACE study (PMID: 31473137) recruited Alzheimer's disease (AD) patients from Fundació ACE, Institut Català de Neurociències Aplicades (Catalonia, Spain), and control individuals from three centers: Fundació ACE (Barcelona, Spain), Valme University Hospital (Seville, Spain), and the Spanish National DNA Bank–Carlos III (University of Salamanca, Spain) (http://www.bancoadn.org). At all sites, AD diagnosis was established by a multidisciplinary working group—including neurologists, neuropsychologists, and social workers—according to the DSM-IV criteria for dementia and the National Institute on Aging and Alzheimer's Association's (NIA–AA) 2011 guidelines for diagnosing AD. In our study, we considered as AD cases any individuals with dementia diagnosed with probable or possible AD at any point in their clinical course. Genotyping was conducted using the Axiom 815K Spanish biobank array (Thermo Fisher) at the Spanish National Centre for Genotyping (CeGEN, Santiago de Compostela, Spain). The genotyping array not only is an adaptation of the Axiom biobank genotyping array but also contains rare population-specific variations observed in the Spanish population.

The Genome Research @ Fundació ACE project (GR@ACE) is supported by Grifols SA, Fundación bancaria 'La Caixa', Fundació ACE, and CIBERNED (Centro de Investigación Biomédica en Red Enfermedades Neurodegenerativas (Program 1, Alzheimer Disease to MB and AR)). A.R. and M.B. receive support from the European Union/EFPIA Innovative Medicines Initiative Joint undertaking ADAPTED and MOPEAD projects (grant numbers 115975 and 115985, respectively). M.B. and A.R. are also supported by national grants PI13/02434, PI16/01861, PI17/01474 and PI19/01240. Acción Estratégica en Salud is integrated into the Spanish National R + D + I Plan and funded by ISCIII (Instituto de Salud Carlos III)–Subdirección General de Evaluación and the Fondo Europeo de Desarrollo Regional (FEDER–'Una manera de hacer Europa'). Some control samples and data from patients included in this study were provided in part by the National DNA Bank Carlos III (www.bancoadn.org, University of Salamanca, Spain) and Hospital Universitario Virgen de Valme (Sevilla, Spain); they were processed following standard operating procedures with the appropriate approval of the Ethical and Scientific Committee.

#### 1.5 European Alzheimer's Disease Initiative (EADI) Consortium

EADI is composed of several case-control studies and one population-based cohort, the 3C study (PMID: 14598854, PMID: 19734903). Case-control studies are comprised of AD cases and cognitively normal controls across France. The population-based cohort, the 3C study, is a prospective study of the relationship between vascular factors and dementia carried out in the three French cities Bordeaux, Montpellier and Dijon. The AD status was defined based on 12 years follow-up for Dijon participants, 14-15 years follow-up for Montpellier participants and 17-18 years follow-up for Bordeaux participants. All other non demented subjects of 3C were included as controls. All AD cases, both in the case-control studies and the 3C study, were ascertained by neurologists and the clinical diagnosis of probable AD was established according to the DSM-III-R and NINCDS-ADRDA criteria. Samples that passed DNA quality control were genotyped with Illumina Human 610-Quad BeadChips.

This work has been developed and supported by the LABEX (laboratory of excellence program investment for the future) DISTALZ grant (Development of Innovative Strategies for a Transdisciplinary approach to ALZheimer's disease) including funding from MEL (Metropole européenne de Lille), ERDF (European Regional Development Fund) and Conseil Régional Nord Pas de Calais. This work was supported by INSERM, the National Foundation for Alzheimer's disease and related disorders, the Institut Pasteur de Lille and the Centre National de Recherche en Génomique Humaine, CEA, the JPND PERADES, the Laboratory of Excellence GENMED (Medical Genomics) grant no. ANR-10-LABX-0013 managed by the National Research Agency (ANR) part of the Investment for the Future program, and the FP7 AgedBrainSysBio. The Three-City Study was performed as part of collaboration between the Institut National de la Santé et de la Recherche Médicale (Inserm), the Victor Segalen Bordeaux II University and Sanofi-Synthélabo. The Fondation pour la Recherche Médicale funded the preparation and initiation of the study. The 3C Study was also funded by the Caisse Nationale Maladie des Travailleurs Salariés, Direction Générale de la Santé, MGEN, Institut de la Longévité, Agence Française de Sécurité Sanitaire des Produits de Santé, the Aquitaine and Bourgogne Regional Councils, Agence Nationale de la Recherche, ANR supported the COGINUT and COVADIS projects. Fondation de France and the joint French Ministry of Research/INSERM "Cohortes et collections de données biologiques" programme. Lille Génopôle received an unconditional grant from Eisai. The Three-city biological bank was developed and maintained by the laboratory for genomic analysis LAG-BRC - Institut Pasteur de Lille.

#### 1.6 Genetic and Environmental Risk in AD (GERAD) Consortium

Genetic and Environmental Risk in AD (GERAD) Consortium/Defining Genetic, Polygenic, and Environmental Risk for Alzheimer's Disease (PERADES) Consortium
The GERAD/PERADES sample comprises 3,177 Alzheimer's disease cases and 7,277 controls with available age and gender data (PMID: 19734902). Cases and elderly screened controls were recruited by the Medical Research Council (MRC) Genetic Resource for Alzheimer's disease (Cardiff University; Institute of Psychiatry, London; Cambridge University; Trinity College Dublin), the Alzheimer's Research Trust (ART) Collaboration (University of Nottingham; University of Manchester; University of Southampton; University of Bristol; Queen's University Belfast; the Oxford Project to Investigate Memory and Ageing (OPTIMA), Oxford University); Washington University, St Louis, United States; MRC PRION Unit, University College

London; London and the South East Region Alzheimer's disease project (LASER-AD), University College London; Competence Network of Dementia (CND) and Department of Psychiatry, University of Bonn, Germany; the National Institute of Mental Health (NIMH) Alzheimer's disease Genetics Initiative. 6129 population controls were drawn from large existing cohorts with available GWAS data, including the 1958 British Birth Cohort (1958BC) (http://www.b58cgene.sgul.ac.uk), the KORA F4 Study and the Heinz Nixdorf Recall Study. All Alzheimer's disease cases met criteria for either probable (NINCDS-ADRDA, DSM-IV) or definite (CERAD) Alzheimer's disease. All elderly controls were screened for dementia using the MMSE or ADAS-cog, were determined to be free from dementia at neuropathological examination or had a Braak score of 2.5 or lower. Genotypes from all cases and 4617 controls were previously included in the AD GWAS by Harold and colleagues (PMID: 19734902). Genotypes for the remaining 2660 population controls were obtained from WTCCC2.

We thank all individuals who participated in this study. Cardiff University was supported by the Wellcome Trust, Alzheimer's Society (AS; grant RF014/164), the Medical Research Council (MRC; grants G0801418/1, MR/K013041/1, MR/L023784/1), the European Joint Programme for Neurodegenerative Disease (JPND, grant MR/L501517/1), Alzheimer's Research UK (ARUK, grant ARUK-PG2014-1), Welsh Assembly Government (grant SGR544:CADR), a donation from the Moondance Charitable Foundation, UK Dementia's Platform (DPUK, reference MR/L023784/1), and the UK Dementia Research Institute at Cardiff. Cambridge University acknowledges support from the MRC. ARUK supported sample collections at the Kings College London, the South West Dementia Bank, Universities of Cambridge, Nottingham, Manchester and Belfast. King's College London was supported by the NIHR Biomedical Research Centre for Mental Health and Biomedical Research Unit for Dementia at the South London and Maudsley NHS Foundation Trust and Kings College London and the MRC. Alzheimer's Research UK (ARUK) and the Big Lottery Fund provided support to Nottingham University. Ulster Garden Villages, AS, ARUK, American Federation for Aging Research, NI R&D Office and the Royal College of Physicians/Dunhill Medical Trust provided support for Queen's University, Belfast. The University of Southampton acknowledges support from the AS. The MRC and Mercer's Institute for Research on Ageing supported the Trinity College group. DCR is a Wellcome Trust Principal Research fellow. The South West Dementia Brain Bank acknowledges support from Bristol Research into Alzheimer's and Care of the Elderly. The Charles Wolfson Charitable Trust supported the OPTIMA group. Washington University was funded by NIH grants, Barnes Jewish Foundation and the Charles and Joanne Knight Alzheimer's Research Initiative. Patient recruitment for the MRC Prion Unit/UCL Department of Neurodegenerative Disease collection was supported by the UCLH/UCL Biomedical Research Centre and their work was supported by the NIHR Queen Square Dementia BRU, the Alzheimer's Research UK and the Alzheimer's Society. LASER-AD was funded by Lundbeck SA. The AgeCoDe study group was supported by the German Federal Ministry for Education and Research grants 01 GI 0710, 01 GI 0712, 01 GI 0713, 01 GI 0714, 01 GI 0715, 01 GI 0716, 01 GI 0717. Genotyping of the Bonn case-control sample was funded by the German centre for Neurodegenerative Diseases (DZNE), Germany. The GERAD Consortium also used samples ascertained by the NIMH AD Genetics Initiative. HH was supported by a grant of the Katharina-Hardt-Foundation, Bad Homburg vor der Höhe, Germany. The KORA F4 studies were financed by Helmholtz Zentrum München; German Research Center for Environmental Health; BMBF; German National Genome Research Network and the Munich Center of Health Sciences. The Heinz Nixdorf Recall cohort was funded by the Heinz Nixdorf Foundation (Dr. Jur. G.Schmidt, Chairman) and BMBF. We acknowledge use of genotype data from the 1958 Birth Cohort collection and National Blood Service, funded by the MRC and the Wellcome Trust which was genotyped by the Wellcome Trust Case Control Consortium and the Type-1 Diabetes Genetics Consortium, sponsored by the National Institute of Diabetes and Digestive and Kidney Diseases, National Institute of Allergy and Infectious Diseases, National Human Genome Research Institute, National Institute of Child Health and Human Development and Juvenile Diabetes Research Foundation International. The project is also supported through the following funding organisations under the aegis of JPND - www.jpnd.eu (United Kingdom, Medical Research Council (MR/L501529/1; MR/R024804/1) and Economic and Social Research Council

(ES/L008238/1)) and through the Motor Neurone Disease Association. This study represents independent research part funded by the National Institute for Health Research (NIHR) Biomedical Research Centre at South London and Maudsley NHS Foundation Trust and King's College London. Prof Jens Wiltfang is supported by an Ilídio Pinho professorship and iBiMED (UID/BIM/04501/2013), at the University of Aveiro, Portugal.

#### 1.7 Bonn studies

The Bonn group would like to thank Dr. Heike Koelsch for her scientific support.
The Bonn samples are part of the German Dementia Competence Network (DCN) and the German Research Network on Degenerative Dementia (KNDD, AgeCoDe study group), which are funded by the German Federal Ministry of Education and Research (BMBF): grants DCN: 01G10102, 01GI0420, 01GI0422, 01GI0423, 01GI0429, 01GI0431, 01GI0433, 04GI0434, 01GI0711; grants KNDD: 01GI1007A, 01GI0710, 01GI0711, 01GI0712, 01GI0713, 01GI0714, 01GI0715, 01GI0716, 01GI0717, 01ET1006B. Controls for the Bonn samples were also obtained from the Heinz Nixdorf Recall cohort that was funded by the Heinz Nixdorf Foundation (Dr. Jur. G.Schmidt, Chairman) and BMBF. Genotyping of the Bonn case-control sample was funded by the German Centre for Neurodegenerative Diseases (DZNE), Germany. Markus M Nöthen and Alfredo Ramirez are members of the German Research Foundation (DFG) cluster of excellence ImmunoSensation.

Bonn OMNI cohort: the Bonn OMNI cohort consists of AD patients and controls derived from a larger German GWAS cohort which was recruited from the following three sources: (i) the German Dementia Competence Network; (ii) the German study on Aging, Cognition, and Dementia in primary care patients (AgeCoDe); and (iii) the interdisciplinary Memory Clinic at the University Hospital of Bonn. The control sample comprised of individuals from the population-based study Heinz Nixdorf Recall (HNR) study cohort. This sample was previously used for replication in Lambert et al.[33]. *The German study on aging, cognition and dementia (AgeCoDe)*: see description above. *The German competence network cohort (DCN)*: The DCN cohort includes 1,095 patients with mild cognitive impairment (MCI) and 648 cases with mild Alzheimer's disease (AD) clinical dementia syndrome that were recruited from 14 university hospital memory clinics across Germany between 2003 and 2005[9]. Exclusion criteria were substance abuse or dependence, insufficient German language skills, multi-morbidity, comorbid condition with excess mortality, circumstances that would have made regular attendance at follow-up visits questionable and lack of an informant. The diagnosis of mild dementia according to ICD-10 criteria required a decline of cognitive ability (at least 1 SD) from a previous level in at least 2 domains as evidenced by age-corrected standardized tests, impairment in activities of daily living (i.e. B-ADL > 6), changes in personality, drive, social behavior or control of emotion but no clouding of consciousness. These changes must have persisted for at least 3 months. The etiological diagnosis of AD was assigned according to NINCDS-ADRDA criteria[8]. *Memory clinic Bonn*: The interdisciplinary Memory Clinic of the Department of Psychiatry and Department of Neurology at the University Hospital in Bonn provided further patients. Diagnoses were assigned according the NINCDS/ADRDA criteria[8] and on the basis of clinical history, physical examination, neuropsychological testing (using the CERAD neuropsychological battery, including the MMSE), laboratory assessments, and brain imaging. *Control sample:* In the Heinz Nixdorf Recall (Risk Factors, Evaluation of Coronary Calcification, and Lifestyle) study, participants were randomly sampled in three cities in Germany. The study design has previously been described[34,35]. Briefly, 4814 participants aged 45 to 75 years were enrolled between 2000 and 2003 (t0, baseline). Cognitive performance of participants was evaluated at follow up scheduled 5 years after baseline (t1, n = 4157, 2005–2008) and then again at follow up 5 years after t1 (t2, n = 3087, 2010–2015). Controls sample was selected if participant did not present cognitive impairment as reported at the last available evaluation. Cognitive evaluation has been described extensively previously[36,37]. Herein, cognitive impairment at t1 was defined as a performance of one standard deviation (SD) below the age- and education-adjusted mean except for the clock-drawing test, where a performance ≥3 was rated as impaired (for a detailed description, see the study by Winkler et al.[36]). The study was approved by the University of Duisburg-Essen Institutional Review Board and followed established guidelines of good epidemiological practice.

#### 1.8 The Alzheimer's Disease Sequencing Project (ADSP)

The Alzheimer's Disease Sequencing Project (ADSP) is comprised of two Alzheimer's Disease (AD) genetics consortia and three National Human Genome Research Institute (NHGRI) funded Large Scale Sequencing and Analysis Centers (LSAC). The two AD genetics consortia are the Alzheimer's Disease Genetics Consortium (ADGC) funded by NIA (U01 AG032984), and the Cohorts for Heart and Aging Research in Genomic Epidemiology (CHARGE) funded by NIA (R01 AG033193), the National Heart, Lung, and Blood Institute (NHLBI), other National Institute of Health (NIH) institutes and other foreign governmental and non-governmental organizations. The Discovery Phase analysis of sequence data is supported through UF1AG047133 (to Drs. Schellenberg, Farrer, Pericak-Vance, Mayeux, and Haines); U01AG049505 to Dr. Seshadri; U01AG049506 to Dr. Boerwinkle; U01AG049507 to Dr. Wijsman; and U01AG049508 to Dr. Goate and the Discovery Extension Phase analysis is supported through U01AG052411 to Dr. Goate, U01AG052410 to Dr. Pericak-Vance and U01 AG052409 to Drs. Seshadri and Fornage.

Sequencing for the Follow Up Study (FUS) is supported through U01AG057659 (to Drs. PericakVance, Mayeux, and Vardarajan) and U01AG062943 (to Drs. Pericak-Vance and Mayeux). Data generation and harmonization in the Follow-up Phase is supported by U54AG052427 (to Drs. Schellenberg and Wang). The FUS Phase analysis of sequence data is supported through U01AG058589 (to Drs. Destefano, Boerwinkle, De Jager, Fornage, Seshadri, and Wijsman), U01AG058654 (to Drs. Haines, Bush, Farrer, Martin, and Pericak-Vance), U01AG058635 (to Dr. Goate), RF1AG058066 (to Drs. Haines, Pericak-Vance, and Scott), RF1AG057519 (to Drs. Farrer and Jun), R01AG048927 (to Dr. Farrer), and RF1AG054074 (to Drs. Pericak-Vance and Beecham).

The ADGC cohorts include: Adult Changes in Thought (ACT) (U01 AG006781, U19 AG066567), the Alzheimer's Disease Research Centers (ADRC) (P30 AG062429, P30 AG066468, P30 AG062421, P30 AG066509, P30 AG066514, P30 AG066530, P30 AG066507, P30 AG066444, P30 AG066518, P30 AG066512, P30 AG066462, P30 AG072979, P30 AG072972, P30 AG072976, P30 AG072975, P30 AG072978, P30 AG072977, P30 AG066519, P30 AG062677, P30 AG079280, P30 AG062422, P30 AG066511, P30 AG072946, P30 AG062715, P30 AG072973, P30 AG066506, P30 AG066508, P30 AG066515, P30 AG072947, P30 AG072931, P30 AG066546, P20 AG068024, P20 AG068053, P20 AG068077, P20 AG068082, P30 AG072958, P30 AG072959), the Chicago Health and Aging Project (CHAP) (R01 AG11101, RC4 AG039085, K23 AG030944), Indiana Memory and Aging Study (IMAS) (R01 AG019771), Indianapolis Ibadan (R01 AG009956, P30 AG010133), the Memory and Aging Project (MAP) ( R01 AG17917), Mayo Clinic (MAYO) (R01 AG032990, U01 AG046139, R01 NS080820, RF1 AG051504, P50 AG016574), Mayo Parkinson's Disease controls (NS039764, NS071674, 5RC2HG005605), University of Miami (R01 AG027944, R01 AG028786, R01 AG019085, IIRG09133827, A2011048), the Multi-Institutional Research in Alzheimer's Genetic Epidemiology Study (MIRAGE) (R01 AG09029, R01 AG025259), the National Centralized Repository for Alzheimer's Disease and Related Dementias (NCRAD) (U24 AG021886), the National Institute on Aging Late Onset Alzheimer's Disease Family Study (NIA- LOAD) (U24 AG056270), the Religious Orders Study (ROS) (P30 AG10161, R01 AG15819), the Texas Alzheimer's Research and Care Consortium (TARCC) (funded by the Darrell K Royal Texas Alzheimer's Initiative), Vanderbilt University/Case Western Reserve University (VAN/CWRU) (R01 AG019757, R01 AG021547, R01 AG027944, R01 AG028786, P01 NS026630, and Alzheimer's Association), the Washington Heights-Inwood Columbia Aging Project (WHICAP) (RF1 AG054023), the University of Washington Families (VA Research Merit Grant, NIA: P50AG005136, R01AG041797, NINDS: R01NS069719), the Columbia University Hispanic Estudio Familiar de Influencia Genetica de Alzheimer (EFIGA) (RF1 AG015473), the University of Toronto (UT) (funded by Wellcome Trust, Medical Research Council, Canadian Institutes of Health Research), and Genetic Differences (GD) (R01 AG007584). The CHARGE cohorts are supported in part by National Heart, Lung, and Blood Institute (NHLBI) infrastructure grant HL105756 (Psaty), RC2HL102419 (Boerwinkle) and the neurology working group is supported by the National Institute on Aging (NIA) R01 grant AG033193.

The CHARGE cohorts participating in the ADSP include the following: Austrian Stroke Prevention Study (ASPS), ASPS-Family study, and the Prospective Dementia Registry-Austria (ASPS/PRODEM-Aus), the Atherosclerosis Risk in Communities (ARIC) Study, the Cardiovascular Health Study (CHS), the Erasmus Rucphen Family Study (ERF), the Framingham Heart Study (FHS), and the Rotterdam Study (RS). ASPS is funded by the Austrian Science Fond (FWF) grant number P20545-P05 and P13180 and the Medical University of Graz. The ASPS-Fam is funded by the Austrian Science Fund (FWF) project I904), the EU Joint Programme – Neurodegenerative Disease Research (JPND) in frame of the BRIDGET project (Austria, Ministry of Science) and the Medical University of Graz and the Steiermärkische Krankenanstalten Gesellschaft. PRODEM-Austria is supported by the Austrian Research Promotion agency (FFG) (Project No. 827462) and by the Austrian National Bank (Anniversary Fund, project 15435. ARIC research is carried out as a collaborative study supported by NHLBI contracts (HHSN268201100005C, HHSN268201100006C, HHSN268201100007C, HHSN268201100008C, HHSN268201100009C, HHSN268201100010C, HHSN268201100011C, and HHSN268201100012C). Neurocognitive data in ARIC is collected by U01 2U01HL096812, 2U01HL096814, 2U01HL096899, 2U01HL096902, 2U01HL096917 from the NIH (NHLBI, NINDS, NIA and NIDCD), and with previous brain MRI examinations funded by R01-HL70825 from the NHLBI. CHS research was supported by contracts HHSN268201200036C, HHSN268200800007C, N01HC55222, N01HC85079, N01HC85080, N01HC85081, N01HC85082, N01HC85083, N01HC85086, and grants U01HL080295 and U01HL130114 from the NHLBI with additional contribution from the National Institute of Neurological Disorders and Stroke (NINDS). Additional support was provided by R01AG023629, R01AG15928, and R01AG20098 from the NIA. FHS research is supported by NHLBI contracts N01-HC-25195 and HHSN268201500001I. This study was also supported by additional grants from the NIA (R01s AG054076, AG049607 and AG033040 and NINDS (R01 NS017950). The ERF study as a part of EUROSPAN (European Special Populations Research Network) was supported by European Commission FP6 STRP grant number 018947 (LSHG-CT-2006-01947) and also received funding from the European Community's Seventh Framework Programme (FP7/2007-2013)/grant agreement HEALTH-F4- 2007-201413 by the European Commission under the programme "Quality of Life and Management of the Living Resources" of 5th Framework Programme (no. QLG2-CT-2002- 01254). High-throughput analysis of the ERF data was supported by a joint grant from the Netherlands Organization for Scientific Research and the Russian Foundation for Basic Research (NWO-RFBR 047.017.043). The Rotterdam Study is funded by Erasmus Medical Center and Erasmus University, Rotterdam, the Netherlands Organization for Health Research and Development (ZonMw), the Research Institute for Diseases in the Elderly (RIDE), the Ministry of Education, Culture and Science, the Ministry for Health, Welfare and Sports, the European Commission (DG XII), and the municipality of Rotterdam. Genetic data sets are also supported by the Netherlands Organization of Scientific Research NWO Investments (175.010.2005.011, 911-03-012), the Genetic Laboratory of the Department of Internal Medicine, Erasmus MC, the Research

Institute for Diseases in the Elderly (014-93-015; RIDE2), and the Netherlands Genomics Initiative (NGI)/Netherlands Organization for Scientific Research (NWO) Netherlands Consortium for Healthy Aging (NCHA), project 050-060-810. All studies are grateful to their participants, faculty and staff. The content of these manuscripts is solely the responsibility of the authors and does not necessarily represent the official views of the National Institutes of Health or the U.S. Department of Health and Human Services.

The FUS cohorts include: the Alzheimer's Disease Research Centers (ADRC) (P30 AG062429, P30 AG066468, P30 AG062421, P30 AG066509, P30 AG066514, P30 AG066530, P30 AG066507, P30 AG066444, P30 AG066518, P30 AG066512, P30 AG066462, P30 AG072979, P30 AG072972, P30 AG072976, P30 AG072975, P30 AG072978, P30 AG072977, P30 AG066519, P30 AG062677, P30 AG079280, P30 AG062422, P30 AG066511, P30 AG072946, P30 AG062715, P30 AG072973, P30 AG066506, P30 AG066508, P30 AG066515, P30 AG072947, P30 AG072931, P30 AG066546, P20 AG068024, P20 AG068053, P20 AG068077, P20 AG068082, P30 AG072958, P30 AG072959), Alzheimer's Disease Neuroimaging Initiative (ADNI) (U19AG024904), Amish Protective Variant Study (RF1AG058066), Cache County Study (R01AG11380, R01AG031272, R01AG21136, RF1AG054052), Case Western Reserve University Brain Bank (CWRUBB) (P50AG008012), Case Western Reserve University Rapid Decline (CWRURD) (RF1AG058267, NU38CK000480), CubanAmerican Alzheimer's Disease Initiative (CuAADI) (3U01AG052410), Estudio Familiar de Influencia Genetica en Alzheimer (EFIGA) (5R37AG015473, RF1AG015473, R56AG051876), Genetic and Environmental Risk Factors for Alzheimer Disease Among African Americans Study (GenerAAtions) (2R01AG09029, R01AG025259, 2R01AG048927), Gwangju Alzheimer and Related Dementias Study (GARD) (U01AG062602), Hillblom Aging Network (2014-A-004-NET, R01AG032289, R01AG048234), Hussman Institute for Human Genomics Brain Bank (HIHGBB) (R01AG027944, Alzheimer's Association "Identification of Rare Variants in Alzheimer Disease"), Ibadan Study of Aging (IBADAN) (5R01AG009956), Longevity Genes Project (LGP) and LonGenity (R01AG042188, R01AG044829, R01AG046949, R01AG057909, R01AG061155, P30AG038072), Mexican Health and Aging Study (MHAS) (R01AG018016), Multi-Institutional Research in Alzheimer's Genetic Epidemiology (MIRAGE) (2R01AG09029, R01AG025259, 2R01AG048927), Northern Manhattan Study (NOMAS) (R01NS29993), Peru Alzheimer's Disease Initiative (PeADI) (RF1AG054074), Puerto Rican 1066 (PR1066) (Wellcome Trust (GR066133/GR080002), European Research Council (340755)), Puerto Rican Alzheimer Disease Initiative (PRADI) (RF1AG054074), Reasons for Geographic and Racial Differences in Stroke (REGARDS) (U01NS041588), Research in African American Alzheimer Disease Initiative (REAAADI) (U01AG052410), the Religious Orders Study (ROS) (P30 AG10161, P30 AG72975, R01 AG15819, R01 AG42210), the RUSH Memory and Aging Project (MAP) (R01 AG017917, R01 AG42210Stanford Extreme Phenotypes in AD (R01AG060747), University of Miami Brain Endowment Bank (MBB), University of Miami/Case Western/North Carolina A&T African American (UM/CASE/NCAT) (U01AG052410, R01AG028786), and Wisconsin Registry for Alzheimer's Prevention (WRAP) (R01AG027161 and R01AG054047).

The four LSACs are: the Human Genome Sequencing Center at the Baylor College of Medicine (U54 HG003273), the Broad Institute Genome Center (U54HG003067), The American Genome Center at the Uniformed Services University of the Health Sciences (U01AG057659), and the Washington University Genome Institute (U54HG003079). Genotyping and sequencing for the ADSP FUS is also conducted at John P. Hussman Institute for Human Genomics (HIHG) Center for Genome Technology (CGT).

Biological samples and associated phenotypic data used in primary data analyses were stored at Study Investigators institutions, and at the National Centralized Repository for Alzheimer's Disease and Related Dementias (NCRAD, U24AG021886) at Indiana University funded by NIA. Associated Phenotypic Data used in primary and secondary data analyses were provided by Study Investigators, the NIA funded Alzheimer's Disease Centers (ADCs), and the National Alzheimer's Coordinating Center (NACC, U24AG072122) and the National Institute on Aging Genetics of Alzheimer's Disease Data Storage Site (NIAGADS, U24AG041689) at the University of Pennsylvania, funded by NIA. Harmonized phenotypes were provided by the ADSP Phenotype Harmonization Consortium (ADSP-PHC), funded by NIA (U24 AG074855, U01 AG068057 and R01 AG059716) and Ultrascale Machine Learning to Empower Discovery in Alzheimer's Disease Biobanks (AI4AD, U01 AG068057). This research was supported in part by the Intramural Research Program of the National Institutes of health, National Library of Medicine. Contributors to the Genetic Analysis Data included Study Investigators on projects that were individually funded by NIA, and other NIH institutes, and by private U.S. organizations, or foreign governmental or nongovernmental organizations.

The ADSP Phenotype Harmonization Consortium (ADSP-PHC) is funded by NIA (U24 AG074855, U01 AG068057 and R01 AG059716). The harmonized cohorts within the ADSP-PHC include: the Anti-Amyloid Treatment in Asymptomatic Alzheimer's study (A4 Study), a secondary prevention trial in preclinical Alzheimer's disease, aiming to slow cognitive decline associated with brain amyloid accumulation in clinically normal older individuals. The A4 Study is funded by a public-private-philanthropic partnership, including funding from the National Institutes of Health-National Institute on Aging, Eli Lilly and Company, Alzheimer's Association, Accelerating Medicines Partnership, GHR Foundation, an anonymous foundation and additional private donors, with in-kind support from Avid and Cogstate. The companion observational Longitudinal Evaluation of Amyloid Risk and Neurodegeneration (LEARN) Study is funded by the Alzheimer's Association and GHR Foundation. The A4 and LEARN Studies are led by Dr. Reisa Sperling at Brigham and Women's Hospital, Harvard Medical School and Dr. Paul Aisen at the Alzheimer's Therapeutic Research Institute (ATRI), University of Southern California. The A4 and LEARN Studies are coordinated by ATRI at the University of Southern California, and the data are made available through the Laboratory for Neuro Imaging at the University of Southern California. The participants screening for the A4 Study provided permission to share their de-identified data in order to advance the quest to find a successful treatment for Alzheimer's disease. We would like to acknowledge the dedication of all the participants, the site personnel, and all of the partnership team members who continue to make the A4 and LEARN Studies possible. The complete A4 Study Team list is available on: a4study.org/a4-study-team.; the Adult Changes in Thought study (ACT), U01 AG006781, U19 AG066567; Alzheimer's Disease Neuroimaging Initiative (ADNI): Data collection and sharing for this project was funded by the Alzheimer's Disease Neuroimaging Initiative (ADNI) (National Institutes of Health Grant U01 AG024904) and DOD ADNI (Department of Defense award number W81XWH-12-2-0012). ADNI is funded by the National Institute on Aging, the National Institute of Biomedical Imaging and Bioengineering, and through generous contributions from the following: AbbVie, Alzheimer's Association; Alzheimer's Drug Discovery Foundation; Araclon Biotech; BioClinica, Inc.; Biogen; Bristol-Myers Squibb Company; CereSpir, Inc.; Cogstate; Eisai Inc.; Elan Pharmaceuticals, Inc.; Eli Lilly and Company; EuroImmun; F. Hoffmann-La Roche Ltd and its affiliated company Genentech, Inc.; Fujirebio; GE Healthcare; IXICO Ltd.;Janssen Alzheimer Immunotherapy Research & Development, LLC.; Johnson & Johnson Pharmaceutical Research & Development LLC.; Lumosity; Lundbeck; Merck

& Co., Inc.;Meso Scale Diagnostics, LLC.; NeuroRx Research; Neurotrack Technologies; Novartis Pharmaceuticals Corporation; Pfizer Inc.; Piramal Imaging; Servier; Takeda Pharmaceutical Company; and Transition Therapeutics. The Canadian Institutes of Health Research is providing funds to support ADNI clinical sites in Canada. Private sector contributions are facilitated by the Foundation for the National Institutes of Health (www.fnih.org). The grantee organization is the Northern California Institute for Research and Education, and the study is coordinated by the Alzheimer's Therapeutic Research Institute at the University of Southern California. ADNI data are disseminated by the Laboratory for Neuro Imaging at the University of Southern California; Estudio Familiar de Influencia Genetica en Alzheimer (EFIGA): 5R37AG015473, RF1AG015473, R56AG051876; Memory & Aging Project at Knight Alzheimer’s Disease Research Center (MAP at Knight ADRC): The Memory and Aging Project at the Knight-ADRC (Knight-ADRC). This work was supported by the National Institutes of Health (NIH) grants R01AG064614, R01AG044546, RF1AG053303, RF1AG058501, U01AG058922 and R01AG064877 to Carlos Cruchaga. The recruitment and clinical characterization of research participants at Washington University was supported by NIH grants P30AG066444, P01AG03991, and P01AG026276. Data collection and sharing for this project was supported by NIH grants RF1AG054080, P30AG066462, R01AG064614 and U01AG052410. We thank the contributors who collected samples used in this study, as well as patients and their families, whose help and participation made this work possible. This work was supported by access to equipment made possible by the Hope Center for Neurological Disorders, the Neurogenomics and Informatics Center (NGI: https://neurogenomics.wustl.edu/) and the Departments of Neurology and Psychiatry at Washington University School of Medicine; National Alzheimer’s Coordinating Center (NACC): The NACC database is funded by NIA/NIH Grant U24 AG072122. NACC data are contributed by the NIA-funded ADRCs: P30 AG062429 (PI James Brewer, MD, PhD), P30 AG066468 (PI Oscar Lopez, MD), P30 AG062421 (PI Bradley Hyman, MD, PhD), P30 AG066509 (PI Thomas Grabowski, MD), P30 AG066514 (PI Mary Sano, PhD), P30 AG066530 (PI Helena Chui, MD), P30 AG066507 (PI Marilyn Albert, PhD), P30 AG066444 (PI John Morris, MD), P30 AG066518 (PI Jeffrey Kaye, MD), P30 AG066512 (PI Thomas Wisniewski, MD), P30 AG066462 (PI Scott Small, MD), P30 AG072979 (PI David Wolk, MD), P30 AG072972 (PI Charles DeCarli, MD), P30 AG072976 (PI Andrew Saykin, PsyD), P30 AG072975 (PI David Bennett, MD), P30 AG072978 (PI Neil Kowall, MD), P30 AG072977 (PI Robert Vassar, PhD), P30 AG066519 (PI Frank LaFerla, PhD), P30 AG062677 (PI Ronald Petersen, MD, PhD), P30 AG079280 (PI Eric Reiman, MD), P30 AG062422 (PI Gil Rabinovici, MD), P30 AG066511 (PI Allan Levey, MD, PhD), P30 AG072946 (PI Linda Van Eldik, PhD), P30 AG062715 (PI Sanjay Asthana, MD, FRCP), P30 AG072973 (PI Russell Swerdlow, MD), P30 AG066506 (PI Todd Golde, MD, PhD), P30 AG066508 (PI Stephen Strittmatter, MD, PhD), P30 AG066515 (PI Victor Henderson, MD, MS), P30 AG072947 (PI Suzanne Craft, PhD), P30 AG072931 (PI Henry Paulson, MD, PhD), P30 AG066546 (PI Sudha Seshadri, MD), P20 AG068024 (PI Erik Roberson, MD, PhD), P20 AG068053 (PI Justin Miller, PhD), P20 AG068077 (PI Gary Rosenberg, MD), P20 AG068082 (PI Angela Jefferson, PhD), P30 AG072958 (PI Heather Whitson, MD), P30 AG072959 (PI James Leverenz, MD); National Institute on Aging Alzheimer’s Disease Family Based Study (NIA-AD FBS): U24 AG056270; Religious Orders Study (ROS): P30AG10161,R01AG15819, R01AG42210; Memory and Aging Project (MAP - Rush): R01AG017917, R01AG42210; Minority Aging Research Study (MARS): R01AG22018, R01AG42210; Washington Heights/Inwood Columbia Aging Project (WHICAP): RF1 AG054023;and Wisconsin Registry for Alzheimer’s Prevention (WRAP): R01AG027161 and R01AG054047. Additional acknowledgments include the National Institute on Aging Genetics of Alzheimer’s Disease Data Storage Site (NIAGADS, U24AG041689) at the University of Pennsylvania, funded by NIA.

#### 1.9 Japanese GWAS Cohorts

This work was supported by the Japan Agency for Medical Research and Development (AMED) under Grant Nos. JP24dk0207060, JP21dk0207045.

***J-ADNI***

504 samples (139 ADs, 220 MCIs, and 145 controls) J-ADNI enrolled cognitively unimpaired participants, participants with late MCI, and patients with mild AD dementia using criteria consistent with those of the North American ADNI (Weiner et a. Alzheimers Dement. 2017;13(4):e1-e85).

J-ADNI was supported by the following grants: Translational Research Promotion Project from the New Energy and Industrial Technology Development Organization of Japan; Research on Dementia, Health Labor Sciences Research Grant; Life Science Database Integration Project of Japan Science and Technology Agency; Research Association of Biotechnology (contributed by Astellas Pharma Inc., Bristol-Myers Squibb, Daiichi-Sankyo, Eisai, Eli Lilly and Company, Merck-Banyu, Mitsubishi Tanabe Pharma, Pfizer Inc., Shionogi & Co., Ltd., Sumitomo Dainippon, and Takeda Pharmaceutical Company), Japan, and a grant from an anonymous Foundation.

***Clinical cohort (CL)***

Probable AD cases were ascertained on the basis of the criteria of the National Institute of Neurological and Communicative Disorders, and Stroke-Alzheimer's Disease and Related Disorders (NINCDS/ADRDA). The Mini-Mental State Examination, Clinical Dementia Rating, and/or Function Assessment Staging were primarily used for evaluation of cognitive impairment. Elders living in an unassisted manner in the local community with no signs of dementia were used as controls.

***Neuropathological cohort (NP)***

565 samples (207 cases and 358 controls) In this cohort, 358 control donors had little pathological findings associated with AD and 212 case donors had those consistent with AD. All AD patients were neuropathologically diagnosed by senile plaque and neurofibrillary tangle. No neuropathological features of other neurodegenerative disorders such as dementia with Lewy body disease, frontotemporal lobal degeneration, and Parkinson's disease, were observed. Control individuals did not show the typical neuropathological hallmarks of AD. As no clinical diagnosis is provided in this cohort, the term case or control is used in this study.

#### 1.10 GARD

The Gwangju Alzheimer's & Related Dementias (GARD) cohort is a longitudinal, single center study based in Gwangju, Republic of Korea, designed to develop clinical, imaging, genetic, and other biomarkers for the early detection and progression tracking of dementia, primarily Alzheimer's disease. Participants aged 60 years or older were recruited from the local community. The study includes standardized clinical evaluations, neuropsychological testing (Seoul Neuropsychological Screening Battery), 3T MRI imaging, and optional $^{18}$F-Florbetaben amyloid PET scans. All procedures were approved by the Institutional Review Board of Chosun University Hospital (CHOSUN 2013-12-018-070), and written informed consent was obtained from all participants or their legal representatives.

This research was supported by the Basic Research Program through the Korea Brain Research Institute (No. 23-BR-03.05), the Healthcare AI Convergence Research & Development Program through the National IT Industry Promotion Agency of Korea (No. 1711196566) funded by the Ministry of Science and ICT, and Global-LAMP Program (No. RS-2023-00285353) through the National Research Foundation of Korea funded by the Ministry of Education

#### 1.11 HK Chinese

Participants in HK Chinese cohort were recruited from the Prince of Wales Hospital, Haven of Hope Christian Service, and Community CareAge Foundation. Patients with AD were diagnosed according to the American Psychiatric Association's Diagnostic and Statistical Manual of Mental

Disorders, 5th Edition (DSM-5). This study was approved by the Clinical Research & Ethics Committees of the Joint Chinese University of Hong Kong–New Territories East Cluster for the Prince of Wales Hospital (CREC ref. no. 2015.461) and the Human Participants Research Panel of the Hong Kong University of Science and Technology (CRP#180 and CRP#225). All participants provided written informed consent for both study enrolment and sample collection.

### 2 eQTL analysis datasets

**ROSMAP RNAseq and WGS datasets**

The data available in the AD Knowledge Portal would not be possible without the participation of research volunteers and the contribution of data by collaborating researchers. The results published here are in whole or in part based on data obtained from the AD Knowledge Portal (https://adknowledgeportal.org). Study data were provided by the Rush Alzheimer's Disease Center, Rush University Medical Center, Chicago. Data collection was supported through funding by NIA grants P30AG10161, R01AG15819, R01AG17917, U01AG46152, U01AG046170, U01AG046139, and U01AG61356. Additional phenotypic data can be requested at www.radc.rush.edu.

### 3 Supplementary discussion

Limitations of the study

In most datasets AD case status was based on medical records and an ICD based definition and in one cohort the use of Alzheimer's disease medication was additionally included to define AD status. This may have induced a misclassification bias, and may potentially lead to identification of loci that are not related to true AD or may, due to the underdiagnosis of AD in general, lead to conservative estimates in the cohorts. The first issue is to some extend mitigated by using a meta-analysis setup where cohorts with different AD definitions were used and the requirement that a signal needed to be present in multiple cohorts.

Furthermore, we have excluded the *APOE* ε42 strata from all analyses and this exclusion may have induced an ascertainment bias, and the identified interactions between a genetic variant and *APOE* genotypes may not extend to individuals in the *APOE* ε42 strata. Also, we have a considerable residual population stratification which may also have induced a bias in the analyses.

Influence of age

Because late-onset AD is strongly associated with age, we adjusted all analyses for age. To ensure that bias due to heterogeneity in selection processes of cases and controls across studies was not a major challenge, we performed sensitivity analyses of the eight main cohorts where age was not adjusted. Similar findings were observed when comparing analyses with and without age adjustment. Despite differences in age distributions between cases and controls and between *APOE* strata, they were not of substantial size to influence the main observations.

The signals that were augmented in the *APOE* ε33 strata (and thus attenuated in ε44+ε43), *SLC50A1*, *TMEM106B*, *NPAS3* and *SHARPIN*, might be influenced by the effect of statistical power and age differences discussed above. For example, *TMEM106B* associates with the tauopathy-endophenotype which has a later mean onset compared to the *APOE*-related-AD endophenotype[1]. Because the ε33 strata display a higher mean age the observed interaction could be driven by this mean age difference. On the other hand, a clear attenuation of the effect on risk of AD in an *APOE* ε4 allele context was observed. Future mechanistic studies would be important to further scrutinize this finding, and to substantiate if the *APOE* ε4 allele biologically interacts with the tauopathy-endophenotype. Similar considerations are valid for *SLC50A1* and *SHARPIN*. Conversely, *NPAS3* has been reported to modify *reelin* expression and *reelin* binds to both the VLDL- and APOER2-receptors to which apoE is also a ligand. It is known that the apoE4 isoform has a higher binding affinity to these receptors[2,3]. Hence, in this situation it may be plausible that the *APOE* ε4 allele confers a protective effect on AD risk conferred by *NPAS3* and the reeling pathway.

<u>Interaction analysis</u>
Interaction analysis is notoriously difficult to perform convincingly as it constitutes an extra level of complexity compared to a normal association study. It is however of clear importance to determine if the observed differences across *APOE* strata with some certainty can be judged true. Our approach was to use several layers of evidence to claim an interaction.

We used two complementary approaches to test for SNP x *APOE* interactions: a genome-wide SNP x *APOE* ε4 carrier interaction GWAS meta-analysis and a stratified effect comparison. The SNP x *APOE* ε4 carrier interaction GWAS is standard for testing gene-gene or gene-environment interactions as the different terms in the model are evaluated under the same statistical framework with a proper variance structure and correlation modeling. However, the methodology requires high statistical power, and the interpretation of the results can be challeging.. The stratified effect comparison difference method requires less statistical power, has a clear interpretation and allows for direct visualization of the stratified effects. However, it assumes independence between the strata, does not formally model the interaction term in a single framework, may identify differences driven by power asymmetry between strata, and has multiple testing considerations. Performing both complementary interaction strategies allows a more nuanced interpretation.

To address the limitation in the stratified effect comparison approach, we only considered the lead variants in the loci that reached genome wide significance in either the ε33 or the ε44+ε43 strata. This was to avoid data dredging and only to consider genetic variants which had sufficient statistical power. For each lead variant we have provided the Forest plot of the effect on risk for each stratum. This allows scrutinization of tendencies across *APOE* strata and visual inspection of confidence interval overlaps and stepwise effects as a function of number of ε4 alleles. We did not use a simple t-test to test for overlapping confidence intervals because we have performed a meta-analysis across several cohorts with different diagnostic criteria, different cohort types (case-control vs biobanks), different allele frequencies and potential differences between strata due to relatedness. Hence, using different fixed- and mixed-effect models to investigate the possible interactions between *APOE* ε4 and lead variants seem appropriate. Further, the Forest plots showing the effect difference between ε44+ε43 and ε33 strata indicate that a possible bias due to relatedness between strata in some of the cohorts is minimal. Unfortunately, the SNP x *APOE* ε4 carrier interaction GWAS meta-analysis was underpowered to find significant interactions, but for a few SNPs the interaction GWAS results support at a suggestive level the main *DDHD1* finding

The *APOE* ε33 and ε44+ε43 strata are comparable in power, however we cannot exclude that some of the observed differences among these two strata could be explained by differences in power. A difference in statistical power would however mostly affect the confidence interval size and therefore the p-value, whereas the effect size (beta) should remain relatively stable. In contrast, and in support of our interaction analysis strategy, we consistently observe large shifts in the effect size when an interaction is present between a lead variant and *APOE* ε4 carrier status.

In the main dominant stratified effect comparison analysis where the effect difference and SE are calculated within each cohort, the dataset only contains 8 rows – one from each of the cohorts. In the additive tests and the dominant-sensitivity test, the datasets contain 18 and 24 rows – one for each cohort+strata. This difference leads to the higher observed heterogeneity.

Although we find significant stratified effect comparison differences, the analysis is affected by limited statistical power even when using eight cohorts. This can for example be seen in the patterns for the *TMEM106B*, *GRN* and *MAPT* loci only being significant in the *APOE* ε33 strata. They are all known to associate with the tauopathy dementia endophenotype, but only *TMEM106B* has a significant stratified effect comparison test. However, the tests for *GRN* and *MAPT* show a similar trend and will possibly become significant if statistical power is increased.

Summary data-based Mendelian Randomization (SMR)

*STAG3* has previously been reported in SMR in previous studies using non-stratified summary GWAS data[4,5]. In our study, it was only significant in the ε44+ε43 stratum, however there was a non-significant signal in the ε33 stratum indicating some shared effects between the strata. The *LRRC37A*, *ARL17B* and *LRRC37A2* are novel findings all located in the *MAPT* locus. The novel loci nominated by the stratified *APOE* meta-analyses were not present in the MetaBrain dataset for testing and would need further investigation to determine whether the expression of these genes influences disease risk.

#### References for Supplementary discussion

1. Shade, L.M.P. *et al.* GWAS of multiple neuropathology endophenotypes identifies new risk loci and provides insights into the genetic risk of dementia. *Nat Genet* **56**, 2407-2421 (2024).
2. Erbel-Sieler, C. *et al.* Behavioral and regulatory abnormalities in mice deficient in the NPAS1 and NPAS3 transcription factors. *Proc Natl Acad Sci U S A* **101**, 13648-53 (2004).
3. Stranahan, A.M., Erion, J.R. & Wosiski-Kuhn, M. Reelin signaling in development, maintenance, and plasticity of neural networks. *Ageing Res Rev* **12**, 815-22 (2013).
4. Alvarado, C.X. *et al.* omicSynth: an Open Multi-omic Community Resource for Identifying Druggable Targets across Neurodegenerative Diseases. *medRxiv* (2023).
5. Pyun, J.M. *et al.* Transcriptional risk scores in Alzheimer's disease: From pathology to cognition. *Alzheimers Dement* **20**, 243-252 (2024).
